# Beyond Volume: Unraveling the Genetics of Human Brain Geometry

**DOI:** 10.1101/2024.06.25.24309376

**Authors:** Sabrina A. Primus, Felix Hoffstaedter, Federico Raimondo, Simon B. Eickhoff, Juliane Winkelmann, Konrad Oexle, Kaustubh R. Patil

## Abstract

Brain geometry impacts brain function. A quantitative encoding of form is provided by the Laplace-Beltrami operator’s spectrum of eigenvalues (LBS). We examined LBS genetics of 22 subcortical brain structures including cerebellum in 19,862 healthy White-British UK Biobank participants by multivariate GWAS (MOSTest) on the first 49 eigenvalues each. Controlling for surface and volume, we identified 80 unique variants (p<1/22*5E-8) influencing the shapes of one or several structures, with the highest yield (37 variants) for brain stem. The previously known influence of several of these loci on basic morphology, such as volume, is thus shown to also influence complex shape. Known associations of observed loci with blood pressure, neurodegeneration, alcohol consumption, and mental disorders hint at preclinical stages of these conditions potentially mediating the genetic effect on brain morphology. Significant correlations between LBS of several brain structures and the polygenic risks of hypertension, ischemic stroke and schizophrenia evince brain shapes as early biomarkers.

## Introduction

The human brain comprises an intricate constellation of diverse substructures, each of which has specific functions and forms complex interactions with other parts of the brain. Investigating the morphological properties of these substructures and deciphering their genetic underpinnings is imperative for advancing our understanding of the human brain in health and disease. Previous genetic studies have investigated crude parameters such as volume and surface area (*1–5*) The genetics of the intricate shapes of brain structures remain largely unexamined, however, representing a gap in comprehensive understanding of brain anatomy.

The structures within the human brain exhibit a range of shapes, from the more spherical amygdala to the elongated hippocampus. Basic metrics such as volume and surface area do not adequately capture the nuanced details of these shapes and thus fall short of providing a sufficient description of their morphology. There is evidence for a high inter-individual variance in these intricate details, with a substantial proportion being heritable (*6–8*). Initial efforts to delve into the genetic underpinning of this heritable aspect used techniques like voxel-based analysis in combination with dimension reduction methods (*9*, *10*), or global-to-local representations of brain shapes (*11*, *12*). Each of these techniques carries its own challenges, including reduced robustness and accuracy due to suboptimal image registration, high computational effort, or a lack of physical interpretability due to dimensionality reduction or a prohibitive dimensionality of image-derived features.

The Laplace-Beltrami operator provides a multivariate spectral representation of a shape, capturing its characteristics in detail (*13*, *14*). The Laplace-Beltrami spectrum (LBS), a set of ordered eigenvalues, is obtained by solving the Helmholtz equation, a time-independent form of the wave equation, on a Riemannian manifold which can be the surface of a given brain structure. Solutions are decomposed into eigenfunctions (also referred to as “eigenmodes”) and their corresponding eigenvalues representing the natural vibrations and their squared frequencies, respectively, on the manifold underlying the shape, akin to the harmonic frequencies of the membrane of a drum. Each sound or other vibration of a membrane can be represented as a weighted sum of its eigenmodes, relating closely to the theory of spectral analysis in Fourier series. Necessarily, the eigenmodes are strongly linked to a shape’s geometry, as visualized in a seminal experiment by Ernst Chladni using vibrating plates (*15*). It has been asked since then whether a shape can be represented by a unique series of eigenvalues or, metaphorically, whether the shape of a drum can be heard (*16*). While the answer is affirmative for one-dimensional shapes whose lengths are made audible by the harmonics or pure tones of string instruments, for instance, this is not true in general for shapes with more than one dimension as there exist isospectral shapes (*17*). Nevertheless, the known counterexamples appear to be rare (*13*) and under certain symmetry assumptions, a unique assignment is possible (*18*). Consequentially, a relatively small subsequence (e.g. 50 eigenvalues) of the increasingly ordered spectrum contains enough geometric information to uniquely describe a shape by adequately capturing its curvatures (*13*, *19–21*), although the spectrum usually contains an infinite number of eigenvalues. Following Ge et al. (*6*), we refer to this subsequence as the LBS (also known as Shape-DNA (*13*)). This multidimensional intrinsic shape representation, as an isometric invariant, is independent of rotation, translation, and scaling of the coordinate system, eliminating the need for error-prone inter-individual image registration (*22*), and behaves continuously with any change in the manifold. Recently, there is considerable interest in using geometric eigenmodes to explain shape-associated biological mechanisms (*23*). The LBS appears to be a more straightforward shape descriptor, however, which is computed efficiently (*21*) and thus well suited for large-scale genome-wide association studies (GWASs) which require a quantitative representation of the shape of brain structures at the individual level.

By performing GWAS, we aimed to reveal information on gene loci contributing to the heritability of brain morphology as quantified by LBS. We derived the LBSs of 22 different brain structures from a large MRI dataset provided by the UK Biobank (UKB). To reveal shape-specific signals, we controlled for global characteristics such as brain volume and surface area. Since all eigenvalues in an LBS contribute to the description of the respective shape, it is important to study them jointly while accounting for their mutual dependencies. To achieve this, we employed the state-of-the-art multivariate GWAS tool MOSTest (*2*), which accounts for the pairwise correlations between eigenvalues and, by considering their joint distribution, has increased power for detecting genetic associations. Our study focused on subcortical structures, brain stem and cerebellum, in keeping with several studies which studied the genetics of their volumes (*2*, *5*, *24–26*). These parts of the brain are not only involved in learning and decision-making processes (*27*) but also hotspots of various brain disorders (*7*, *24–26*, *28*). We identified specific genetic influences on their shapes, investigated genetic asymmetries and similarities among structures, obtained precise estimates of the heritability of these shapes, and performed genetic correlation and enrichment analyses with respect to biological pathways, traits, and diseases.

## Results

### Multivariate Genome-wide Analyses

MOSTest on the LBS of each of the 22 brain structures in 19,862 healthy, unrelated, White-British individuals (10,427 female, with mean age±SD of 64.3±7.4 years) yielded a total of 148 significant SNPs (Bonferroni-corrected for multiple testing p < 1/22 * 5E-8 = 2.27E-9) which were each independent (linkage disequilibrium (LD) of r2 < 0.6) in their respective LBS GWAS. Some of them were significant in multiple brain structures. Thus, across all 22 GWASs, there were 80 unique SNPs independently associated with the shape of at least one brain structure (Table S1, Fig. S1-S3). Using FUMA (*29*) for clumping the results of each brain structure (see Methods), we identified 62 genomic risk loci in total, of which 48 were shared by at least 2 of the 22 brain structures (Fig. 1). The largest number of independent significant SNPs were found for brain stem (37) (Fig. 2B), followed by left cerebellum white matter (29). Only the amygdala did not show any significant signals after Bonferroni correction. The strongest signal was observed for lead SNP rs13107325 on chromosome 4 with a p-value of 2.15E-74 in association with the LBS of the left cerebellum white matter. This SNP association was observed most frequently, appearing in 14 of the 22 brain structures.

**Fig. 1.**
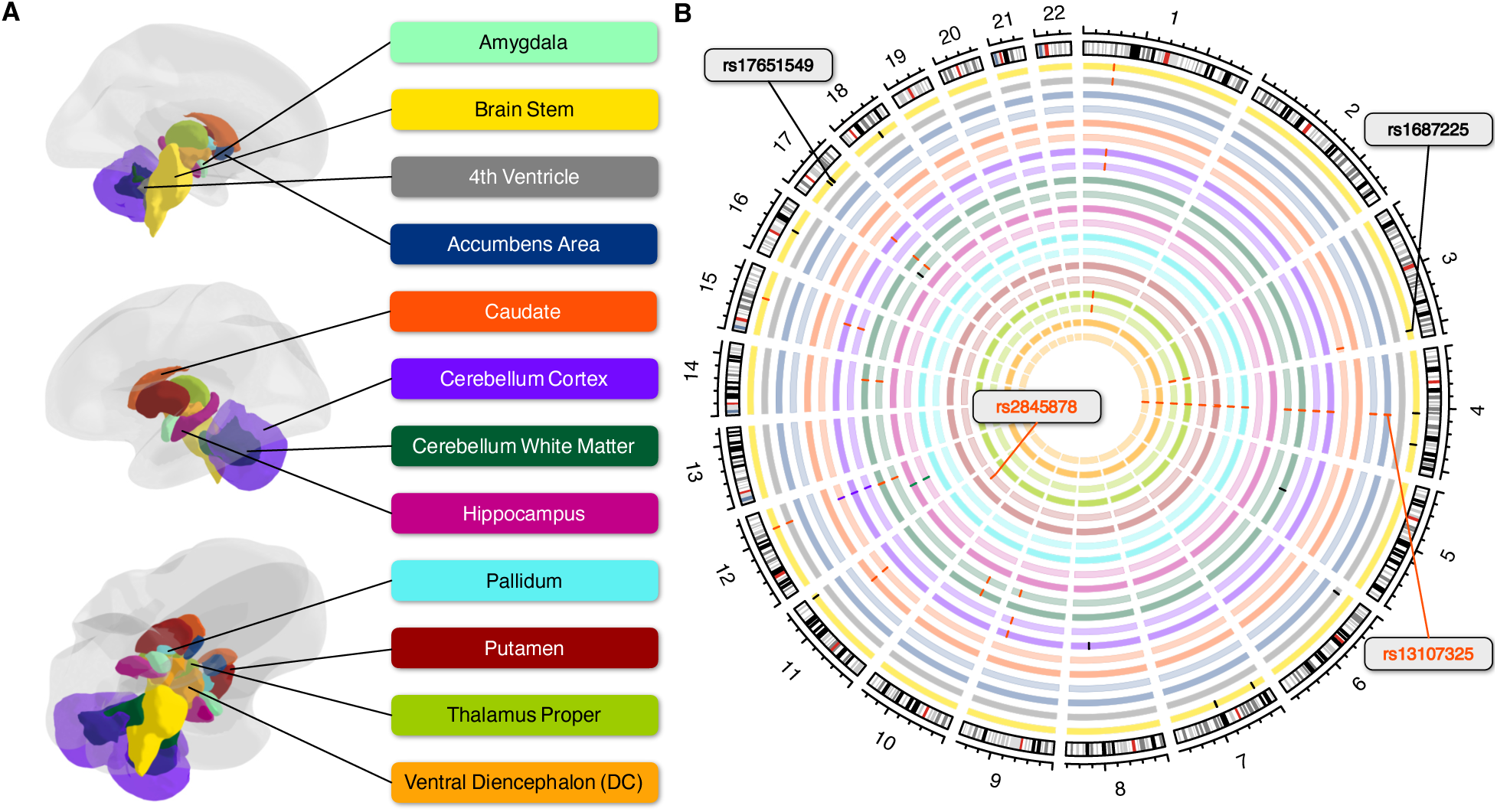
Overview of Genomic Risk Loci in Different Brain Structures. **A:** Anatomical representation of the subcortical brain structures assessed in the present study. **B:** Circular plot indicating the genomic risk loci derived by MOSTest GWAS on each brain structure. Bilateral structures are on neighboring circles and are the same color as in A, with the right-side structure represented more centrally and lighter. If multiple structures share a locus, the bars indicating their position on the respective circles are the same color.

**Fig. 2.**
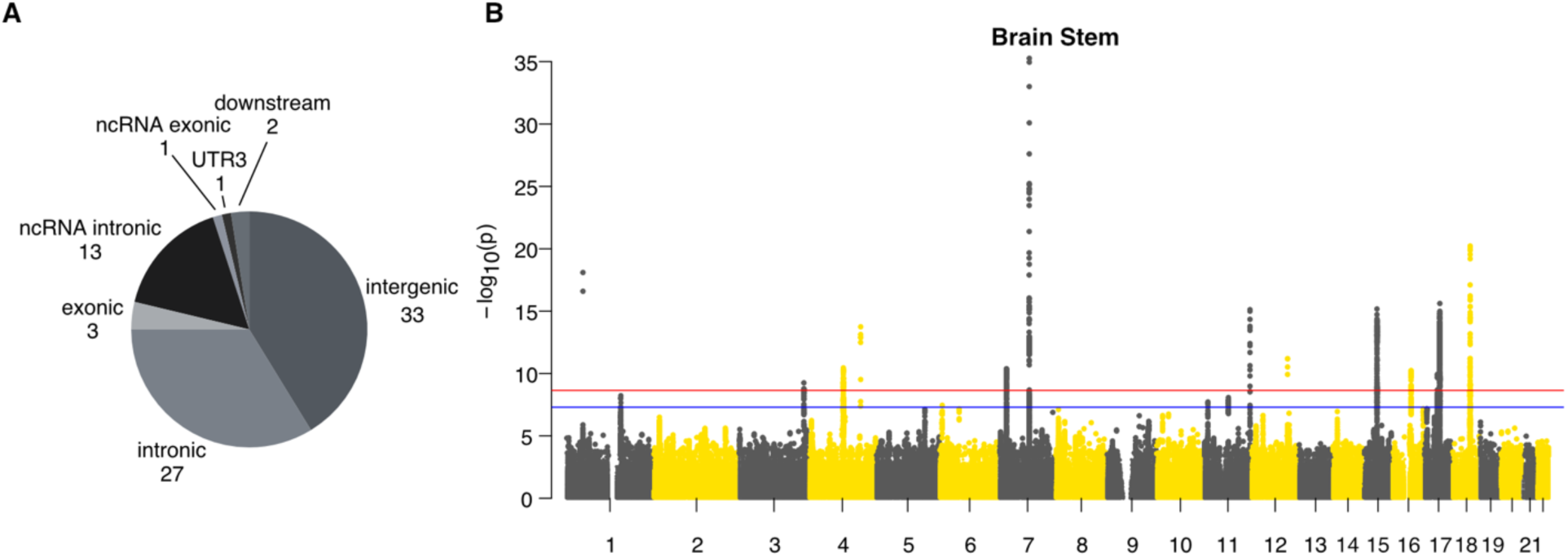
SNP-based Associations with the LBS. **A:** Pie chart showing the frequency of functional annotations of all 80 independent significant SNPs. **B:** Manhattan plot of multivariate MOSTest results of the brain stem with corrected and non-corrected genome-wide significant threshold (p=2.27E-9 (red line), p = 5E-8 (blue line)).

49 of the 80 independent SNPs, either themselves or via a proxy variant (*r*2 ≥ 0.6), have previously been identified in GWASs on brain shape-related traits (e.g., volume, surface area, or cortical thickness) and are listed in the GWAS catalog implemented in FUMA. Of these 80, 13 have been associated exclusively with other traits (not brain shape-related), and 18 were not related to any GWAS catalog entry (Table S1).

### Functional Annotations of Genomic Risk Loci

We annotated our results using ANNOVAR (*30*) as implemented in FUMA. Of the 80 independent significant SNPs, 3 were exonic, 27 were intronic, and 33 were in intergenic regions (Fig. 2A, Table S2). All 3 exonic variants (rs13107325, rs1687225, rs601558) were nonsynonymous, each having a combined annotation-dependent depletion (CADD) score higher than 15.23 and belong, therefore, to the 3% most deleterious SNPs (*31*). We further found 28 exonic nonsynonymous (ExNS) variants in LD (*r*2 ≥ 0.6) with one of the independent significant SNPs, with 17 of them having a CADD score higher than 15.23 or a Regulome DB score (RDB; small scores indicating a high likelihood of being a regulatory SNP) lower than 2 (Table 1, Table S3).

**Table 1.** Exonic Nonsynonymous Variants in LD (*r*2 ≥ 0.6) with one of the Independent Significant SNPs Across all Brain Structures. Variants are listed with non-effect allele (NEA), effect allele (EA), and effect allele frequency (EAF), minimal significance level (minP) across all structures, CADD and RDB scores, most likely affected gene, and the independent significant SNPs (IndSigSNP) to which they are linked most strongly as indicated by the correlation value r2. The respective IndSigSNP may differ between brain structures according to the structure-specific GWAS results. Blue-marked variants have CADD > 15.23 or RDB < 2. Bold typing of a variant indicates genome-wide significance in at least one brain structure. Variants in italics are independent significant SNPs.

| rsID | Position | EA | NEA | EAF | MinP | CADD | RDB | Gene | Brain Structure (IndSigSNP, r2) |
| --- | --- | --- | --- | --- | --- | --- | --- | --- | --- |
| **rs1687225** | chr3:183948663 | A | G | 0.23 | 6E-10 | 19.0 | 4 | VWA5B2 | Brain Stem (**rs1687225**, 1.00) |
| rs902417 | chr3:183951431 | T | C | 0.25 | 7E-08 | 15.4 | 5 | VWA5B2 | Brain Stem (**rs1687225**, 0.86) |
| rs3733197 | chr4:102839287 | A | G | 0.35 | 2E-06 | 14.2 | 7 | BANK1 | Left Cerebellum White Matter (**rs114614648**, 0.72) |
| **rs13107325** | chr4:103188709 | T | C | 0.08 | 2E-74 | 23.1 | 5 | SLC39A8 | Accumbens area (right + left: **rs13107325**, 1.00);<br>Cerebellum Cortex (right + left: **rs13107325**, 1.00);<br>Cerebellum White Matter (right + left: **rs13107325**, 1.00);<br>Pallidum (right + left: **rs13107325**, 1.00);<br>Putamen (right + left: **rs13107325**, 1.00);<br>Ventral DC (right + left: **rs13107325**, 1.00);<br>Thalamus Proper (right: **rs13107325**, 1.00; left: **rs13135092**, 0.93) |
| rs7800072 | chr7:84628989 | G | T | 0.32 | 3E-08 | 18.3 | 5 | SEMA3D | Brain Stem (**rs10247311**, 0.90) |
| **rs601558** | chr8:108970367 | G | A | 0.34 | 8E-11 | 22.0 | 5 | RSPO2 | Left Cerebellum Cortex (**rs601558**, 1.00) |
| rs36045050 | chr14:69257858 | T | C | 0.26 | 8E-09 | 7.0 | 2b | ZFP36L1 | Cerebellum White Matter (right: **rs1547050**, 0.74; left: **rs12435718**, 0.73) |
| **rs4646626** | chr15:58256127 | T | C | 0.46 | 6E-15 | 19.7 | NA | ALDH1A2 | Brain Stem (*rs1061278*, 0.91); Cerebellum Cortex (right: *rs3742960*, 0.92; left: *rs3742959*, 0.92) |
| rs1877031 | chr17:37814080 | A | G | 0.33 | 1E-06 | 23.1 | NA | STARD3 | Brain Stem (*rs2271308*, 0.80) |
| rs1058808 | chr17:37884037 | G | C | 0.33 | 1E-07 | 23.5 | 5 | ERBB2 | Brain Stem (*rs2271308*, 0.64) |
| rs12949256 | chr17:43507297 | T | C | 0.19 | 1E-08 | 15.0 | 4 | ARHGAP27 | Brain Stem (*rs568589031*, 0.70) |
| **rs16940674** | chr17:43910507 | T | C | 0.24 | 1E-13 | 17.6 | 1f | CRHR1 | Brain Stem (*rs568589031*, 0.96) |
| **rs16940681** | chr17:43912159 | C | G | 0.24 | 1E-13 | 5.0 | 4 | CRHR1 | Brain Stem (*rs568589031*, 0.96) |
| **rs62621252** | chr17:43922942 | C | T | 0.24 | 6E-14 | 6.3 | 5 | SPPL2C | Brain Stem (*rs568589031*, 0.96) |
| **rs62054815** | chr17:43923266 | A | G | 0.24 | 1E-13 | 0.0 | 5 | SPPL2C | Brain Stem (*rs568589031*, 0.96) |
| **rs12185233** | chr17:43923654 | C | G | 0.24 | 5E-14 | 23.5 | 1f | SPPL2C | Brain Stem (*rs568589031*, 0.95) |
| **rs12185268** | chr17:43923683 | G | A | 0.24 | 1E-13 | 8.7 | 1f | SPPL2C | Brain Stem (*rs568589031*, 0.96) |
| **rs12373123** | chr17:43924073 | C | T | 0.24 | 1E-13 | 23.3 | 1f | SPPL2C | Brain Stem (*rs568589031*, 0.96) |
| **rs12373139** | chr17:43924130 | A | G | 0.24 | 1E-13 | 4.0 | 1f | SPPL2C | Brain Stem (*rs568589031*, 0.96) |
| **rs12373142** | chr17:43924200 | G | C | 0.24 | 7E-13 | 7.5 | 1f | SPPL2C | Brain Stem (*rs568589031*, 0.96) |
| **rs63750417** | chr17:44060775 | T | C | 0.24 | 1E-13 | 11.9 | 5 | MAPT | Brain Stem (*rs568589031*, 0.96) |
| **rs62063786** | chr17:44061023 | A | G | 0.24 | 9E-14 | 7.9 | 5 | MAPT | Brain Stem (*rs568589031*, 0.96) |
| **rs62063787** | chr17:44061036 | C | T | 0.24 | 1E-13 | 0.2 | 5 | MAPT | Brain Stem (*rs568589031*, 0.96) |
| **rs17651549** | chr17:44061278 | T | C | 0.24 | 1E-13 | 26.8 | 1f | MAPT | Brain Stem (*rs568589031*, 0.96) |
| **rs10445337** | chr17:44067400 | C | T | 0.24 | 2E-13 | 19.2 | 1f | MAPT | Brain Stem (*rs568589031*, 0.96) |
| **rs62063857** | chr17:44076665 | G | A | 0.24 | 1E-13 | 1.5 | 7 | STH | Brain Stem (*rs568589031*, 0.96) |
| **rs34579536** | chr17:44108906 | G | A | 0.24 | 2E-13 | 14.4 | 3a | KANSL1 | Brain Stem (*rs568589031*, 0.96) |
| **rs34043286** | chr17:44117119 | G | A | 0.24 | 1E-13 | 21.0 | 4 | KANSL1 | Brain Stem (*rs568589031*, 0.96) |

The pleiotropic missense variant rs13107325, as mentioned above, affects the metal transporter *SLC39A8* and is, with a CADD score higher than 20 (23.1), among the 1% most deleterious SNPs. While ClinVar (*32*) and AlphaMissense (*33*) classify this pleiotropic variant as benign, it is a well-known risk factor for schizophrenia (*34–36*) and has also been found in conjunction with inflammation-based diseases like Crohn’s disease (*37*), blood pressure (*38*) as well as brain imaging phenotypes (*9*).

*VWA5B2* on chromosome 3 comprises the ExNS missense variant rs1687225, an independent significant SNP associated with brain stem LBS (p=5.6E-10). The CADD score of this variant is relatively high (19), but this SNP is not reported in ClinVar and AlphaMissense predicts it to be likely benign. Of note, the variant is a brain expression quantitative trait locus (eQTL) for different genes and an independent brain cis-eQTL for *VWA5B2* itself according to GTEx (v.8) (Table S4).

The ExNS missense variant rs601558 on chromosome 8 was significantly associated with the LBS of the left cerebellum cortex (p=8.0E-11) and is contained in *RSPO2*, for which it is also a brain eQTL. Despite a high CADD score of 22, this variant is classified as benign by ClinVar and AlphaMissense.

Despite its benign classification by ClinVar and AlphaMissense, rs17651549 in *MAPT* on chromosome 17, which was associated with the brain stem LBS at p=1.0E-13, had the highest CADD score (26.8) among the ExNS variants. It was in nearly complete LD (r2=0.96) with the lead SNP rs568589031 (brain stem, p=2.4E-16) of this large genomic risk locus that included altogether 6 ExNS SNPs which, according to CADD score, all belong to the 3% most deleterious SNPs (Table 1) and act as brain eQTLs of *CRHR1*, *SPPL2C*, *MAPT* and *KANSL1*. Due to its high linkage, this large genomic region, also referred to as *MAPT* locus, is known to be highly complex (*39*) and we observed 18 ExNS SNPs being brain eQTLs of 17 protein-coding genes in total (Table S5).

### Asymmetry and Similarity of Brain Structures

To assess possible genetic asymmetries and similarities of brain structures, we compared the p-values of the 80 unique independent SNPs between structures and hemispheres (Table S6). Applying a conservative Bonferroni correction for 80*22 comparisons to the nominal threshold of 0.05, we found five genome-wide significant (p<2.27E-9) SNPs for cerebellum white matter and one for putamen that were not significant in the respective contralateral structures. At nominal significance, only the intron variant rs2845878 in *FAT3*, which came up in the GWAS on right putamen, lacked symmetry. Proxy variants of rs2845878 (*r*2 ≥ 0.6) have been linked to the mean volume of caudate and putamen according to GWAS catalog (*4*). Principal component (PC) analysis of -log10 transformed p-values of these 80 SNPs showed a high level of symmetry and similarity in genetic signals (Fig. 3): All subcortical structures as well as the cerebellum cortex were mapped close to each other in this PC space, independent of their hemispherical assignment. Moreover, most structures shared genetic architecture according to their placement adjacent to each other as in case of the basal ganglia, for instance. However, brain stem, cerebellum white matter and hippocampus were substantially distanced from the others in at least one of the first three PCs. Hippocampus was probably notable for its dissimilar pattern of few associated SNPs while the particular position of brain stem and cerebellum white matter could be due to their highly significant signals in addition to their quantity.

**Fig. 3.**
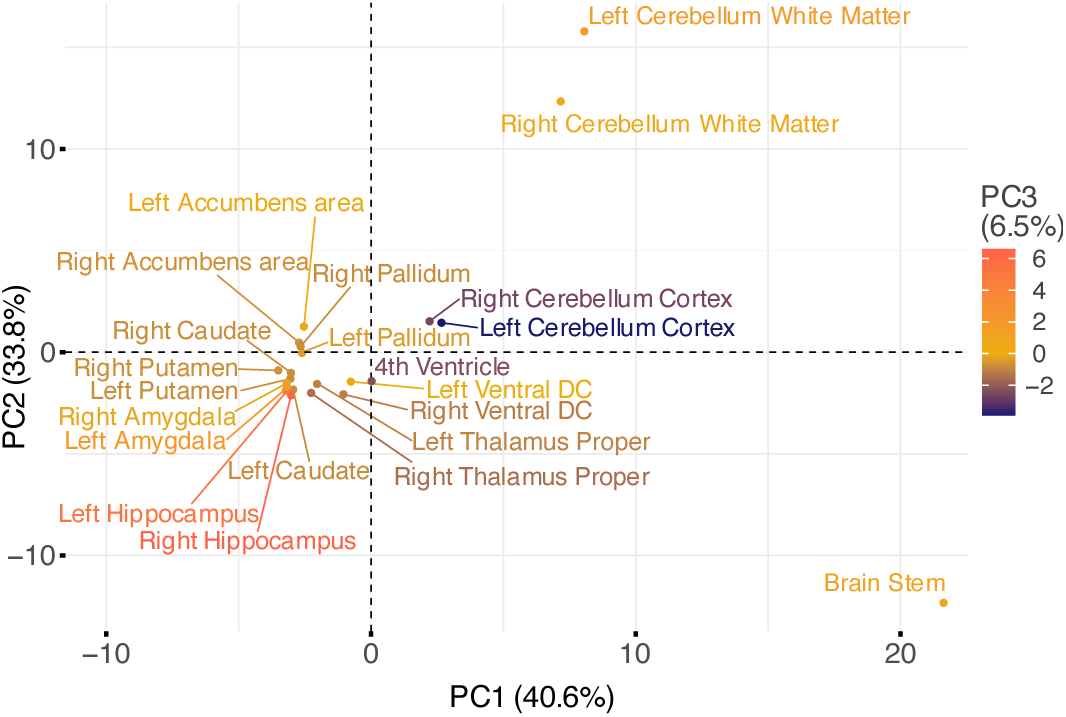
PCA of -log10 scaled p-values of the 80 independent significant SNPs across all structures. Plot shows the first two PCs with corresponding explained variance in brackets behind. Structures are colored according to the third principal component.

When investigating genetics that were similar for several structures, the ExNS variant rs13107325 in *SLC39A8* along with several others within the same risk locus on chromosome 4, the intergenic SNP rs6658111 on chromosome 1 and the intronic variant rs12146713 in *NUAK1* on chromosome 12 emerged as the most frequently shared independent SNPs. After Bonferroni correction for 80*22 comparisons, these three were significant in 10 to 15 structures and nominally significant in 19, respectively, 20 structures (Table S6). Apart from the pleiotropic rs13107325, the other two variants have so far only been linked to various brain shape traits such as volume, surface area and cortical thickness according to the GWAS catalog (Table S7). Interestingly, rs12146713 and rs13107325 have also recently been linked to structural connectivity measures (*40*) which supports the notion that they have an overarching effect on brain structuring.

### Gene-mapping and Gene-enrichment Analysis

We first annotated all SNPs passing QC to 18383 protein-coding genes using MAGMA (*41*) and calculated a p-value for each gene by applying the SNP-wise mean model (see Methods). The brain stem stood out with 20 protein-coding genes being significantly associated with its shape in the gene-based analysis after Bonferroni correction for 22 brain structures and 18383 protein-coding genes (p < 0.05 / (22*18383)) (Fig. 4A). Here, *CRHR1* showed the strongest signal (p= 5.2E-15). This gene belongs to the extended *MAPT* locus mentioned above, which came up as generally highly significant and has, together with *MAPT*, which encodes for the microtubule-associated protein tau, often been associated with neurodegenerative disorders (*42*, *43*).

**Fig. 4.**
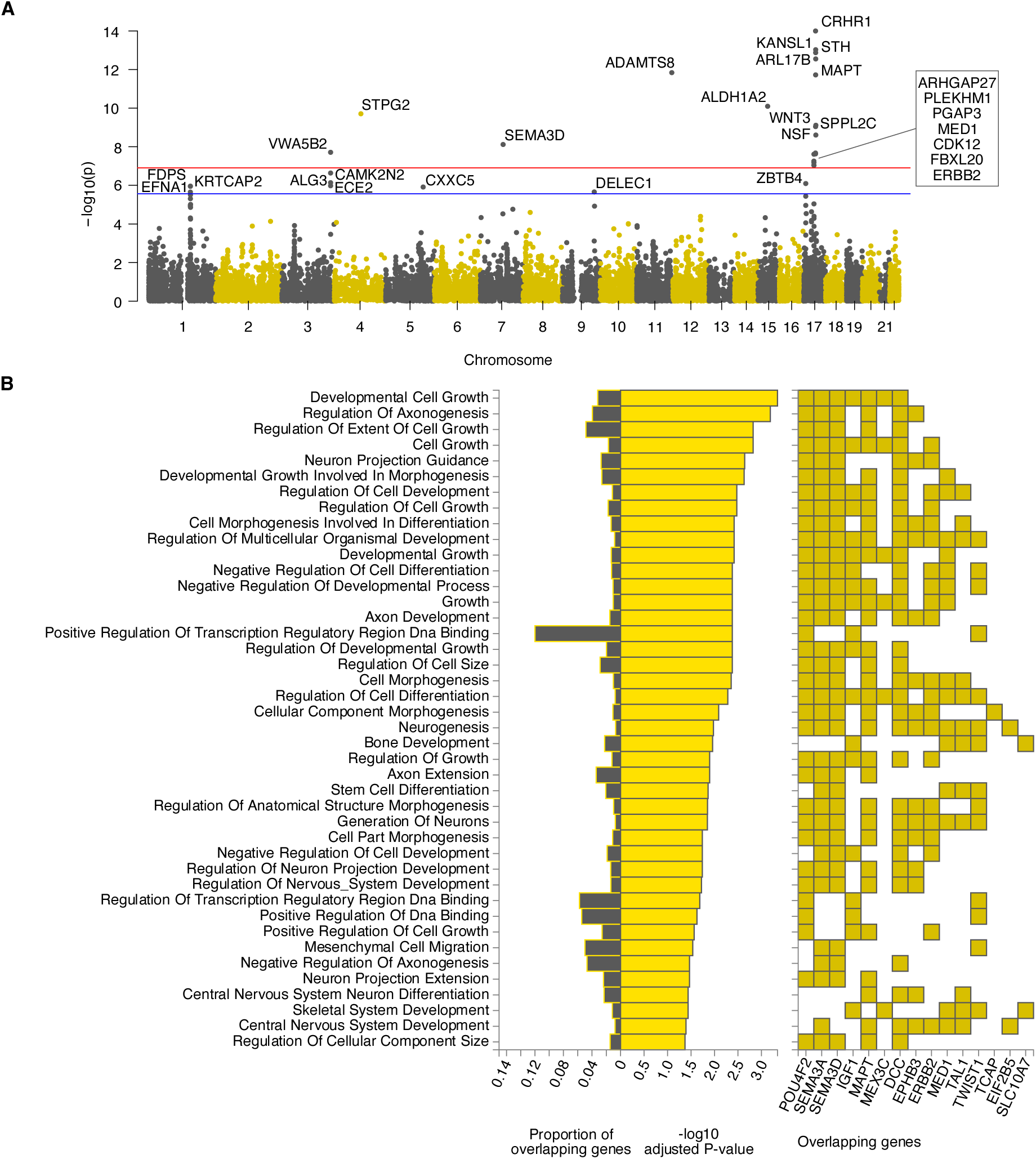
Gene Analysis and Enrichment in Brain Stem. **A:** Manhattan plot of MAGMA gene analysis of brain stem results using SNP-wise mean model. **B:** Hypergeometric tests of overrepresentation and overlap of prioritized genes with predefined gene sets using FUMA Gene2Func. Overlap with gene sets of GO biological processes (MsigDB c5). P-values were corrected by the Benjamini-Hochberg method for multiple testing.

Next, we used PoPS (*44*) for gene prioritization, a tool that assigns polygenic priority scores to each gene by fitting their MAGMA z-scores based on trait-relevant gene features extracted from cell-type-specific gene expressions, biological pathways, and protein-protein interactions. Since PoPS works best when combined with orthogonal methods, we also mapped each SNP to the nearest genes. Reranking the genes according to the mean of the two ranks that resulted from the two methods, we prioritized all genes with rank ≤ 2 at each locus (see Methods, Table S8).

Based on these genes, a gene set enrichment analysis was conducted using FUMA’s Gene2Func method. For bilateral brain structures the two gene sets were joined in this analysis due to similar genetic architectures on both sides (see above and Fig. 3). Afterwards, the overlap and degree of overrepresentation of all prioritized genes in predefined gene sets were examined.

At the locus of the most significant SNP in this study, rs13107325, two genes were usually prioritized, *BANK1* and *SLC39A8* (Table S8). The previously known pleiotropy of this genomic region was confirmed by the diversity of our enrichment results which included adventurousness, hypertension, multisite chronic pain, general cognitive ability, and alcohol consumption (Fig. 5A, Fig. S4-S5 for all other structures). Especially the overlap with alcohol consumption was most prominent.

**Fig. 5.**
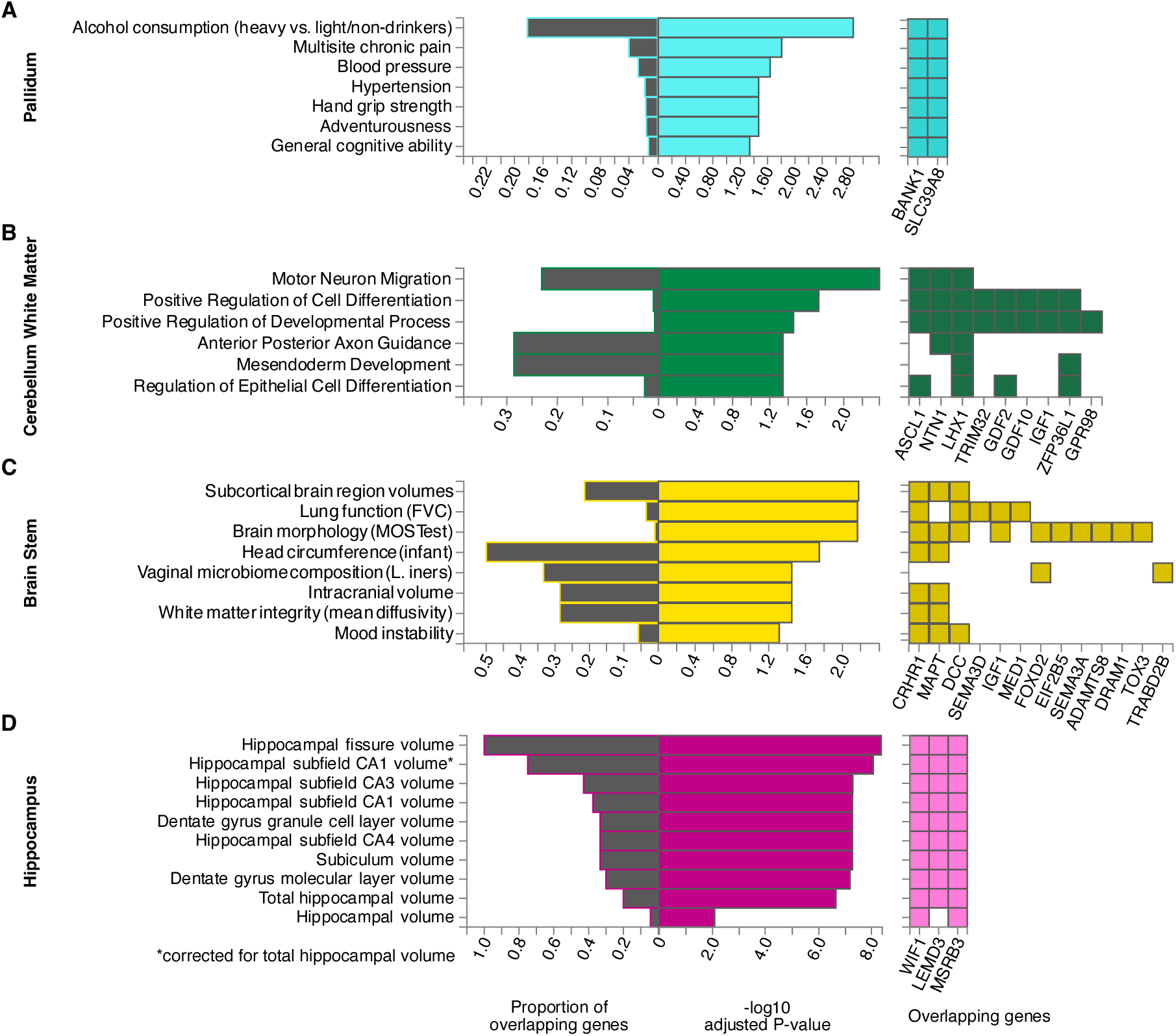
Significant Gene Enrichment Results of Selected Brain Structures. Hypergeometric tests of overrepresentation and overlap of prioritized genes with predefined gene sets from GWAS catalog traits (**A**, **C, D**) and GO biological processes (**B**). Pallidum (**A**) was chosen as a representative for structures prioritizing *BANK1* and *SLC39A8* at the pleiotropic locus of rs13107325. All other structures can be found in the supplements. Cerebellum white matter (**B**), hippocampus (**C**) and brain stem (**D**) showed a substantially different genetic pattern before (Fig. 3). P-values were corrected by the Benjamini-Hochberg method for multiple tests in each category.

These two genes and 23 others were prioritized for the cerebellum white matter. However, given this entire set of genes, *BANK1* and *SLC39A8* were not significantly overrepresented in any gene set. Instead, the other genes, which were primarily prioritized by the cerebellar white matter, were enriched in several regulatory pathways associated with neurological and developmental processes (Fig. 5B).

For brain stem, Gene2Func identified gene sets linked to brain morphology and lung function according to GWAS catalog (Fig. 5C) as well as multiple biological processes from the Gene Ontology Resource (GO) involved in cell and nervous system development and regulation (Fig. 4B). Brain-stem-shape-related genes were additionally enriched in a set of genes differentially expressed in early infancy brain tissue (Fig. S6).

Genes prioritized for hippocampus shape revealed a complete overlap with genes associated with its fissure, but only a partial overlap with its total volume (< 20%), reinforcing the validity and utility of our shape analysis (Fig. 5D).

### Associations with Polygenic Risk Scores

To investigate associations between brain-related traits and the LBS of individual brain structures, we extracted polygenic risk scores (PRS) from UKB for Alzheimer’s disease (AD), bipolar disorder (BD), ischemic stroke (ISS), multiple sclerosis (MS), Parkinson’s disease (PD), and schizophrenia (SCZ). Due to results of our gene-set enrichment analyses (see above), we also included the PRS for alcohol use disorder (ALC) as available in the PGScatalog. 10 of the pairwise partial correlations (i.e. adjusted for covariates, see Methods) between the PRSs were significant (Bonferroni-corrected p < 0.05), most prominently between SCZ and BD (r = 0.37, p<1E-300), ALC and SCZ (r = 0.12, p = 1.5E-60), and ALC and BD (r = 0.09, p = 1.7E-38) (Table S9-S11).

Following Sha et. al (*45*), we performed canonical correlation analysis (CCA) between the LBS of each of the 22 brain structures and each of those PRSs (see Methods). Here, the CCA determined a linear combination of the eigenvalues of an LBS (i.e. the canonical variable) which correlates maximally with the PRS across all examined individuals. We found 31 significant correlations after Benjamini-Hochberg (*46*) correction (p<0.05) within each PRS, 11 of which also stayed significant after additional Bonferroni correction for 6 independent PRSs (two-level correction), and 6 survived an overall Bonferroni correction for 22*6 tests (Fig. 6). As a balance between false discoveries and sensitivity, we chose this two-level correction to further investigate associations between PRSs and brain structures. Significant correlations ranged from 0.065 to 0.080 (mean 0.070). While we found several highly significant correlations with various brain structures for ISS and SCZ, the polygenetic risk for BD and PD did not correlate with any brain shape at all. This is particularly notable since the PRS of BD and SCZ had substantial correlation (see above). The PRSs of AD, ALC, and MS indicated only suggestive correlations with LBSs.

**Fig. 6.**
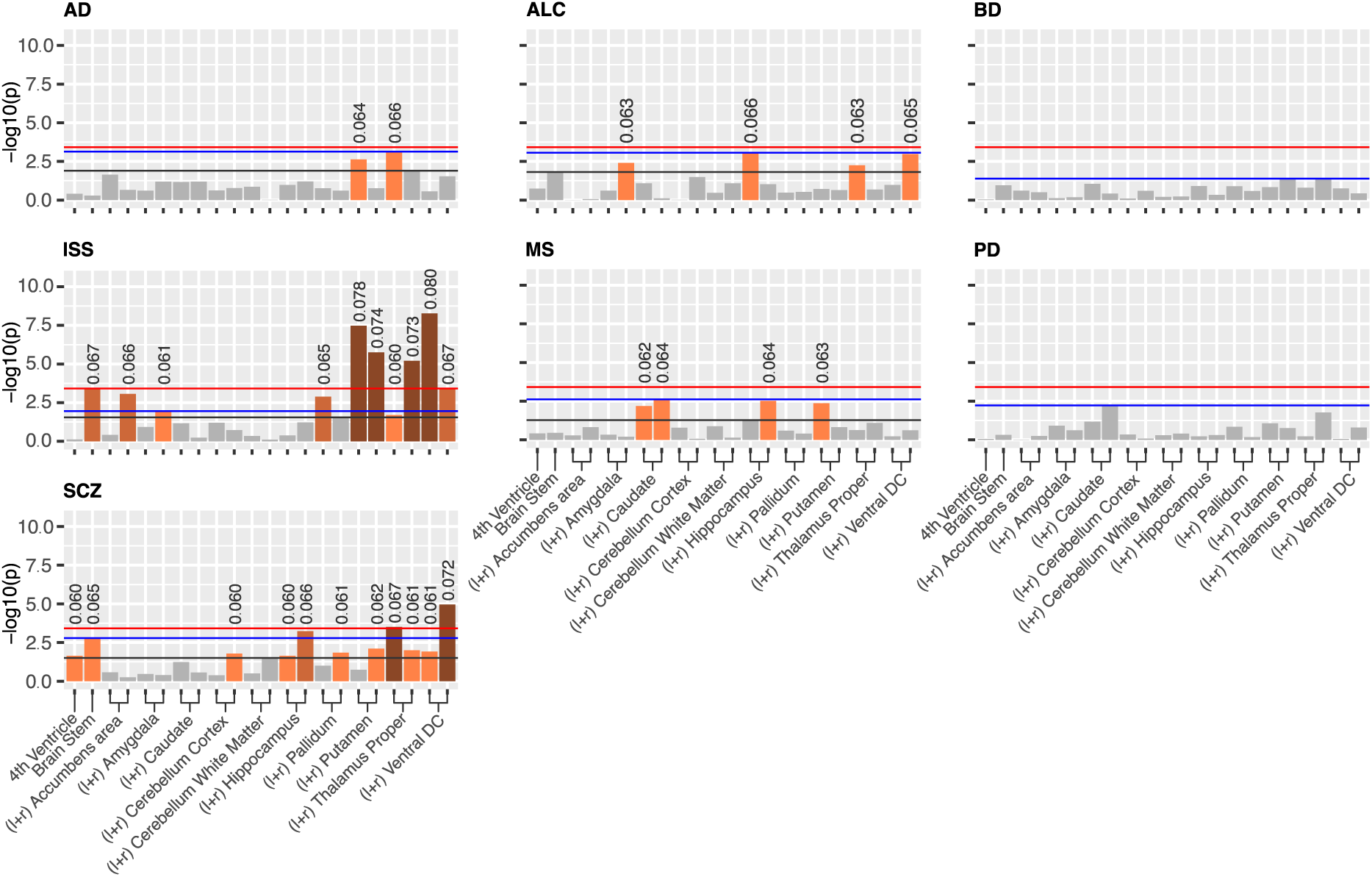
P-values of CCA Results of Different PRS with Various Brain Structures. Bar charts show the −*log*_10_ scaled p-values of the correlations between polygenic risk scores and LBS of different brain structures at different significance levels: Bonferroni-corrected for 22 brain structures and 6 independent PRS (red line), FDR-corrected within each PRS (black line), FDR-corrected within each PRS + Bonferroni-corrected for 6 independent PRS (blue line). Above significant p-values, the correlation value is stated. Bilateral structures are labeled once and the one in the left hemisphere is placed more left.

To interpret these findings at the level of the eigenvalues, we calculated the mean loadings in each CCA and counted the number of negative and positive values. We noticed that in some results it was possible to assess a common direction of effects. Among all significant results, the right ventral diencephalon (DC) revealed the strongest (i.e. largest absolute) mean loading of - 0.295 with 96% of all eigenvalues having a negative correlation with the canonical variable and, in case of a common direction, by that with the PRS of SCZ. This finding suggested that a person may have a higher genetic risk for schizophrenia if the right ventral DC is shaped in such a way that its eigenvalues (frequencies) are lower. This, for example, can be the case for a smoother surface or a less curved shape. A similar deduction in case of brain stem suggested that a less smooth or more curved shape shows a higher risk for ISS (mean loading 0.186 with ∼92% loadings positive).

Although most correlations remained above the significance threshold, clustering of the mean loadings showed similar tendencies for both hemispheres suggesting no asymmetric behavior (Fig. S7).

### Heritability

We used SCORE (*47*) to estimate the univariate heritability of each eigenvalue which utilizes the individual genotype data similar to the analysis by Ge et al. (*6*). We also calculated the multidimensional heritability (*6*) (see Methods) for each brain structure (Table 2, Table S12).

**Table 2.** Heritability Estimates of Different Brain Structures. Heritability estimates are presented for single brain structures (ℎ^2^) and for combined regions (ℎ^2^ combined) as well as their standard errors (SE and SE combined). Here, numbers in bold are significant after Bonferroni correction for 22 brain structures. For comparison, the estimates from Ge et al. are listed in the last two columns. Italic numbers are nominally significant and bold ones are significant after false discovery rate correction in (*6*).

| Brain Structure | $h^2$ | SE | $h^2$<br>combined | SE<br>combined | $h^2$<br>Ge2016 | SE<br>Ge2016 |
| --- | --- | --- | --- | --- | --- | --- |
| 4th Ventricle | 0.169 | 3.0E-3 | 0.169 | 3.0E-03 | 0.005 | 0.208 |
| Left Accumbens area | 0.134 | 2.8E-3 | 0.116 | 1.9E-3 | 0.237 | 0.135 |
| Right Accumbens area | 0.095 | 2.4E-3 |  |  |  |  |
| Left Amygdala | 0.105 | 2.5E-3 | 0.118 | 1.8E-3 | - | - |
| Right Amygdala | 0.134 | 2.8E-3 |  |  |  |  |
| Left Caudate | 0.296 | 1.9E-3 | 0.285 | 1.4E-3 | 0.499 | 0.188 |
| Right Caudate | 0.273 | 2.0E-3 |  |  |  |  |
| Brain Stem | 0.282 | 5.4E-3 | 0.198 | 1.5E-3 | 0.452 | 0.192 |
| Left Cerebellum Cortex | 0.186 | 2.9E-3 |  |  |  |  |
| Right Cerebellum Cortex | 0.194 | 2.8E-3 |  |  |  |  |
| Left Cerebellum White Matter | 0.195 | 2.6E-3 |  |  |  |  |
| Right Cerebellum White Matter | 0.148 | 2.6E-3 |  |  |  |  |
| Left Hippocampus | 0.187 | 2.6E-3 | 0.199 | 1.9E-3 | 0.347 | 0.169 |
| Right Hippocampus | 0.210 | 2.7E-3 |  |  |  |  |
| Left Pallidum | 0.124 | 2.9E-3 | 0.122 | 2.0E-3 | 0.061 | 0.117 |
| Right Pallidum | 0.120 | 2.8E-3 |  |  |  |  |
| Left Putamen | 0.158 | 2.7E-3 | 0.183 | 1.9E-3 | 0.413 | 0.148 |
| Right Putamen | 0.210 | 2.8E-3 |  |  |  |  |
| Left Thalamus Proper | 0.134 | 2.7E-3 | 0.132 | 1.9E-3 | 0.086 | 0.143 |
| Right Thalamus Proper | 0.130 | 2.7E-3 |  |  |  |  |
| Left Ventral DC | 0.122 | 2.7E-3 | 0.146 | 2.1E-3 | - | - |
| Right Ventral DC | 0.169 | 3.2E-3 |  |  |  |  |

Overall, the multidimensional heritability of the LBS was significant for all examined brain structures (Bonferroni corrected for 22 structures; range ℎ^2^: [9.5%, 29.6%], mean 17.2%). The highest heritability was found for the caudate (left: 29.6% and right: 27.3%) as well as for the brain stem (28.2%). We further computed the multidimensional LBS heritability for combined structures to compare them with the results from Ge et al. (Table 2, Fig. S8) and found a positive linear correlation (Pearson’s r = 0.735, p = 0.038). However, Spearman’s rho did not reach statistical significance (rho = 0.714, p = 0.058), suggesting a nonmonotonic relationship. Excluding the 4^th^ Ventricle, which may be an outlier in Ge et al. due to its large standard error and very low heritability, both results affirm a significant correlation (r = 0.844, p = 0.017 and rho = 0.786, p = 0.048). We further conducted a Wald test for different heritability in each combined brain structure with 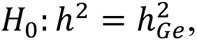 which could not be rejected at a 0.05 significance threshold for any of the structures (see Methods and Table S13), implying that our heritability estimates do not differ significantly from those of Ge et al.

### Replication

On the 20% replication sample (N=4961), we performed a replication analysis of the 80 independent SNPs (see Methods). For each brain structure, we only investigated those variants of the 80 SNPs which had also shown Bonferroni-corrected genome-wide significance in the respective discovery GWAS. We replicated 51.4% (76/148) of all significant associations of these 80 SNPs at a 0.05 significance level after FDR correction and 60.8% (90/148) at nominal significance level (Table S14). Hence, using a similar replication sample size, our replication rate was at least 33% larger than in van der Meer et al. (*2*) who introduced MOSTest in their study on brain volumes. To assess what can be expected as replication rate, we split our discovery set into 4 sets that equaled the replication set in size and sex and age distribution and checked the “replication rate” in these sets. We obtained rates between 66.2% and 80.4% at FDR-controlled significance and between 68.2% and 82.4% at nominal significance (Table S15). In view of the fact that replication rates of GWASs usually are much lower than expected by power analysis (48% observed vs 91% expected), even after correction for winners’ curse (48% vs 54%) (*48*), this simple simulation showed that our actual replication rate in a set of independent individuals was in keeping with the size of that set.

## Discussion

Examining the complex relationship among brain morphology, genetics and neurological disorders is crucial for unravelling the underlying causes and enhancing our understanding of the anatomy of the human brain and diseases. The Laplace-Beltrami operator provides a useful spectral representation for describing shape characteristics. Adequately describing curvatures and being an informative topological fingerprint of the shape (*19*), the Laplace-Beltrami spectrum (LBS) captures more information compared to global shape quantities such as surface area or volume. Specifically, it captures local shape effects but still represents the whole form of an object since the respective eigenmodes are defined everywhere on the structure. Furthermore, the shape is described by a relatively small number of values, which simplifies data handling, and, in contrast to other dimension reduction methods like principal component analysis, spatial interpretability is still given. Also, it does not depend on error-prone interindividual image registration which is required by other methods. Using the LBS thus allowed us to specifically address the variation in shape independent of volume or surface area and to disentangle the genetics of morphology beyond those global properties. While the eigenmodes and their potential to explain shape-associated biological mechanisms have been recently explored (*23*), the eigenvalues were previously found to have considerable heritability in the case of subcortical brain structures and cerebellum (*6*). However, neither the genetic architecture at SNP level of the shape of brain structures nor possible relations to disease genetics have been investigated yet. Using state-of-the-art GWAS and the LBS (a set of eigenvalues) as a multidimensional phenotype, we have dissected the genetics of the shape of 22 brain structures in 19,862 White-British individuals from the UK Biobank.

Overall, we identified 148 significant SNP associations that were independent in their respective GWAS. Since some SNPs were significantly associated with more than one brain structure, this resulted in 80 unique significant SNPs being independent across the 22 GWASs. Most of them, particularly the most significant ones, were detected in association with structures in the posterior fossa (brain stem and cerebellum). In contrast, we found no significant signal for the LBS of the amygdala. Due to the simple almond-like shape, crude measures like volume and surface area might already capture amygdala’s shape well enough (Supplements, Table S16). This could possibly explain the lack of genetic signal in our study since we controlled for both volume and surface area.

Of those 80 SNPs, 37 have previously been reported as genome-wide significant in a large GWAS on subcortical volumes (*2*), while 4 were identified in a large GWAS meta-analysis on total brain volume (*1*). This overlap with volume-associated SNPs suggests pleiotropic effects of the respective gene loci, affecting both volume and shape characteristics independent of volume, while it is unlikely that pure volume associations were picked since we controlled for volume. Indeed, our hippocampus LBS GWAS, for instance, replicated 100% of all loci previously associated with hippocampal fissure size (*49*), which obviously influences hippocampal shape, while accounting for only 20% of the loci associated with total hippocampal volume (Fig. 5D).

31 of the 80 independent SNPs were not associated with any brain shape-related trait according to GWAS catalog, neither by themselves nor by any variant in LD with them (*r*2 ≥ 0.6), and 18 were not listed at all (Table S1). This shows a substantial degree of novelty, that is, undiscovered genetic effects specific to shape of brain structures and supports the notion of SNPs affecting the development of form independent of growth.

Using a 15 times larger sample size, we were able to substantially refine the LBS SNP-heritability estimates of subcortical brain structures, including cerebellum, previously provided by Ge et al. (*6*) (N=1320). While their estimates ranged from 0.005 to 0.5, some being non-significant due to large standard errors of 0.12 to 0.21, our estimates were between 0.1 and 0.3, all being significant with much lower standard errors (≤0.005). In fact, statistical tests implied that our heritability estimates did not differ significantly from those of Ge et al. This was likely due to the much larger standard errors of their estimates but may also be a consequence of the younger age span in their cohort (18-35 years). Overall, we achieved results with higher statistical significance corresponding to our larger sample size and also likely due to our additional control for total brain volume, the respective local surface area, and MRI scan quality. The refined estimates were approximately equal to the SNP-heritability of volumetric traits of subcortical structures which Hibar et al. (*3*) derived from similar sample sizes, or even larger as in the case of caudate, hippocampus and putamen, for which they derived a SNP-heritability of about 0.11 with 95%-CI < 0.16 each, while we found values of 0.29, 0.20, and 0.18 respectively.

Bilateral structures showed similar genetic architecture of LBS (Fig. 3), which could be expected due to their symmetry. Moreover, genetic architecture was also shared between structures, as suggested by their adjacent positions, e.g. in the case of the basal ganglia. The two noticeably frequently shared variants rs13107325 and rs12146713 on *SLC39A8* and *NUAK1*, respectively, seem to affect brain shape globally as they have recently been associated with the structural connectome (*40*). While *SLC39A8*, as a brain metal transporter, is involved in several brain and neurodevelopmental traits (see below), *NUAK1* is also known to regulate axon branching by controlling mitochondrial distribution (*50*). The genetic associations of cerebellum and brain stem LBS were more different. Brain stem stood out due to a high number of specific association signals. These included the exonic nonsynonymous (ExNS) variant rs1687225 in *VWA5B2*, the gene of von Willebrand factor A domain containing protein 5B2. This missense variant belongs to the 3% most deleterious variants (according to CADD score) and was also an independent brain cis-eQTL of *VWA5B2* according to GTEx (v.8). Proxy variants of rs1687225 have been reported as being associated with brain morphology (*2*), cortical thickness (*51*), and educational achievement (*52*).

An extended genomic risk locus on chromosome 17 with lead SNP rs568589031 was also rather specific for the brain stem. ExNS variant rs17651549 at this locus had the highest CADD score (26.8) of all variants in LD with any of the independent significant SNPs. It affects *MAPT* which was among the most significant signals in MAGMA gene-based analysis of the brain stem LBS. However, this locus contains multiple protein-coding genes with several brain cis-eQTLs affecting them and other genes in the region (Table S5). The analysis of this large locus is highly challenging. The locus has previously been mapped to brain morphology (*2*) and several neurodegenerative diseases such as progressive supranuclear palsy, corticobasal degeneration, frontotemporal dementia, Parkinson’s disease, and Alzheimer’s disease (AD) (*42*, *43*, *53–62*). Moreover, variations in cortical morphology have recently been linked to *APOE* ε4 and *MAPT* in young healthy adults (*63*). There is also evidence of brain stem deformations in early stages of AD manifesting as variations in the midbrain (*64*), decreased locus coeruleus (part of the brain stem) volume preceding neuronal loss (*65*), and neurofibrillary tangle-related neurodegeneration in the brain stem potentially causing neuropsychiatric symptoms (*66*). While patients with diagnosed neurodegeneration were excluded from our study, the possibility remains that preclinical stages might potentially mediate the genetic relation between shape and the *MAPT* locus. However, neither gene enrichment analysis (Fig. 4 and 5) nor Canonical Correlation Analysis (CCA, Fig. 6), as an enrichment analysis independent of candidate genes, did provide evidence for a significant correlation between the genetics or phenotype of brain stem LBS and the genetics, respectively the PRS, of neurodegeneration such as AD.

At this locus, significant ExNS SNPs with high CADD scores were also found in *SPPL2C*, *KANSL1* and *CRHR1*, which showed the strongest signal in the MAGMA analysis (Table 1, Fig. 4A). Loss-of-function variants of *KANSL1* are known for causing autosomal-dominant Koolen-de-Vries syndrome (MIM #610443) which is characterized by intellectual disability and structural brain abnormalities (*67*). *SPPL2C* is the gene of signal peptide peptidase-like 2C and has previously been highlighted in a GWAS on total brain volume due to a relatively large number of ExNS (*1*). However, we controlled for volume in our GWAS of brain shape. A recent study found that *CRHR1*, which encodes a receptor for the corticotropin-releasing hormone CRH, moderates brain volume differences, possibly through its stress response function, which in return mediates the relationship between urban environmental exposure and affective symptoms (*68*). Especially, in the context of *CRHR1*, lower brain volume was observed in relation to stronger affective symptoms and greater urban environment exposure. Variants of *CRHR1*, have further been associated with alcohol (*69*) and heavy alcohol consumption following stressful life (*70*). Even light alcohol consumption has recently been related to changes in brain structure (*71*). Indeed, genes associated with the LBS of several other brain structures were enriched in gene sets associated with alcohol consumption (Fig. 5A, Fig. S4-S5). Thus, alcohol consumption may be a potential mediator of their effect on brain shape. However, we would like to note that the enrichment was driven by one locus.

That locus on chromosome 4 with lead SNP rs13107325, an ExNS variant of *SLC39A8,* appeared not only to be significant in most brain structures (15 structures including the brain stem, albeit with subthreshold significance) but also revealed the overall strongest signal across all the GWASs in left cerebellum white matter. This locus, especially the lead SNP, has a pleiotropic effect. Besides alcohol consumption (*72*), it has been linked to schizophrenia (SCZ) (*34–36*), Crohn’s disease (*37*), blood pressure and cardiovascular disease risk (*38*), and several brain imaging phenotypes (*9*). The cerebellothalamic and cerebellar-basal ganglia connectivity dysfunction hypothesis in individuals with SCZ is supported by the significant and prominent occurrence of rs13107325 in precisely these regions (*73*, *74*). SCZ is known to be associated with abnormalities of subcortical brain structures (*75–77*). Clinical features of SCZ might be due to aberration of dendritic spine density (*78*). A knock-in mouse model of rs13107325 points to an increased risk of SCZ by regulating zinc transport and dendritic spine density and subtle effects on cortical development due to this variant (*79*). A recent gene-mapping study concluded strong evidence for pleiotropic genes associated with SCZ and brain structure with evidence of brain variation causing SCZ (*80*).

SCZ shows substantial genetic correlation (0.68) with bipolar disorder (BD) (*81*) which was also evident in the correlation (0.37) between their PRSs. However, BD seems to have less relation to brain morphology. Madre et al. (*82*) investigated cortical morphology in individuals with SCZ and BD and reported shared volume and thickness deficits while abnormalities of geometry and curvature were specific to SCZ. Also, Stauffer et al. reported the association of MRI metrics to be weaker with BD genetics than with SCZ genetics (*80*). Our results are in line with these findings as CCA revealed correlation of the LBS of several brain structures only with the SCZ-PRS while there was no correlation with BD-PRS (Fig. 6). This result suggests that LBS may therefore be sensitive to SCZ-specific brain shape variations and thus potentially helpful for the differential diagnosis of psychosis.

CCA also revealed highly significant links between the LBS of multiple brain structures and the PRS of ischemic stroke (ISS) in our healthy cohort (Fig. 6). In line with the high correlation between ISS-PRS and hypertension (HT) PRS, CCA produced a very similar result for the HT-PRS (Fig. S9). HT is associated with white matter hyperintensities and cognitive dysfunction in elderly patients and is a major risk factor for ISS (*83*). A recent study showed that morphological changes in the basal ganglia and thalamus already occurring in mid-life are associated with blood pressure and, therefore, might be better markers of early HT than volume aberrations (*84*). Our CCA results were especially significant in these brain regions, which in turn support the suggestion of using brain morphology or shape as an early biomarker of HT.

Limitations of our study include the lack of effect sizes (beta values) in our multivariate results, which impedes the use of conventional post-GWAS tools such as genetic correlation and Mendelian randomization analyses. Instead, we explored the relationship to genetics of other traits by performing CCA between LBSs and PRSs. This was limited by the predictive power of each PRS. Furthermore, we cannot establish causal relationship between genetic variation in brain morphology and a particular trait or disease. Restricting our study to healthy individuals may additionally have yielded lower effect sizes and potentially false negative correlations when performing disease-related analyses. A follow-up of our findings, therefore, provides avenues for future research.

In summary, we have provided the first delineation of the genetic architecture of brain shapes beyond global measurements such as volume and surface area by using the Laplace-Beltrami spectrum as a mathematical shape descriptor. In our multivariate GWAS on a large healthy cohort of the UK Biobank, we identified 80 unique independent SNPs with brain stem showing a distinctly high number of specific association signals which included the *MAPT* locus implicated in neurodegenerative diseases. The shapes of most brain structures were significantly associated with the pleiotropic variant rs13107325, a known risk factor for schizophrenia. We further identified significant correlations between brain shape and polygenic risk scores of schizophrenia, hypertension and ischemic stroke, suggesting the potential use of the LBS as an early disease biomarker. As such, the LBS expands the set of tools for investigating brain shapes, holding implications for early detection of a wide spectrum of traits and warrants further research in disease-specific cohorts.

## Materials and Methods

### UKB Data and Sample Filtering

Data was retrieved from the official source of the <u>UK Biobank</u> (application number 41655) and provided as a DataLad dataset, a research data management solution providing data versioning, data transport, and provenance capture (*85*). A total of 487,409 genotyped samples were available (v3 imputed on Haplotype Reference Consortium and UK10K haplotype data, aligned to the + strand of the reference and GRCh37 coordinates, released in 2018, data field 22828). We kept only self-reported White-British individuals for data homogeneity according to data field 21000, resulting in 430,560 individuals. After genetic quality control (QC, see below), there were 393,913 unrelated samples left. 30,080 of them had structural MRI data (T1) on which we calculated the LBS (see below) of 22 brain structures (Fig. 1) of 24,834 individuals without mental or behavioral disorders (ICD index F) or diseases of the nervous system (ICD index G) according to the International Statistical Classification of Diseases and Related Health Problems 10th revision (ICD-10).

We created age and sex-stratified discovery and replication samples by splitting the data into 2 subsets comprising 80% and 20% of the individuals, respectively. Excluding samples without total brain volume data (see below), our final data set for our discovery GWAS contained 19,862 individuals, 9435 male and 10,427 female, ranging in age between 46.0 and 81.7 years with a mean±SD of 64.3±7.4 years (female: 63.6±7.3, male: 65.0±7.5).

### Quality Control of Genetic Data

We conducted QC using whenever possible Plink2 (*86*) and Plink (v1.9) for some missing functionalities on all self-reported White-British individuals and their imputed genotype data, which was initially converted from BGEN to binary Plink2 format, and kept only single-nucleotide polymorphisms (SNPs). As recommended by UK Biobank (*87*), we removed variants with an imputation score less than 0.3, and following Mills et al. (*88*), we excluded all variants with a call rate less than 0.95, a minor allele frequency of less than 0.01 and a Hardy-Weinberg equilibrium exact test p-value below 1E-6. As recommended by Plink2, we used the mid-p adjustment to reduce the filter’s tendency to retain variants with missing data (*89*) and the *keep-fewhet* modifier. We also removed samples with a mismatch between self-reported and genetically inferred sex, with genotype missingness of more than 0.05, and all heterozygosity outliers (±3 SD). Moreover, using a kinship coefficient of 0.088 and the *king-cutoff* command, we randomly excluded one from each pair of individuals related to ≥ 2nd degree. Only autosomal SNPs were examined. In total 8,105,763 SNPs remained after QC. We calculated the first 10 principal components (PC) from them, which were included as covariates in the GWASs.

### Image Data Processing

First, we segmented anatomical structures from all available T1 MRI brain scans with FreeSurfer version 7.2.0 (*90–94*). Second, we created triangular meshes for all structures of interest. Finally, we computed compact shape representations for all structures, using the BrainPrint Python package (*21*) (see below). To account for the quality of the MRI scans, we computed the Euler number with FreeSurfer. This represents the total defect index and is a measurement of the number of holes in the calculated surface.

### Multidimensional Shape Descriptor

A shape, parametrized as a Riemannian manifold *M*, may be described by its intrinsic geometric information, which can be obtained by solving the Helmholtz equation on that manifold:

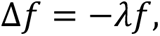

where Δ is the Laplace-Beltrami operator, a generalization of the Laplace operator in Euclidean space, and *f* a real-valued function, with *f* ∈ *C*^2^, defined on *M*. The solutions (*f*_*i*_, *λ*_*i*_) represent the spatial part of the wave equation with eigenfunctions *f*_*i*_ and eigenvalues *λ*_*i*_ where 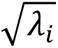 can be interpreted as the natural frequencies. The set of all eigenvalues is called the spectrum of an operator. The LBS or Shape-DNA (*13*) is accordingly defined as the beginning subsequence of the increasingly ordered spectrum of the Laplace-Beltrami operator solved on a Riemannian manifold:

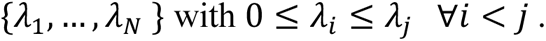

The LBS was computed with the BrainPrint package (*21*) based on the FreeSurfer output. For each individual, we calculated the first, i.e., the smallest, 50 eigenvalues of each brain structure as done in (*6*). Because the first eigenvalue is always zero, since each object is a closed surface without boundary, we used only the next 49 values in our analyses. Each eigenvalue *λ*_*i*,*m*_ was further normalized to volume *V*_*i*_ of each brain structure *i*. Volume and surface area were calculated by BrainPrint. Afterwards *λ*_*i*,*m*_ was divided by its position *m* in the ordered spectrum to balance out the higher eigenvalues which more likely represent noise (*6*, *13*, *21*):

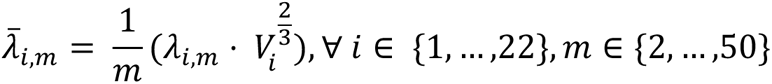

In the end, we analyzed 22 brain structures as provided by FreeSurfer: 4th ventricle, brain stem, accumbens area (left and right), amygdala (left and right), caudate (left and right), cerebellum cortex (left and right), cerebellum white matter (left and right), hippocampus (left and right), pallidum (left and right), putamen (left and right), thalamus proper (left and right), ventral diencephalon (DC) (left and right). By that, we handled the normalized and reweighted LBS 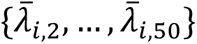 for each brain structure *i* as a 49-dimensional quantitative trait.

### Multivariate Genome-wide Association Analysis

For our multivariate GWAS, we used the MOSTest tool, a multivariate omnibus test, which handles big data efficiently and accounts for the correlation among the phenotypes to increase statistical power (*2*). Since the eigenvalues are not independent in general (Pearson correlation range: [0.285;0.998], mean±SD 0.93±0.06, Fig. S10-S12), we conducted, for each brain structure *i*, a multivariate GWAS on the 49 volume-normalized and scaled eigenvalues 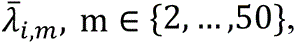 treating each 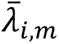 as a single quantitative phenotype. For that, we first calculated residuals of the multidimensional phenotype to control for potential covariates as recommended by MOSTest, i.e., we regressed each 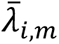 on age, age^2^, sex, first 10 genetic principal components (PCs), Euler number, surface area of each brain structure *i* and the total brain volume. The latter was calculated as the sum of the volume of ventricular cerebrospinal fluid (data field 25004) and the gray and white matter volume (data field 25010) in line with Jansen et al. (*1*). Normalizing the LBS to volume as described above and regressing out total brain volume and surface area of each brain structure assured that we indeed studied the shape and not linear size effects. We defined the residualized eigenvalues 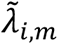 as the sum of the estimated intercept and the residuals of its linear regression. After adjusting for covariates, the range of absolute correlations between all 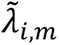 was quite high (Pearson correlation range: [0;0.978], mean±SD 0.65±0.18, Fig. S10-S12). We further inspected the relation of each 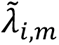 to volume and surface area by calculating their Pearson correlations. There was, as expected, no correlation with surface area and a low correlation with volume (average r = 0.28) (Fig. S13). The effect was small except for the eigenvalues of caudate, which showed a mean correlation of 0.50 (left) and 0.45 (right), likely relating to the long C-shaped form of that structure and by the fact that volume normalization (i.e. multiplication with 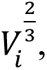 see above) excludes the pure size effects but not a real difference in shape. Including volume as additional covariate in the MOSTest analysis produced nevertheless similar results (Table S17).

All 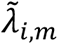 were then passed to MOSTest, which first performs a rank-based inverse-normal transformation (INT) to obtain normally distributed data. This is followed by a standard additive univariate GWAS for each 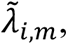 resulting in z-scores for each SNP and 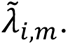 This procedure is repeated with once-permuted genotypes, preserving phenotype correlation. The MOSTest test statistic for a SNP is then calculated as the Mahalanobis norm of the non-permuted z-scores of that SNP and the correlation matrix *R* of the z-scores from once-permuted genotypes of all eigenvalues. The p-value of the multivariate test statistic is then calculated from a cumulative distribution function of a fitted gamma distribution, which eliminates the need for a multiple testing correction of 49 univariate GWASs and therefore allows to determine significance of SNPs with a p-value below the standard genome-wide threshold of 5E-8. We further confirmed that MOSTest controlled the type-I error sufficiently by plotting p-values from empirical distributions of the test statistic and from fitted gamma functions under the null hypothesis as calculated by permutating the phenotype-genotype assignments (Fig. S14-S35).

### Identification of Genomic Loci and Functional Annotations

For functional annotations, we used the web-based platform FUMA (v1.5.6) (*29*). We used default settings to determine genomic risk loci. First, independent significant SNPs were identified as the ones with p-value equal to or smaller than 2.27E-9 (Bonferroni correction for 22 brain structures 5E-8 / 22 = 2.27E-9) and linkage disequilibrium (LD) with other such SNPs in its vicinity of *r*2 < 0.6. These SNPs and those in LD with them (*r*2 ≥ 0.6) were defined as candidate SNPs and used in subsequent analyses. LD computation was based on the European population within the 1000 genomes reference panel (phase 3) (*95*). Those with *r*2 < 0.1 were classified as lead SNPs among the independent significant SNPs. Independent significant SNPs with *r*2 ≥ 0.1, or with a gap of less than 250 kb between their respective LD blocks (all SNPs in LD *r*2 ≥ 0.6 with them) were merged into one genomic risk locus. Therefore, a genomic risk locus can contain multiple independent significant SNPs.

Candidate SNPs were used for functional annotations. Positional annotation was performed with ANNOVAR (*30*). Further, SNPs were annotated with CADD scores, RegulomeDB scores, and 15-core chromatin states. The major histocompatibility (MHC) region was excluded from all annotations. For eQTL mapping, we checked several available databases containing relevant information, i.e., PsychENCODE eQTLs, ComminMind Consortium, BRAINEAC, and GTEx v8 Brain, and extracted all significant SNP-gene pairs with a False Discovery Rate (FDR, Benjamini-Hochberg procedure(*46*)) controlled p-value < 0.05 within the respective database.

### Gene-mapping and Gene-enrichment Analysis

For gene prioritization, we used PoPS (v 0.2) (*44*), a tool that assigns polygenic priority scores to each gene by fitting their MAGMA z-scores by trait-relevant gene features extracted from cell-type-specific gene expression, biological pathways and protein-protein interactions. We calculated these z-scores using MAGMA (v 1.10) (*41*) by first annotating SNPs to 18,383 protein-coding genes within a 0kb window and afterwards performing gene analysis using the SNP-wise mean model and the European population of 1000 Genomes as reference dataset for every brain structure. We then ran PoPS on our MAGMA scores using default settings and all available 57,543 gene features. Gene annotation and location files, as well as gene features, were taken from https://www.finucanelab.org/data.

Lead SNPs, as defined by FUMA (see above), were mapped to genes within a 500kb window up- and downstream. We first selected the four genes with the highest PoP score in each locus. Second, since it was shown that PoPS works best when combined with orthogonal methods like Nearest Gene, we mapped each lead SNP to the two closest genes. After assigning a rank to each gene by both methods and averaging the two ranks, we reranked the genes and finally prioritized all genes with average rank ≤ 2 for each independent significant SNP.

The prioritized genes were given as input to FUMAs Gene2Func tool. Genes of brain structures present in both hemispheres were joined in each analysis. We used all protein-coding genes of Ensembl v110. Hypergeometric tests analyzed the overrepresentation of mapped genes in precalculated gene sets followed by an FDR control (Benjamini-Hochberg, p < 0.05) within each category.

### Canonical Correlation Analysis with PRS

For the canonical correlation analysis (CCA), we extracted PRS of 6 brain shape-related traits and disorders and of one trait that stood out from the FUMA gene-set results. 6 PRS were taken from the UK Biobank which have exclusively been trained on external data sets: Alzheimer’s disease (data field 26206), bipolar disorder (data field 26214), ischemic stroke (data field 26248), multiple sclerosis (data field 26254), Parkinson’s disease (data field 26260) and schizophrenia (data field 26275). Further, we calculated the PRS for alcohol use disorder based on the score definition from the PGScatalog (*96*) (PGS002738 (*97*)). UK Biobank data was not used for training in this last score but for one GWAS in the discovery meta-analysis of the alcohol use disorder study. The data was available for 19850 samples. Each PRS was first linearly regressed on age, age^2^, sex, first 10 genetic principal components (PCs), and total brain volume. The residuals were added to the fitted intercept. Second, a rank-based INT was applied to those PRS. The partial correlation between the PRS was calculated as a Pearson correlation. We used the residualized eigenvalues 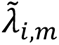 as they were used as input for MOSTest for each brain structure and applied a rank-based INT.

For each pair of PRS and brain structure, we conducted a CCA using the cca() function within the “yacca” R-package (version 1.4-2) (*98*). CCA finds a linear combination of all eigenvalues, a so-called canonical variable, which correlates maximally with the PRS. All resulting p-values of the correlations were adjusted for multiple testing using FDR. Afterwards, we applied an additional Bonferroni correction for 6 effective PRS, which we consider as the two-level correction. For comparison, we also calculated the Bonferroni correction for 22*6 tests. All adjustments were made to the 0.05 significance threshold.

Further, in each CCA, we investigated the loadings, i.e. the correlations of all eigenvalues with their canonical variable, which describe the direction and strength of the impact of each eigenvalue on the risk score. Out of those, we calculated the mean loading and the number of negative/positive loadings of all eigenvalues in each brain structure and PRS. If at least 90% of all loadings had the same direction, we deemed that as a common direction of effects. Hierarchical clusters were computed on the distances of mean loadings among brain structures and PRS, respectively, using the complete linkage method (Fig. S7).

### SNP-based Heritability

Ge et al. investigated the heritability of neuroanatomical shape using the LBS. For estimating its SNP-heritability, they proposed a multidimensional approach where they summed the univariate heritability 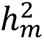 of the eigenvalues, each weighted by γ_*m*_, the relative size of its phenotypic variance:

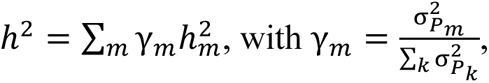

and 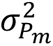 being the phenotypic variance of trait *m*. The univariate heritabilities of the 49 eigenvalues were derived using SCORE. For combined structures, we derived the multidimensional heritability from the union set of eigenvalues. When applying SCORE, we proceeded as with the GWAS described above, using volume-normalized and reweighted eigenvalues and the same set of covariates. The phenotypic variance was also computed using the residualized eigenvalues 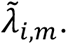 We further calculated the p-value of each heritability using the Wald test statistics, which is distributed as

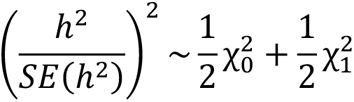

With 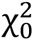 being the point mass at 0 and 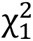 the chi-squared distribution with one degree of freedom since the null hypothesis *H*_0_: ℎ^2^ = 0 lies on the boundary of a constrained parameter space (ℎ^2^ ≥ 0) (*99*). The standard error (SE) of each multidimensional heritability was calculated using Bienayme’s identity for the variance of a sum and the univariate estimates from SCORE as follows:

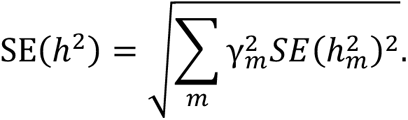

For testing the difference between heritability values in each combined brain structure, we utilized the Wald test statistics distributed as

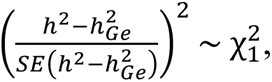

with

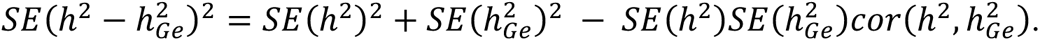

Since *SE*(*h*^2^) is small in comparison to 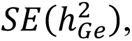 we can approximate 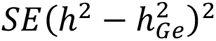 by just 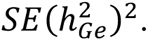

### Replication

Replication analysis was conducted on the 20% sample (see above). This encompassed 4963 individuals, of which 4961 (2605 female) had data of total brain volume available, ranging in age from 45.2 to 81.8 years (age mean±SD: 64.2±7.4, female: 63.6±7.2, male: 64.9±7.6). We used the same procedure and set of covariates to perform MOSTest analyses for each brain structure on this dataset and all SNPs passing QC. For each brain structure, we only investigated those variants of the 80 independent SNPs which had shown Bonferroni corrected genome-wide significance in the respective discovery GWAS. FDR correction was applied to these p-values for each brain structure separately.

## Data Availability

The primary data used in this study is from the UK Biobank. These data can be provided by UK Biobank pending scientific review and a completed material transfer agreement. Requests for the data should be submitted to the UK Biobank: www.ukbiobank.ac.uk. Specific UK Biobank data field codes are given in Materials and Methods. Other publicly available data sources and applications are cited in Materials and Methods. We will make our summary statistics available online within the GWAS catalog: https://ebi.ac.uk/gwas/. We will further publish the FUMA results within the FUMA database. Until then summary statistics and FUMA results can be obtained upon reasonable request to the authors.

## Acknowledgments

The authors gratefully acknowledge the computing time granted by the JARA Vergabegremium and provided on the JARA Partition part of the supercomputer JURECA and the joint lab Supercomputing and Modeling for the Human Brain at Forschungszentrum Jülich.

## Author contributions

Conceptualization: SP, SE, JW, KO, KP

Methodology: SP, KO, KP

Investigation: SP, KO, KP

Visualization: SP

Supervision: KO, KP

Writing—original draft: SP, KO, KP

Writing—review & editing: SP, FH, FR, SE, JW, KO, KP

## Data and materials availability

The primary data used in this study is from the UK Biobank. These data can be provided by UK Biobank pending scientific review and a completed material transfer agreement. Requests for the data should be submitted to the UK Biobank: www.ukbiobank.ac.uk. Specific UK Biobank data field codes are given in Materials and Methods. Other publicly available data sources and applications are cited in Materials and Methods. We will make our MOSTest summary statistics available online within the GWAS catalog: https://ebi.ac.uk/gwas/. We will further publish the FUMA results within the FUMA database.

This study used openly available software and codes, specifically BrainPrint (https://github.com/Deep-MI/BrainPrint), MOSTest (https://github.com/precimed/mostest), FUMA (https://fuma.ctglab.nl/), MAGMA (https://ctg.cncr.nl/software/magma, also implemented in FUMA), SCORE (https://github.com/sriramlab/SCORE), fastman (https://github.com/kaustubhad/fastman), ggseq3d (https://github.com/ggseg/ggseg3d/), circlize (https://github.com/jokergoo/circlize) and yacca (https://cran.r-project.org/web/packages/yacca/). We further used Affinity Designer for figure design.

