## Supplementary Materials for "Beyond Volume: Unraveling the Genetics of Human Brain Geometry"

Sabrina A. Primus *et al.*

**This PDF file includes:**

Supplementary Text
Figs. S1 to S36
Descriptions of Tables S1 to S22

**Other Supplementary Materials for this manuscript include the following:**

Tables S1 to S22

**Supplementary Text**

Replication in Amygdala

Neither for the left nor for the right amygdala did the GWAS on the LBS reveal genome-wide significant signals. We hypothesized that this is due to the simple shape of the amygdala, which is already represented by volume and surface area, both being controlled for in our study. Therefore, we reran MOSTest on non-normalized eigenvalues and did not include surface area as a covariate. We compared the results to the 12 loci that were reported by a recent study on whole amygdala volume (*100*) (Table S16). We replicated 4 (left amygdala) and 5 (right amygdala) loci at 0.05 significance level after FDR correction while our original GWAS on normalized and corrected LBS replicated only 2 loci each, thus supporting our hypothesis.

Robustness Analysis

Since MRI measurements of the brain stem can be influenced by the height of the individual, we conducted the MOSTest analysis again with height as additional covariate (Fig. S36 and Table S22). We replicated 35 of 37 independent significant SNPs at Bonferroni-corrected genome-wide significance level ($p<5E-8/22$) and 2 at genome-wide significance level ($p<5E-8$). All of them were significant after FDR-correction at 0.05 threshold.

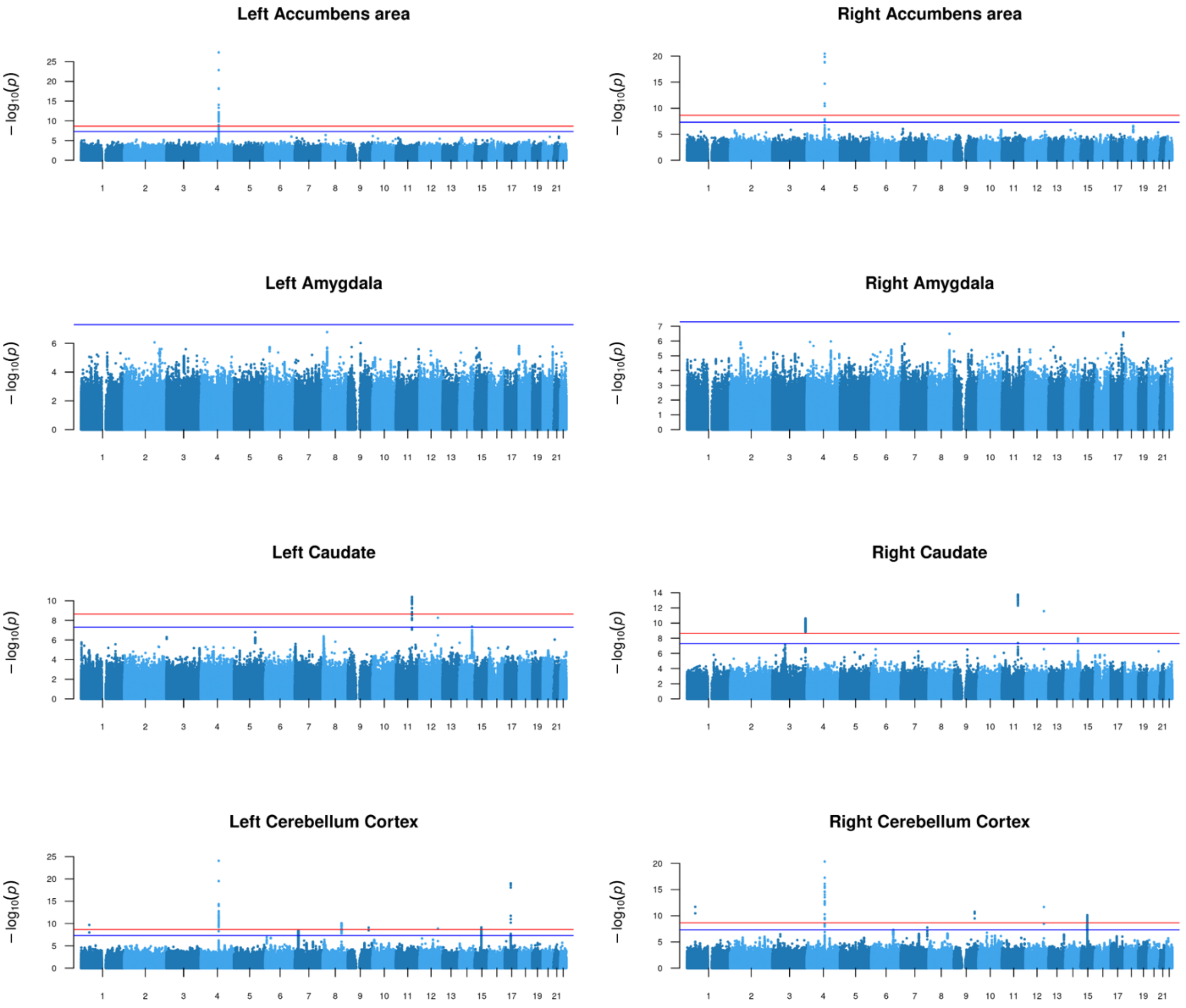


**Fig. S1. MOSTest Results.** This figure and Fig. S2 and S3 display the Manhattan plots for all brain structures, each showing the results of the LBS MOSTest analysis. The chromosomal location of the SNPs is indicated on the x-axis, and -log10 scaled p-values on the y-axis. Blue and red lines indicate standard genome-wide significance (p=5E-8) and genome-wide significance after Bonferroni correction for 22 brain structures (p=5E-8/22), respectively. The Manhattan plot for brain stem analysis is shown in the main text.

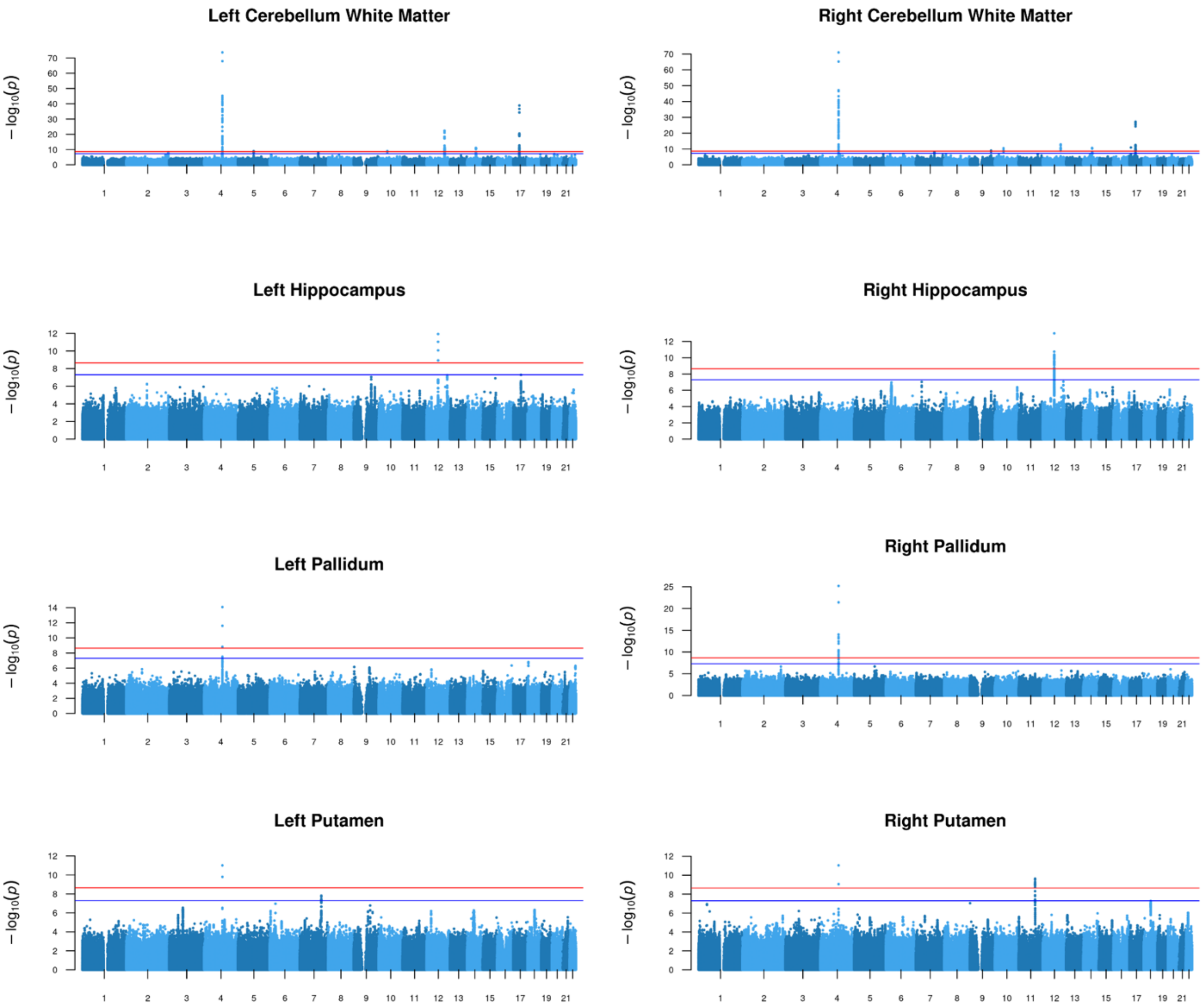


**Fig. S2. MOSTest Results.** See description of Fig. S1.

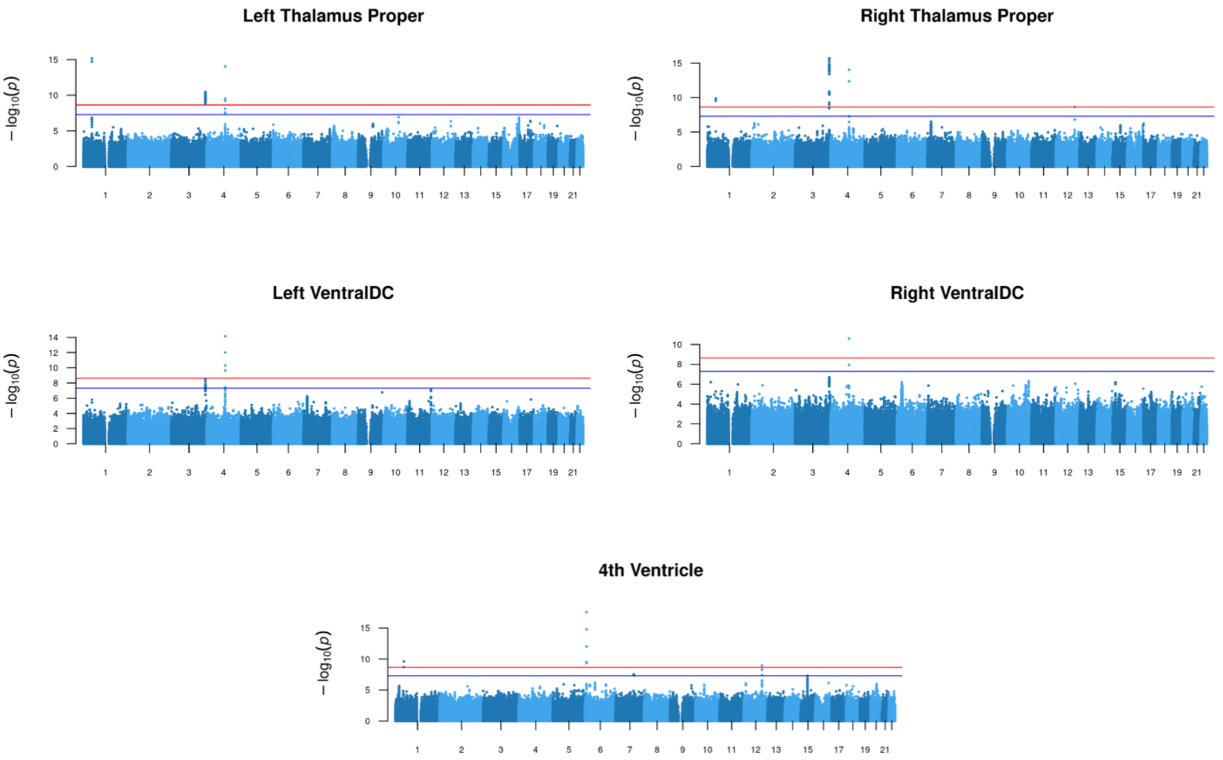


**Fig. S3. MOSTest Results.** See description of Fig. S1.

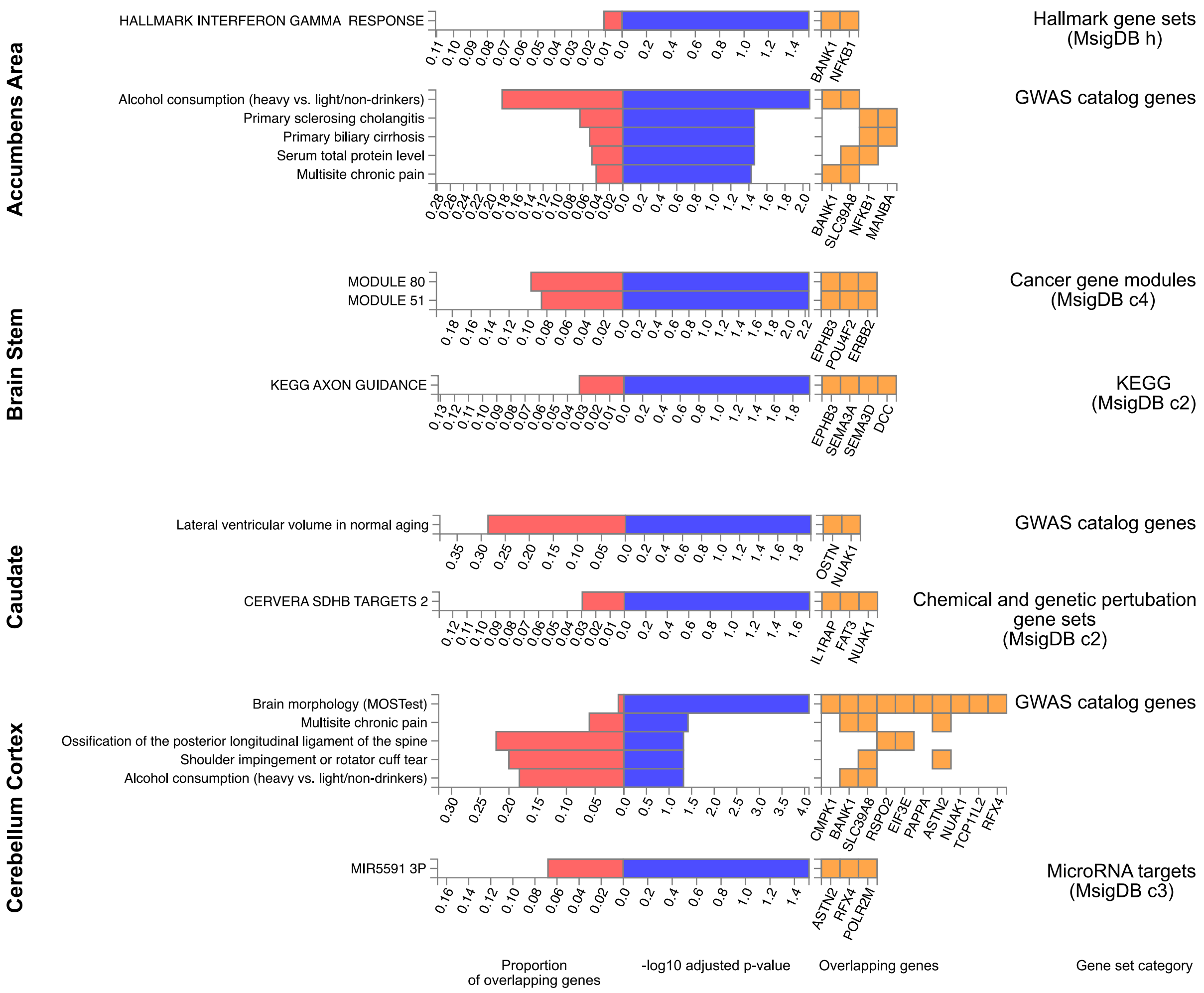


**Fig. S4. Gene Set Enrichment.** Significant gene set overlaps using FUMA's Gene2func method for prioritized genes as resulting from MOSTest on LBS of accumbens area, brain stem, caudate and cerebellum cortex which have not been shown in the main paper. P-values are FDR-adjusted for each gene set category. Enrichment results of further brain structures are shown in Fig. S5.

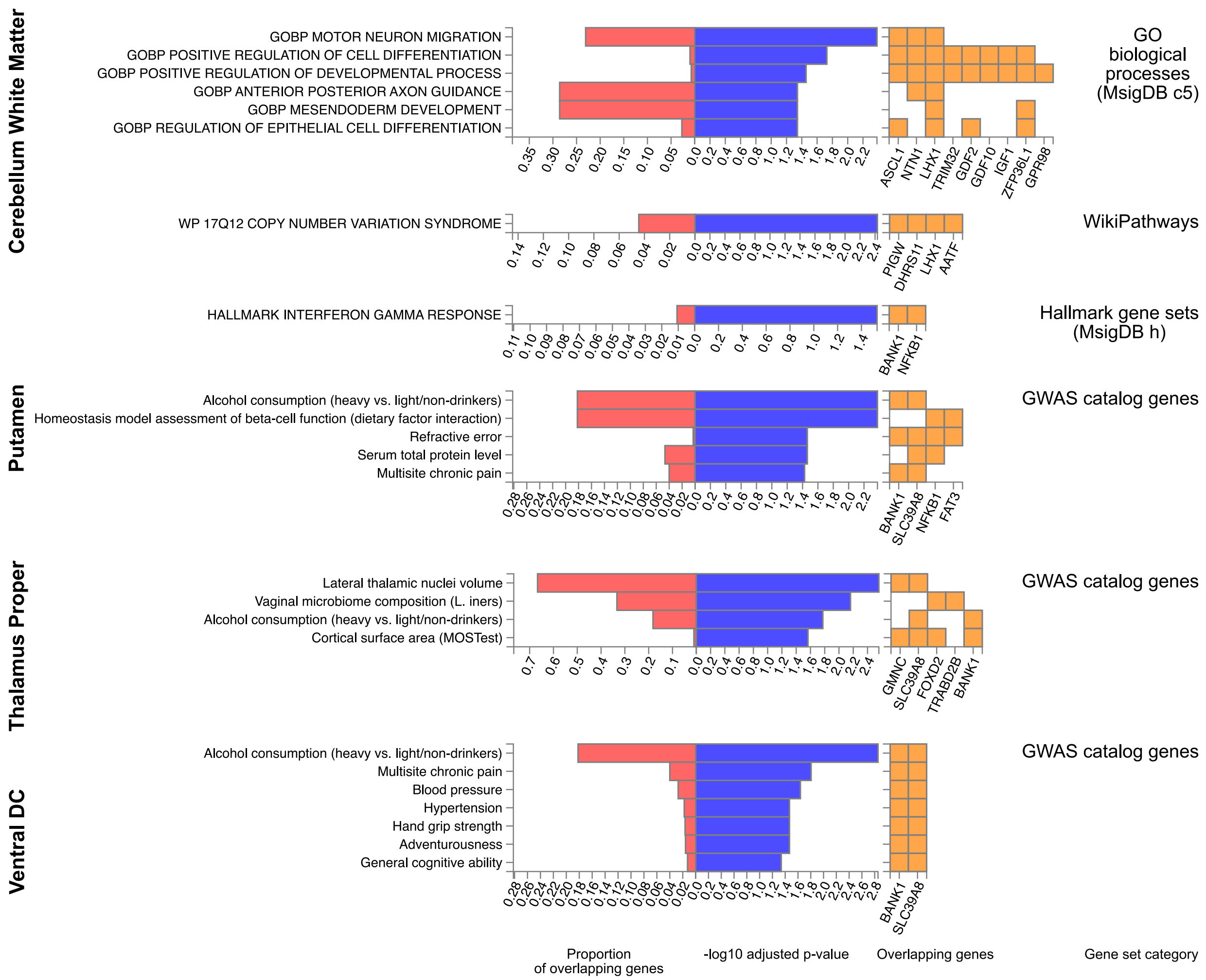


**Fig. S5. Gene Set Enrichment.** Significant gene set overlaps using FUMA's Gene2func method for prioritized genes as resulting from the MOSTest on LBS of cerebellum white matter, putamen, thalamus proper and ventral DC which have not been shown in the main paper. P-values are FDR-adjusted for each gene set category. Enrichment results of further brain structures are shown in Fig. S4.

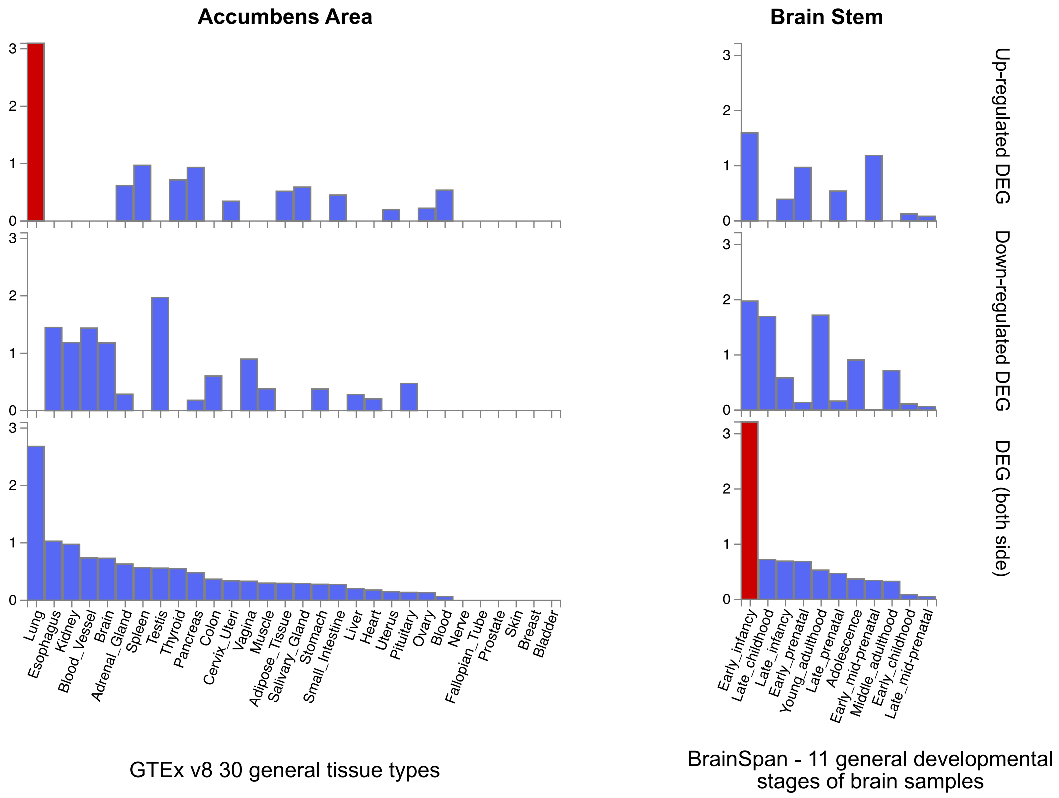


**Fig. S6. Tissue Specificity of Prioritized Genes.** Prioritized genes as resulting from MOSTest on LBS of accumbens area and brain stem were tested for enrichment in differentially expressed gene sets (x-axes) from GTEx v8 and BrainSpan using FUMA's Gene2func method. Y-axes indicate –log10(p-value). Significant enrichments (FDR-corrected) are indicated in red.

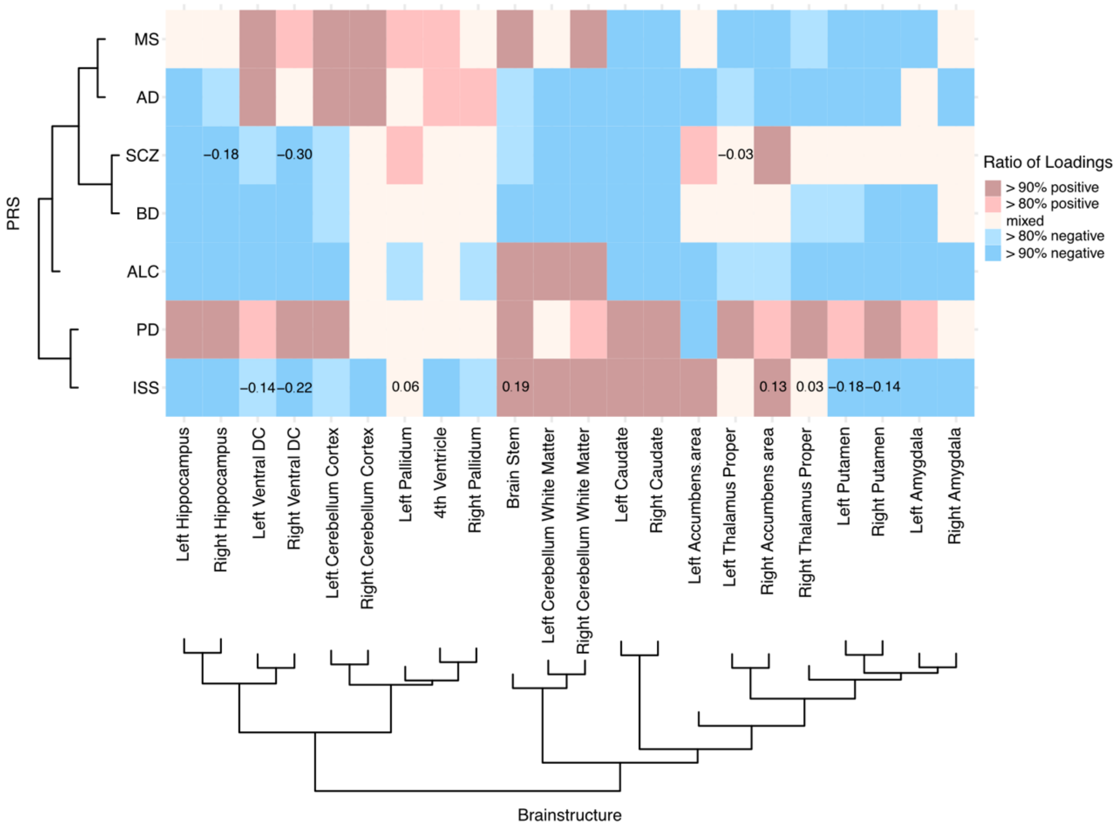


**Fig. S7. Clustered Mean Loadings of CCA between PRS and the LBS.** Polygenic risk scores (PRS) for Alzheimer's disease (AD), bipolar disorder (BD), ischemic stroke (ISS), multiple sclerosis (MS), Parkinson's disease (PD), schizophrenia (SCZ), and alcohol use disorder (ALC) (y-axis) were compared with the Laplace-Beltrami spectra (LBS) of brain structures (x-axis) by canonical correlation analysis (CCA). For each pair of disease and brain structure the mean of the eigenvalue loadings was calculated. These means were used for hierarchical clustering. The heatmap shows for each PRS-LBS pair the proportion of positive (or negative) loadings in the CCA, with red color indicating the predominance of positive loadings and blue color the predominance of negative loadings. For significant CCA results the mean loading is stated in the respective cell.

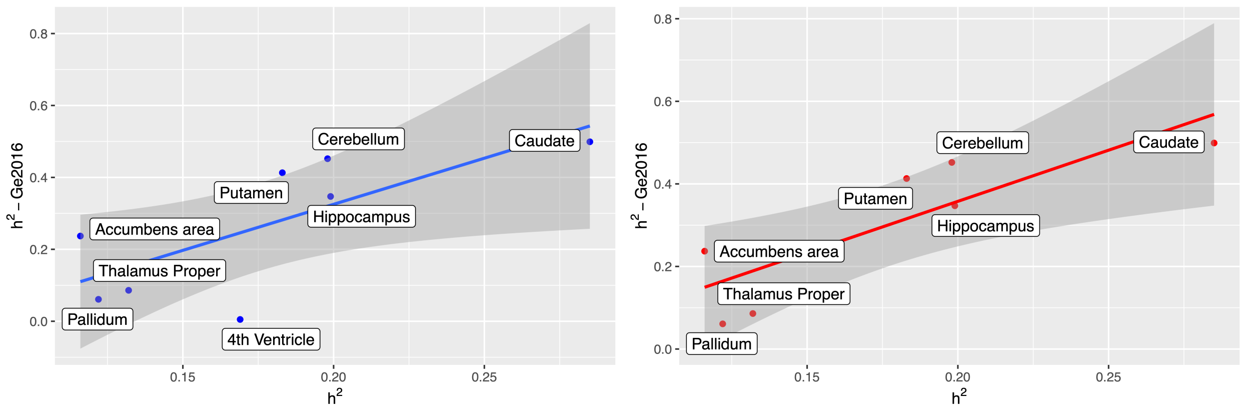


**Fig. S8. Comparison of heritability estimates.** The heritabilities of brain structures, as determined by (*6*), were compared to the heritabilities derived in the present study. For bilateral structures and combined regions, such as cerebellum, a combined heritability measure was calculated (see Methods of main text). The left plot shows all structures for which heritabilities are available from both studies and a linear fit with a 95% confidence interval. Since the result for the 4th ventricle might be an outlier in (*6*), the comparison was repeated without that structure as shown in the right plot.

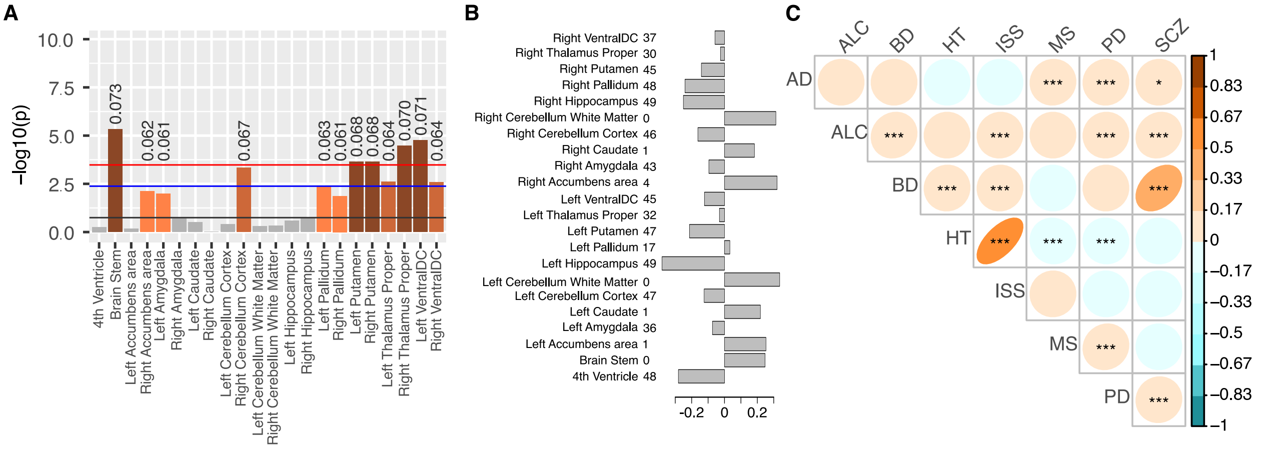


**Fig. S9. CCA Results for Hypertension PRS.** The polygenic risk score (PRS) of hypertension (HT, extracted from data field 26244 of UKB) had a Pearson correlation coefficient of 0.64 (-log(p)= 5210.3) with the PRS of ischemic stroke (ISS). Therefore, the results of the canonical correlation analysis (CCA) between HT PRS and the LBS of different brain structures were quite similar to those of the analogous CCA results for ISS PRS which are shown in the main text. **A:** Bar chart showing the p-values that resulted from CCA of HT PRS and brain structure LBS. Significance thresholds of increasing stringency are analogous to those in Fig. 6 of the main text, i.e., FDR-corrected for the set of brain structures (black line), FDR-corrected for the set of brain structures combined with Bonferroni-correction for 7 independent PRS (blue line), and Bonferroni-correction for 22 brain structures and 7 independent PRS (red line). **B**: Means of the loadings of the eigenvalues which were produced by CCA of HT PRS and the LBS of different brain structures. The number of negative loadings is stated after each structure's name. **C**: Coefficients of Pearson correlations among different disease PRS (abbreviations see Fig. S8) and their significances with p<0.05 indicated by "*", p <0.01 by "**", and Bonferroni-adjusted p<0.05/28 by "***".

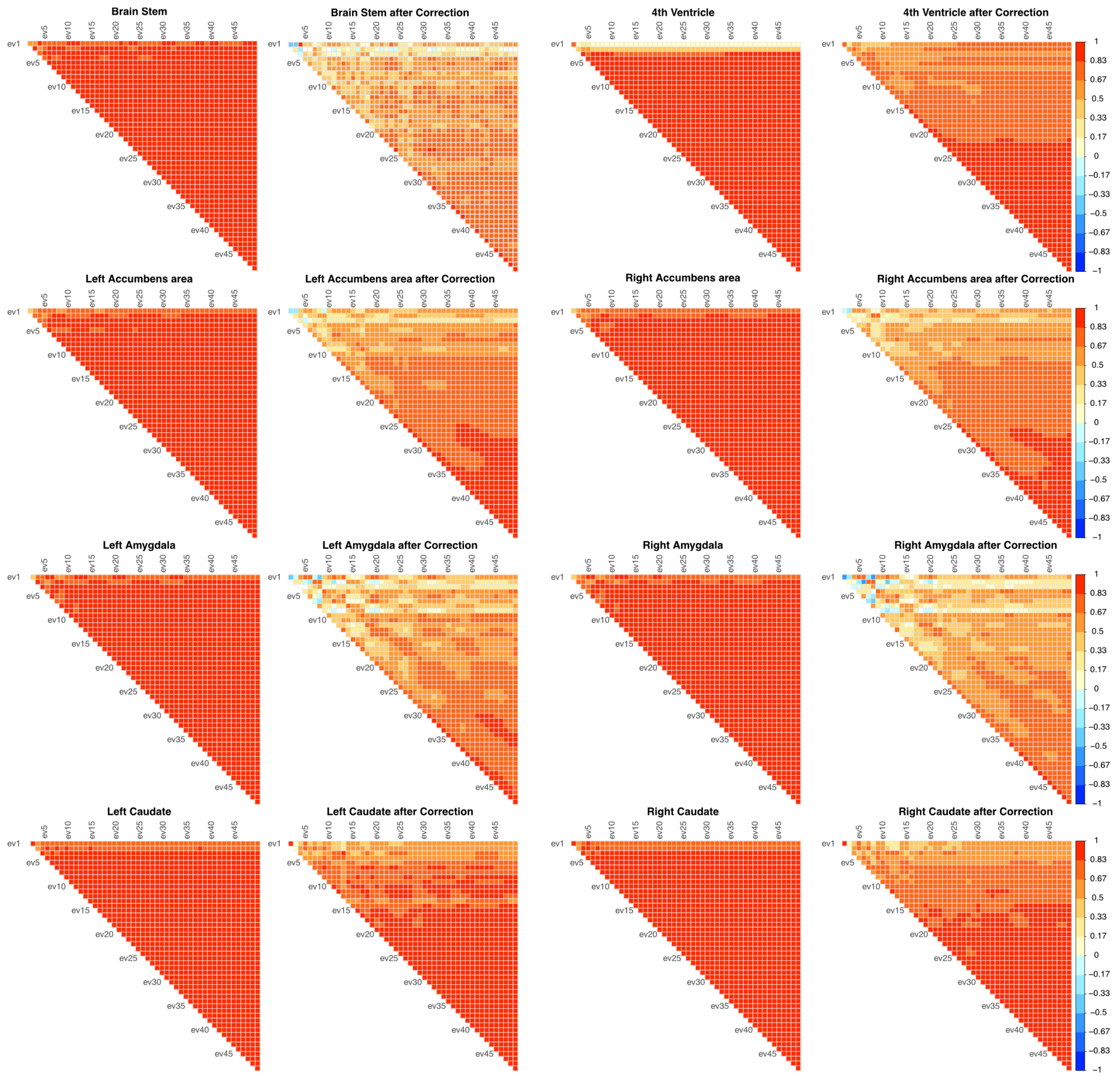


**Fig. S10. Phenotype Correlation.** This figure, as well as Fig. S11 and S12, show for each brain structure the Pearson correlation coefficients between eigenvalues before (raw) and after volume normalization and regressing on covariates. White cells indicate non-significant ($p \geq 0.05/49$) correlations.

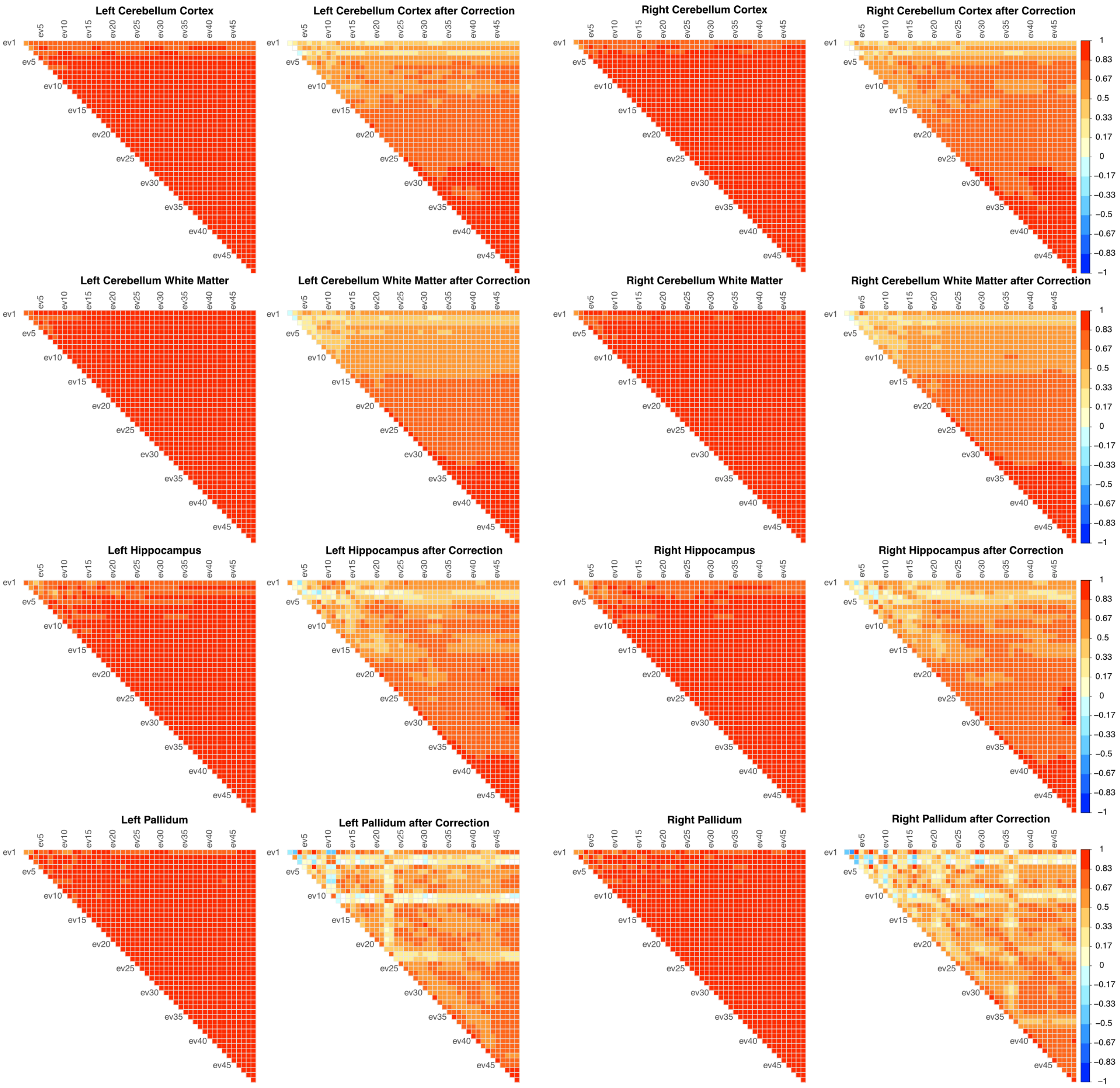


**Fig. S11. Phenotype Correlation.** See description of Fig. S10.

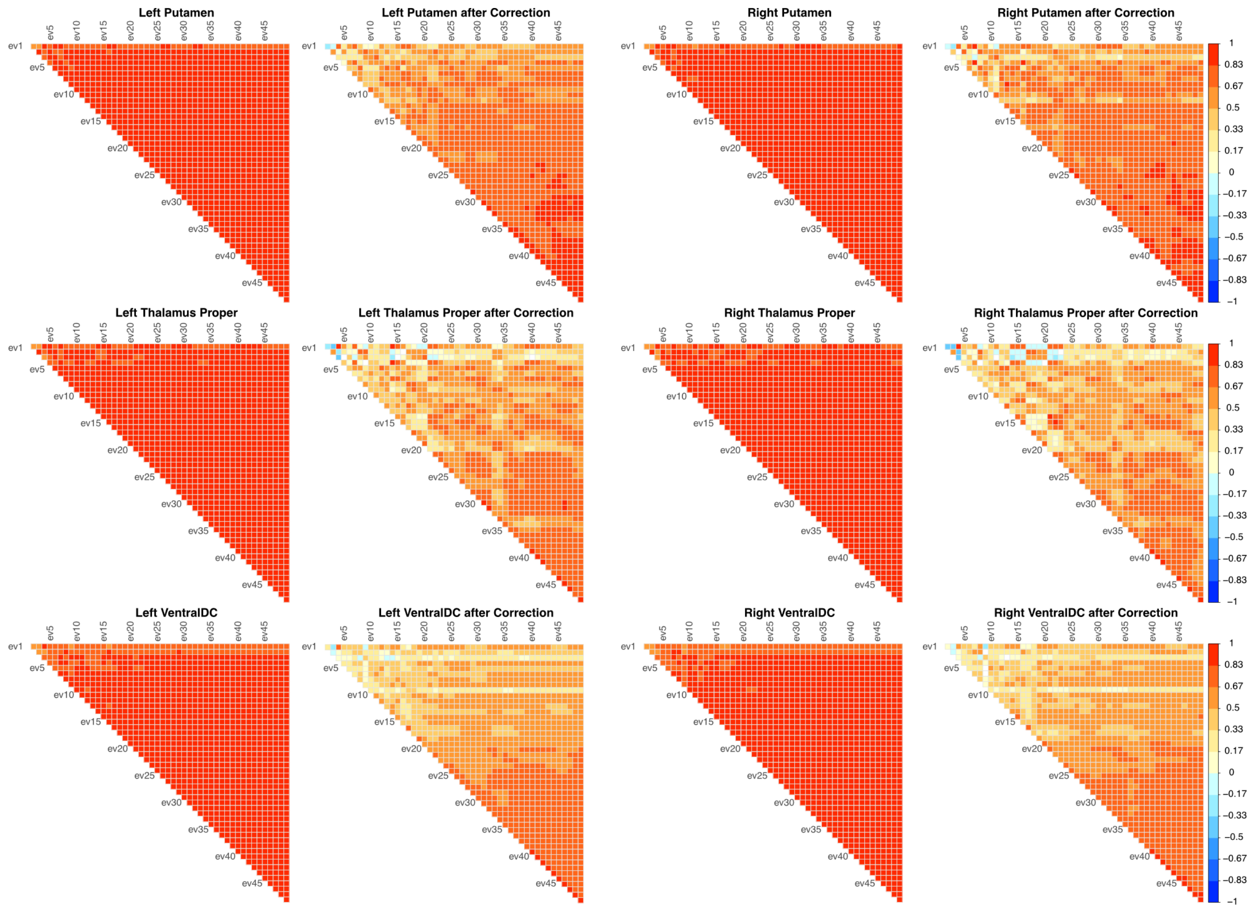


**Fig. S12. Phenotype Correlation.** See description of Fig. S10.

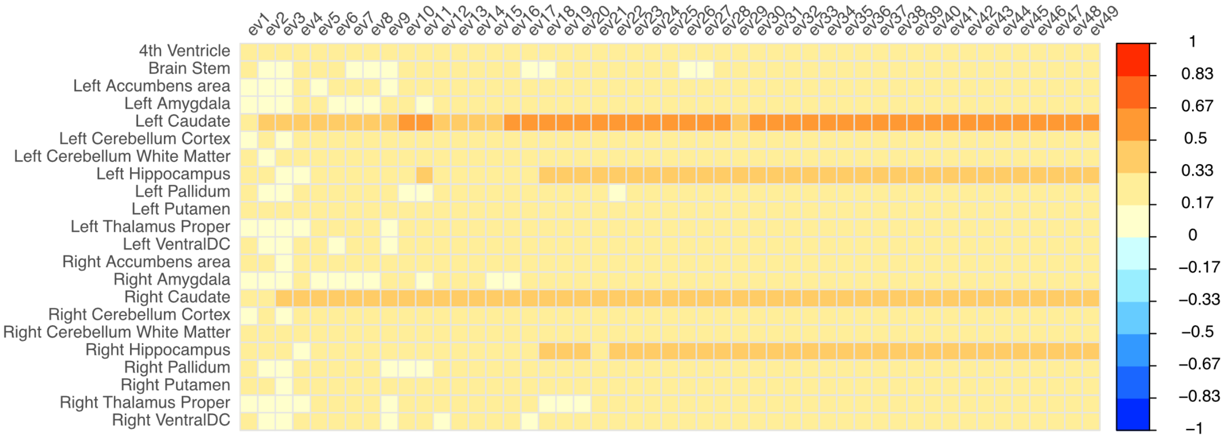


**Fig. S13. Correlation Between LBS and Volume.** Coefficients of Pearson correlation between the volumes of brain structures (y-axis) and their normalized and residualized Laplace-Beltrami eigenvalues.

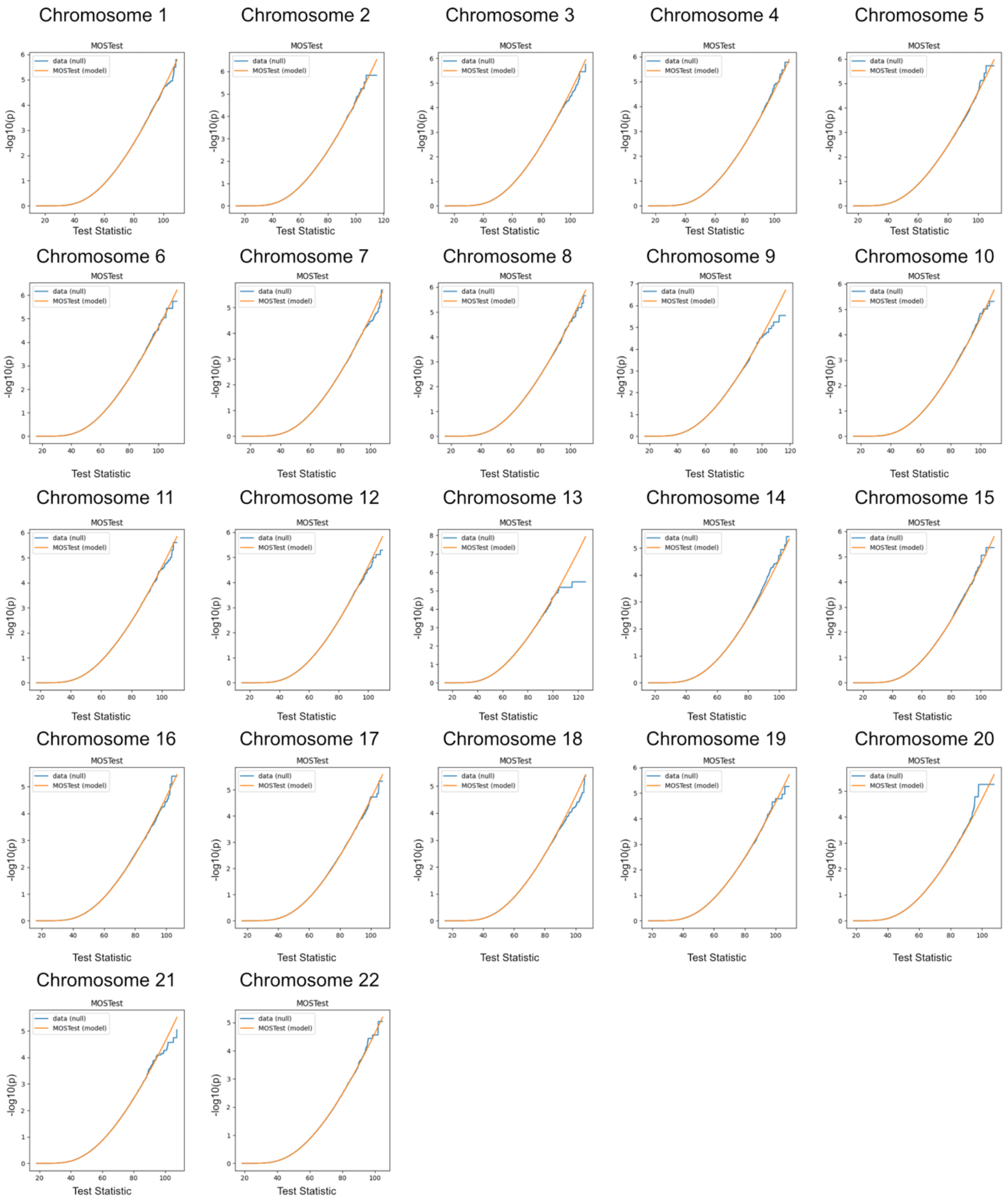


**Fig. S14. Chromosome-wise Comparison of Empirical and Analytical MOSTest Results Under the Null Hypothesis for Brain Stem.** Each subplot covers one chromosome and shows the value of MOSTest test statistic on the x-axis. The MOSTest model curve shows theoretical p-values calculated from a gamma function fitted to the test statistic (orange). The blue curve shows the empirical distribution of the test statistic. Coincidence indicates a uniform distribution of p-values under the null and a well-controlled type I error.

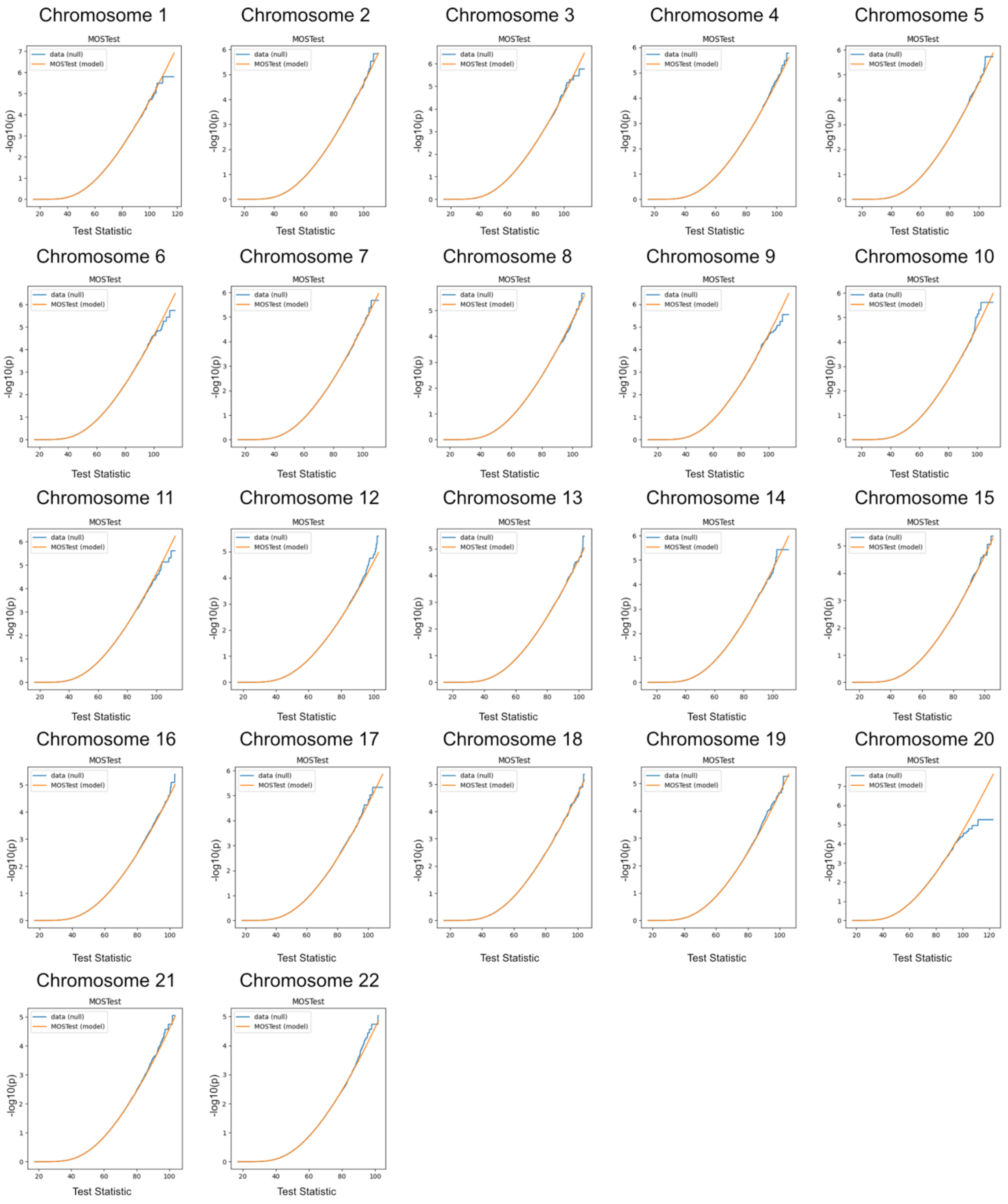


**Fig. S15. Chromosome-wise Comparison of Empirical and Analytical MOSTest Results Under the Null Hypothesis for left Accumbens Area.** See description of Fig. S14.

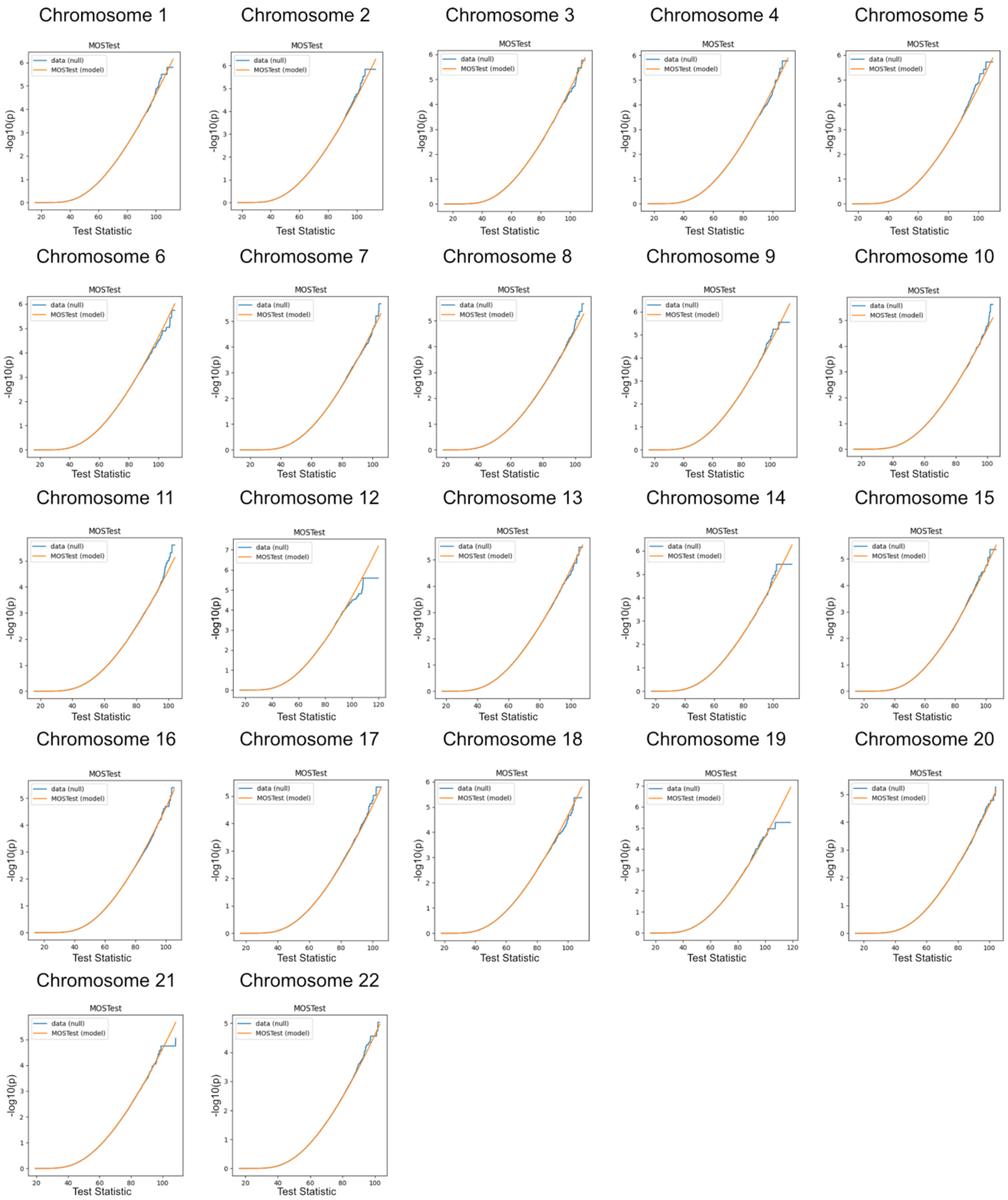


**Fig. S16. Chromosome-wise Comparison of Empirical and Analytical MOSTest Results Under the Null Hypothesis for right Accumbens Area.** See description of Fig. S14.

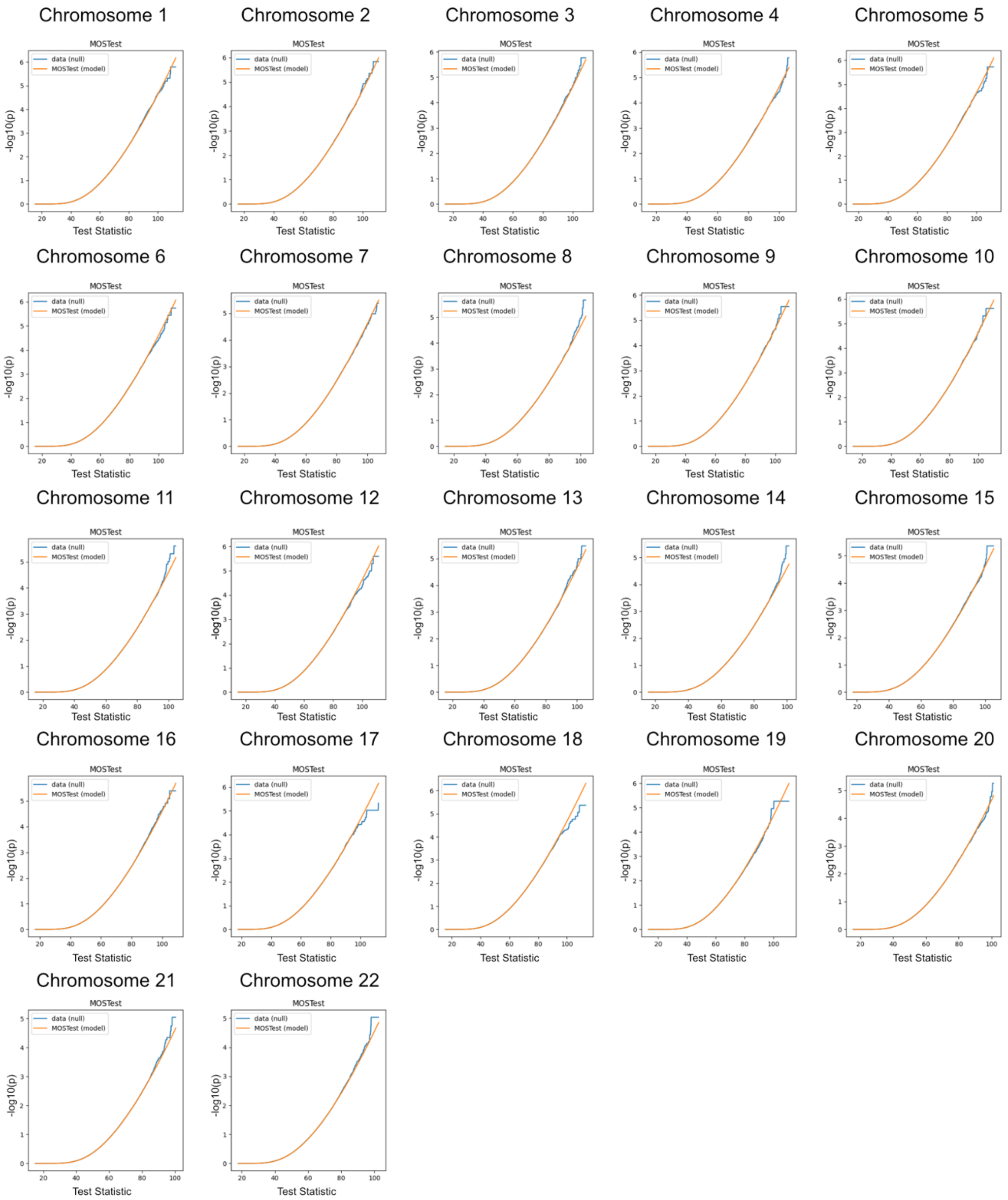


**Fig. S17. Chromosome-wise Comparison of Empirical and Analytical MOSTest Results Under the Null Hypothesis for left Amygdala.** See description of Fig. S14.

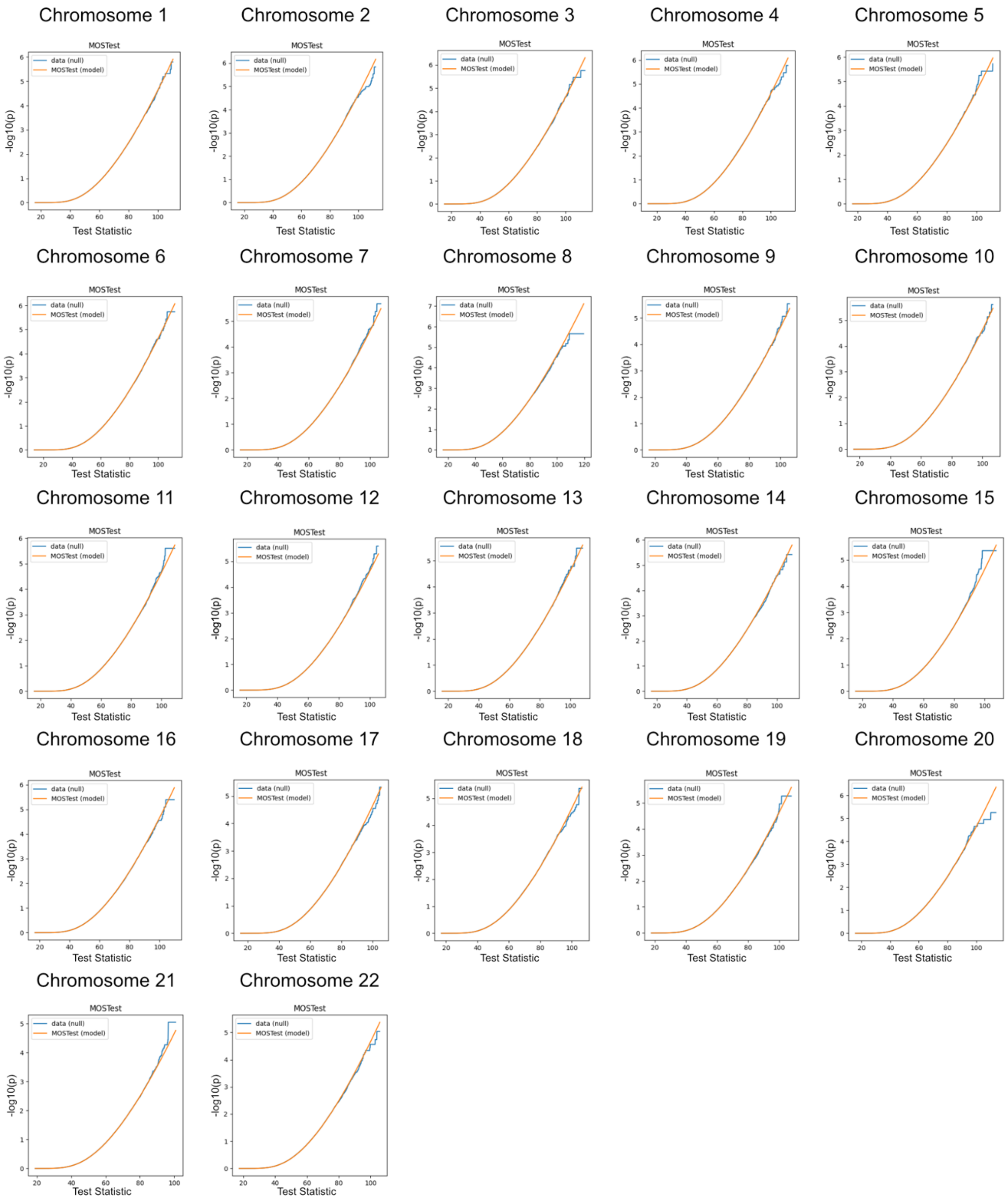


**Fig. S18. Chromosome-wise Comparison of Empirical and Analytical MOSTest Results Under the Null Hypothesis for right Amygdala.** See description of Fig. S14.

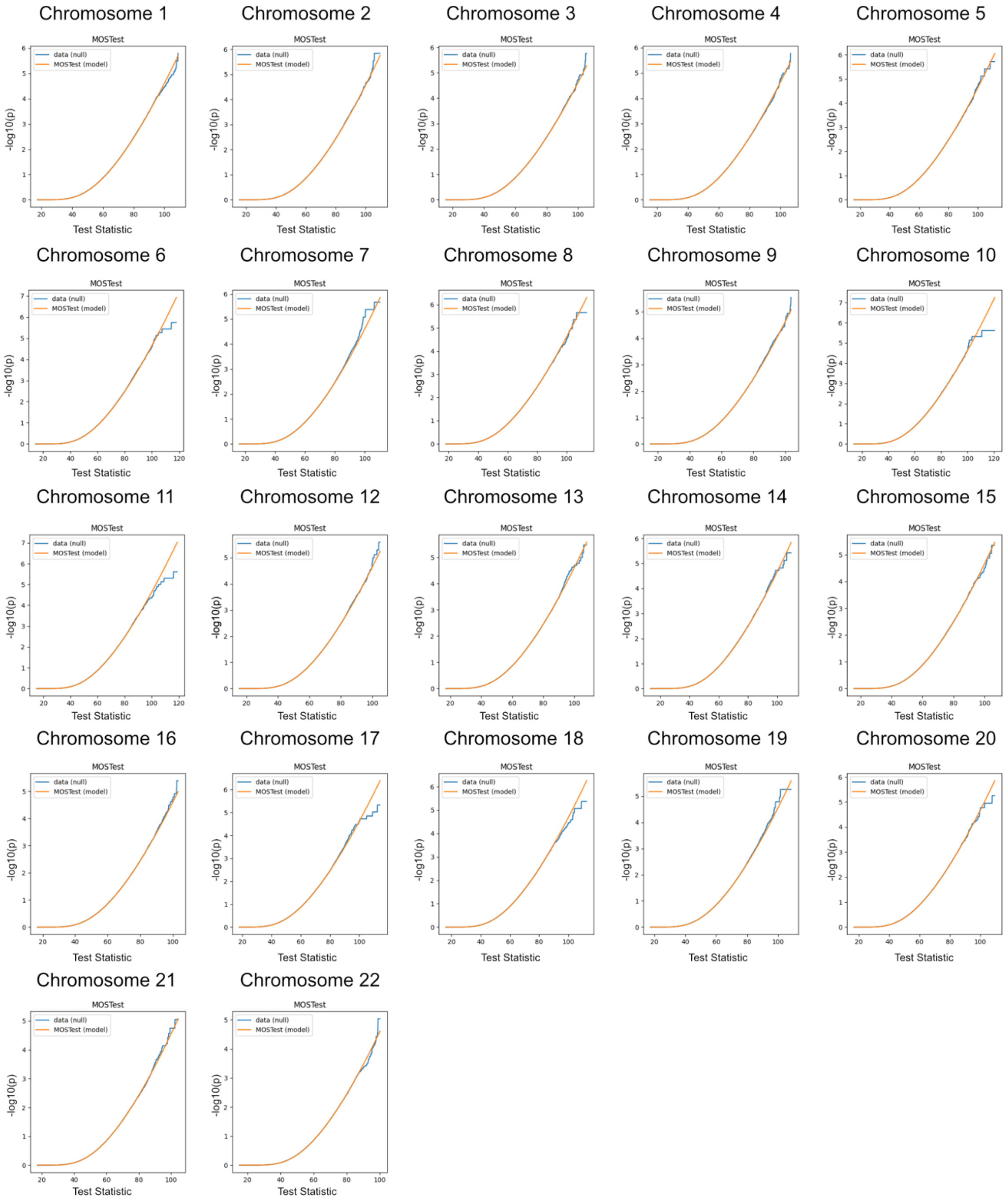


**Fig. S19. Chromosome-wise Comparison of Empirical and Analytical MOSTest Results Under the Null Hypothesis for left Caudate.** See description of Fig. S14.

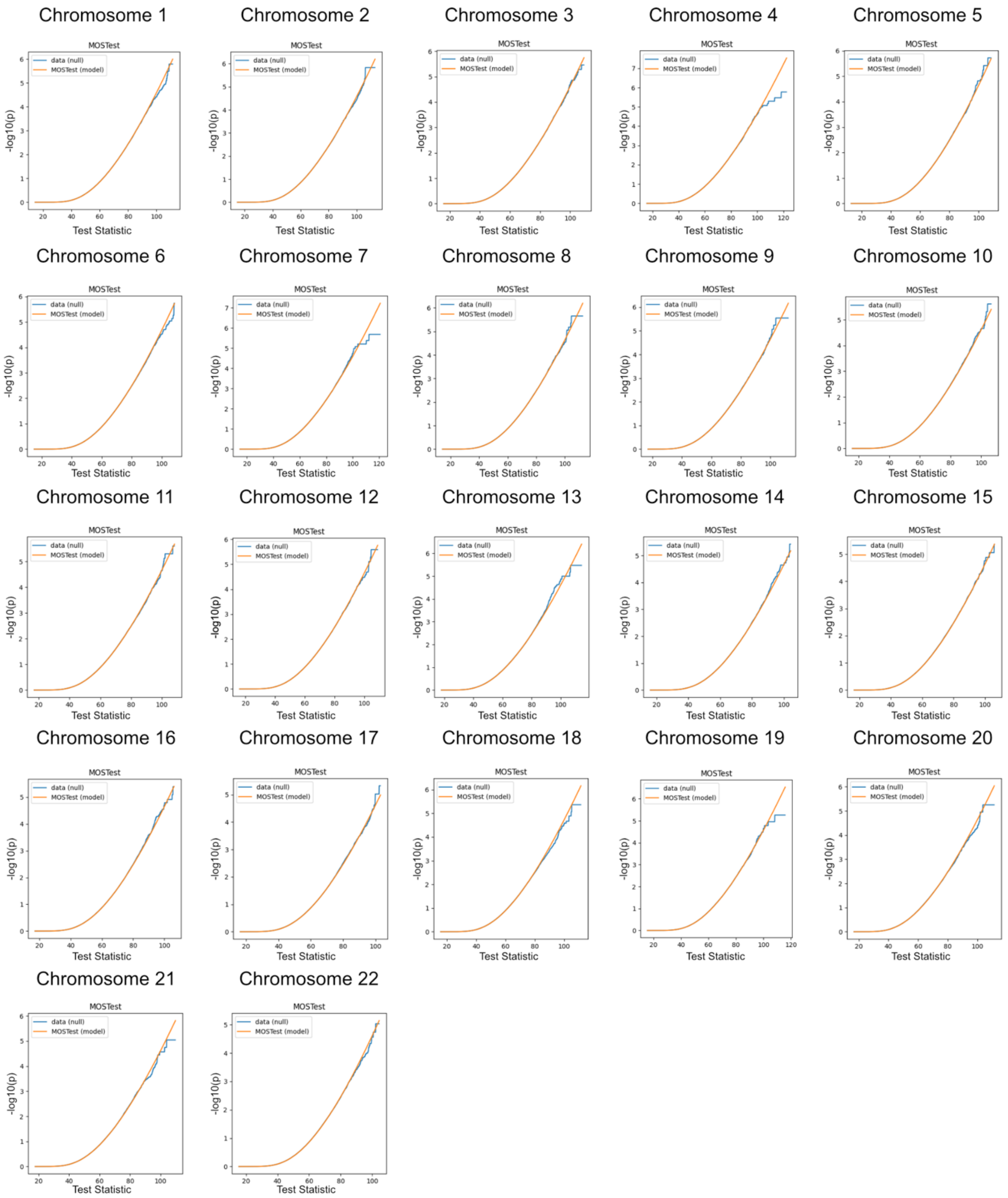


**Fig. S20. Chromosome-wise Comparison of Empirical and Analytical MOSTest Results Under the Null Hypothesis for right Caudate.** See description of Fig. S14.

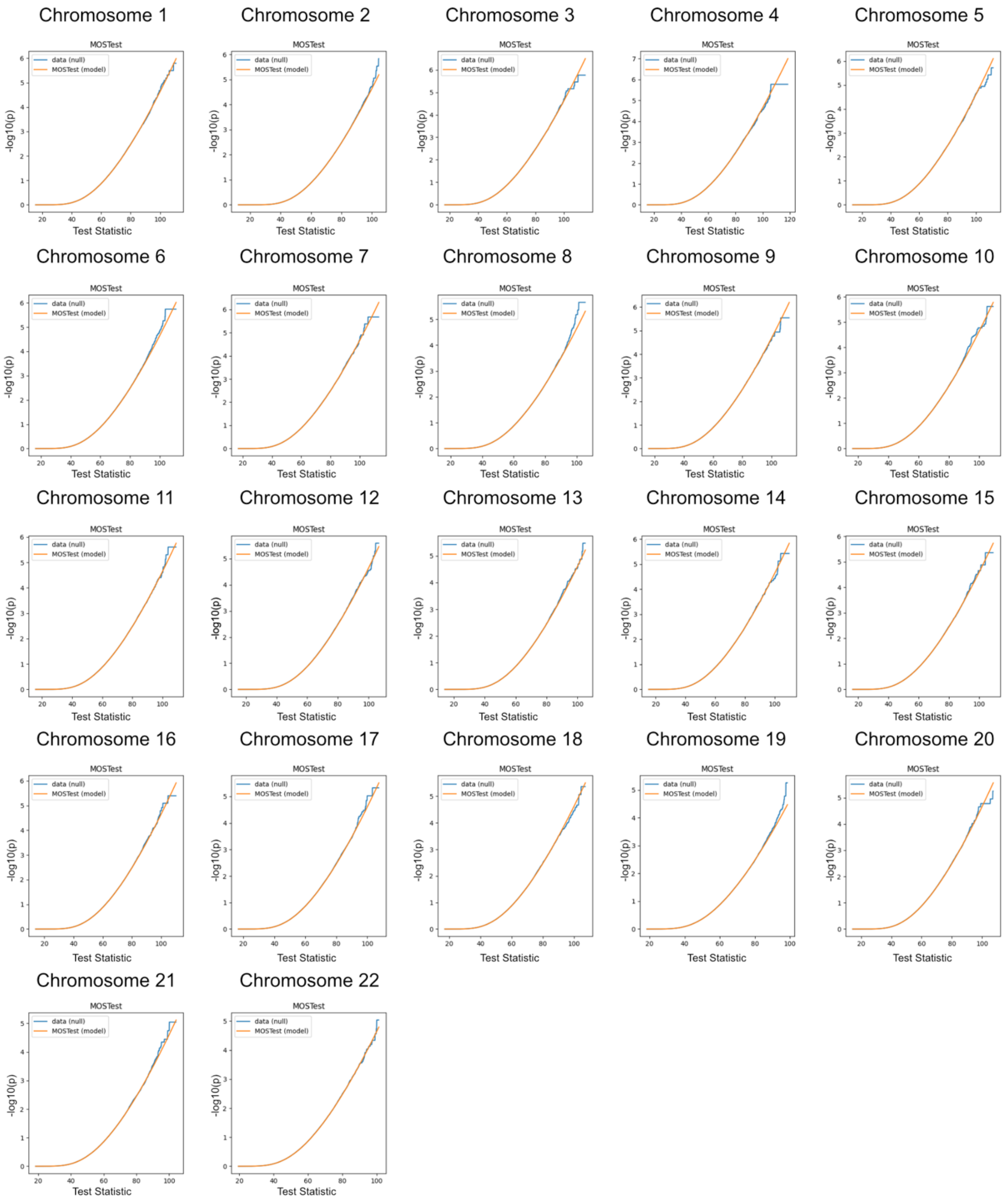


**Fig. S21. Chromosome-wise Comparison of Empirical and Analytical MOSTest Results Under the Null Hypothesis for left Cerebellum Cortex.** See description of Fig. S14.

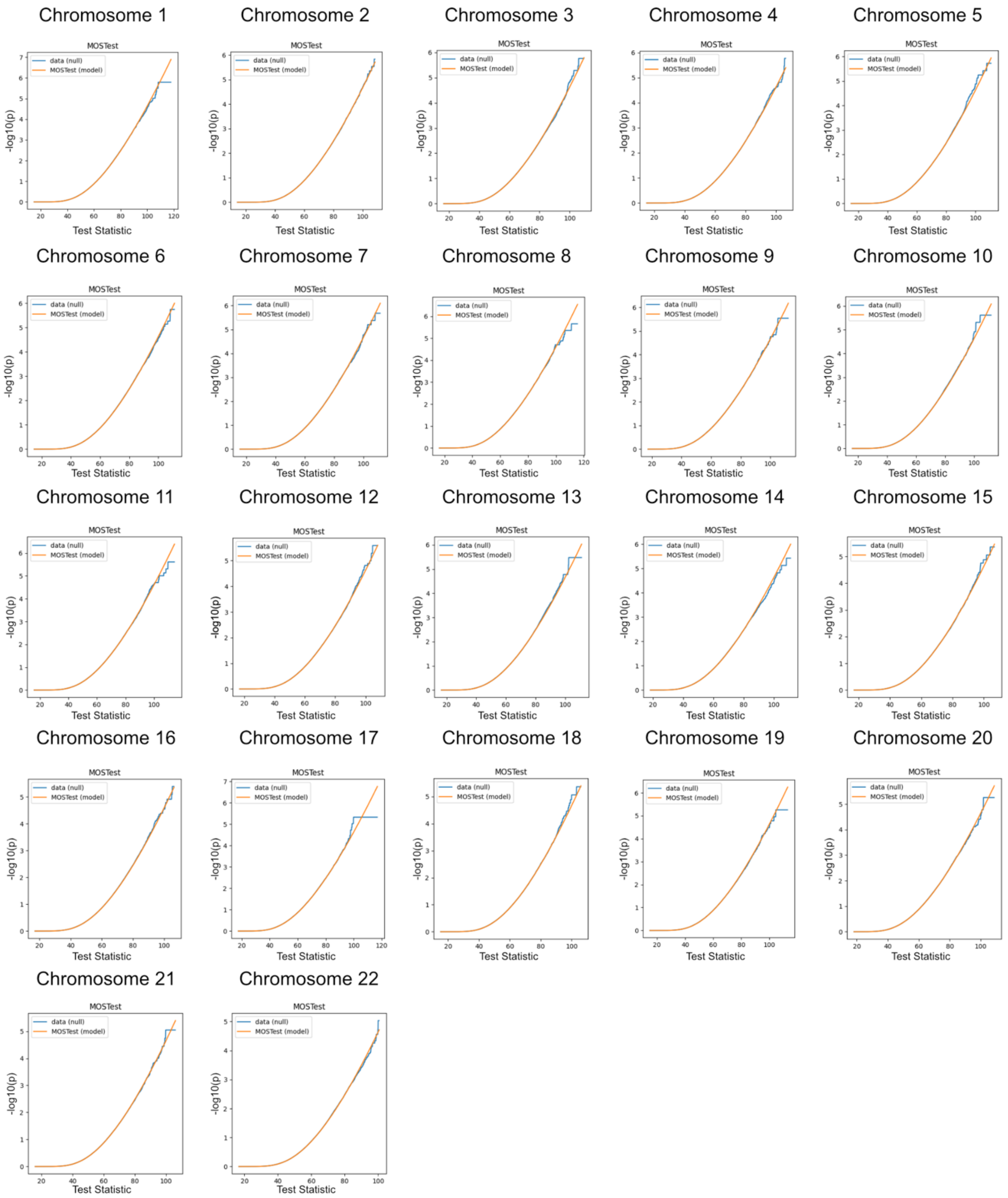


**Fig. S22. Chromosome-wise Comparison of Empirical and Analytical MOSTest Results Under the Null Hypothesis for right Cerebellum Cortex.** See description of Fig. S14.

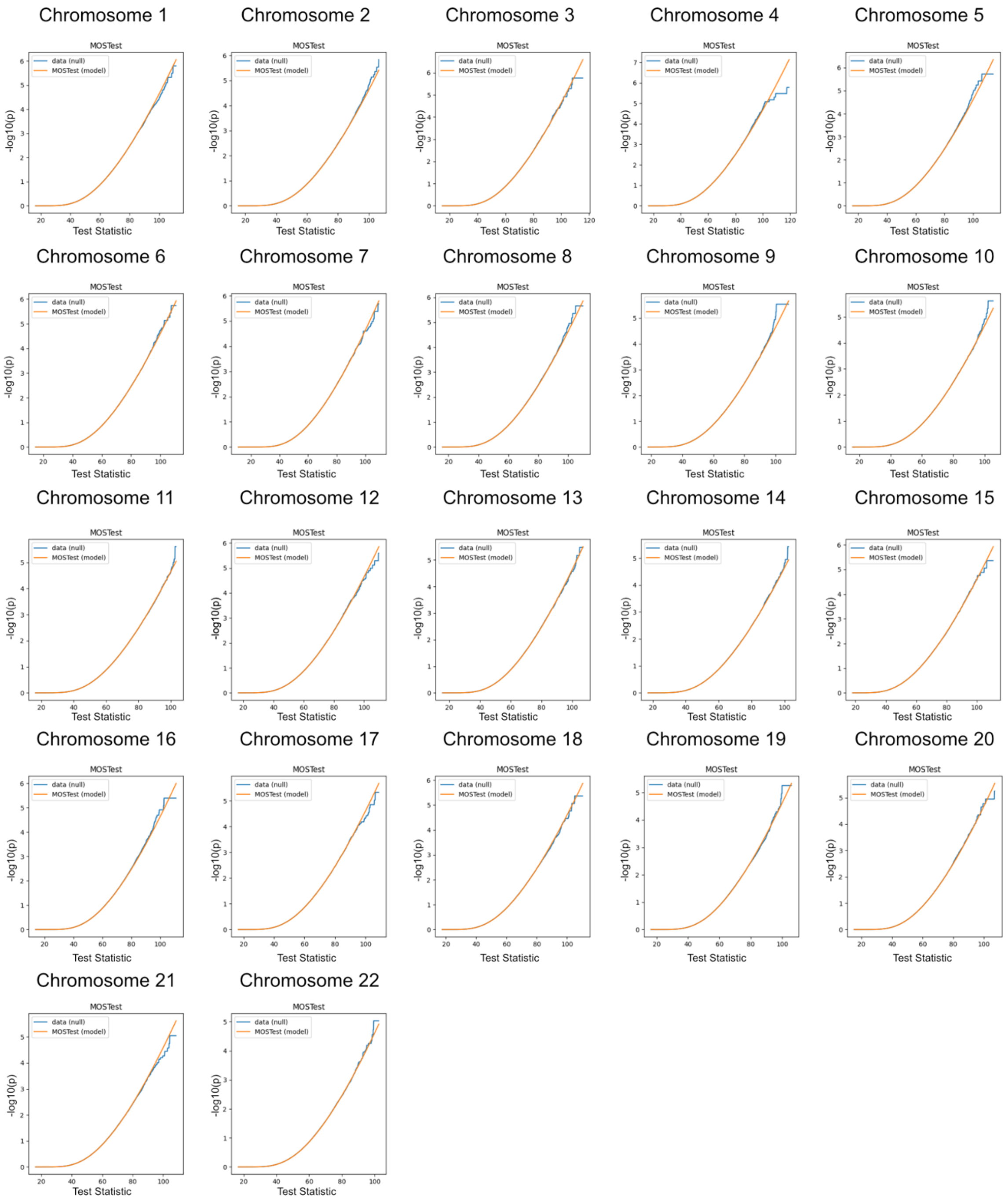


**Fig. S23. Chromosome-wise Comparison of Empirical and Analytical MOSTest Results Under the Null Hypothesis for left Cerebellum White Matter.** See description of Fig. S14.

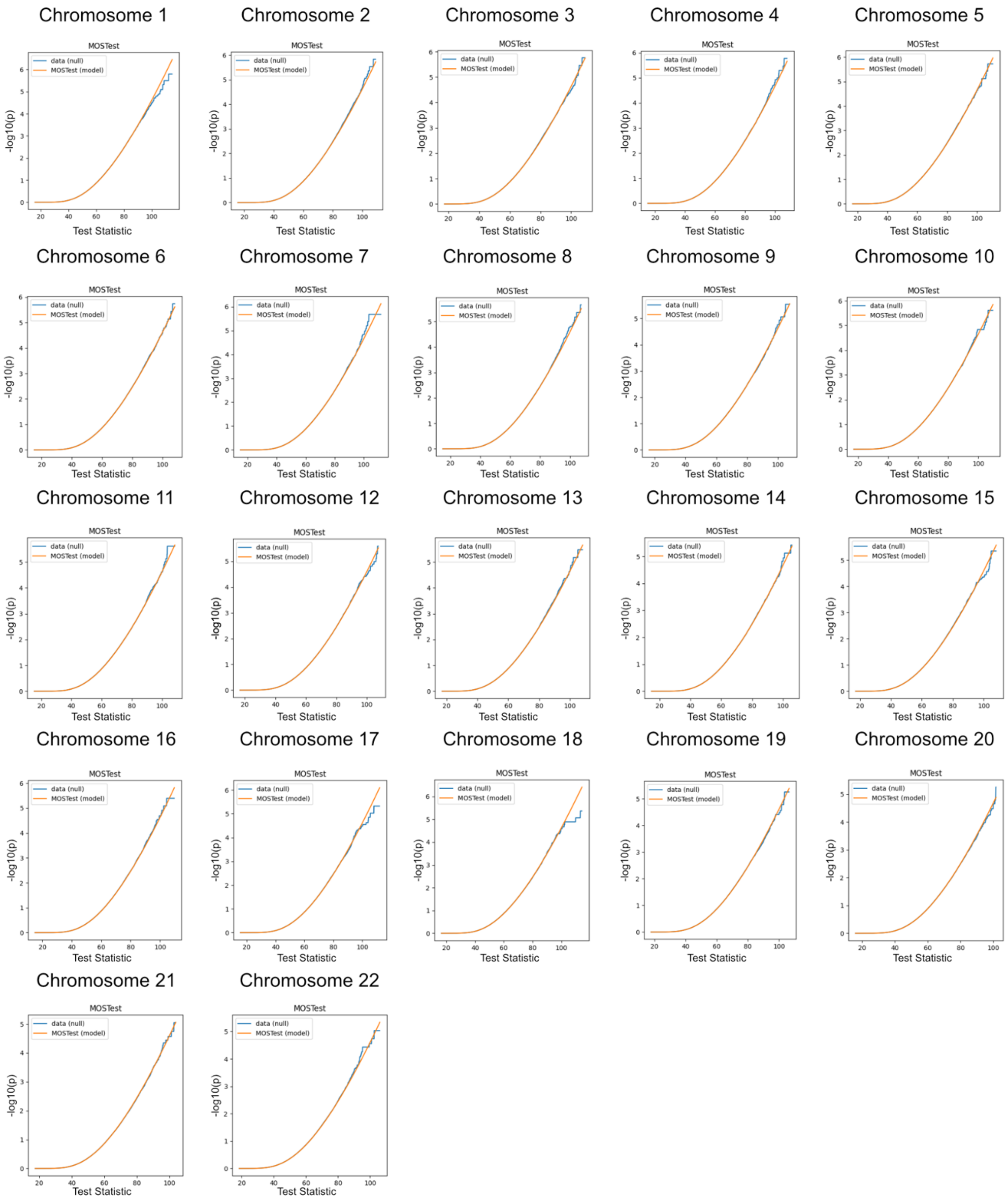


**Fig. S24. Chromosome-wise Comparison of Empirical and Analytical MOSTest Results Under the Null Hypothesis for right Cerebellum White Matter.** See description of Fig. S14.

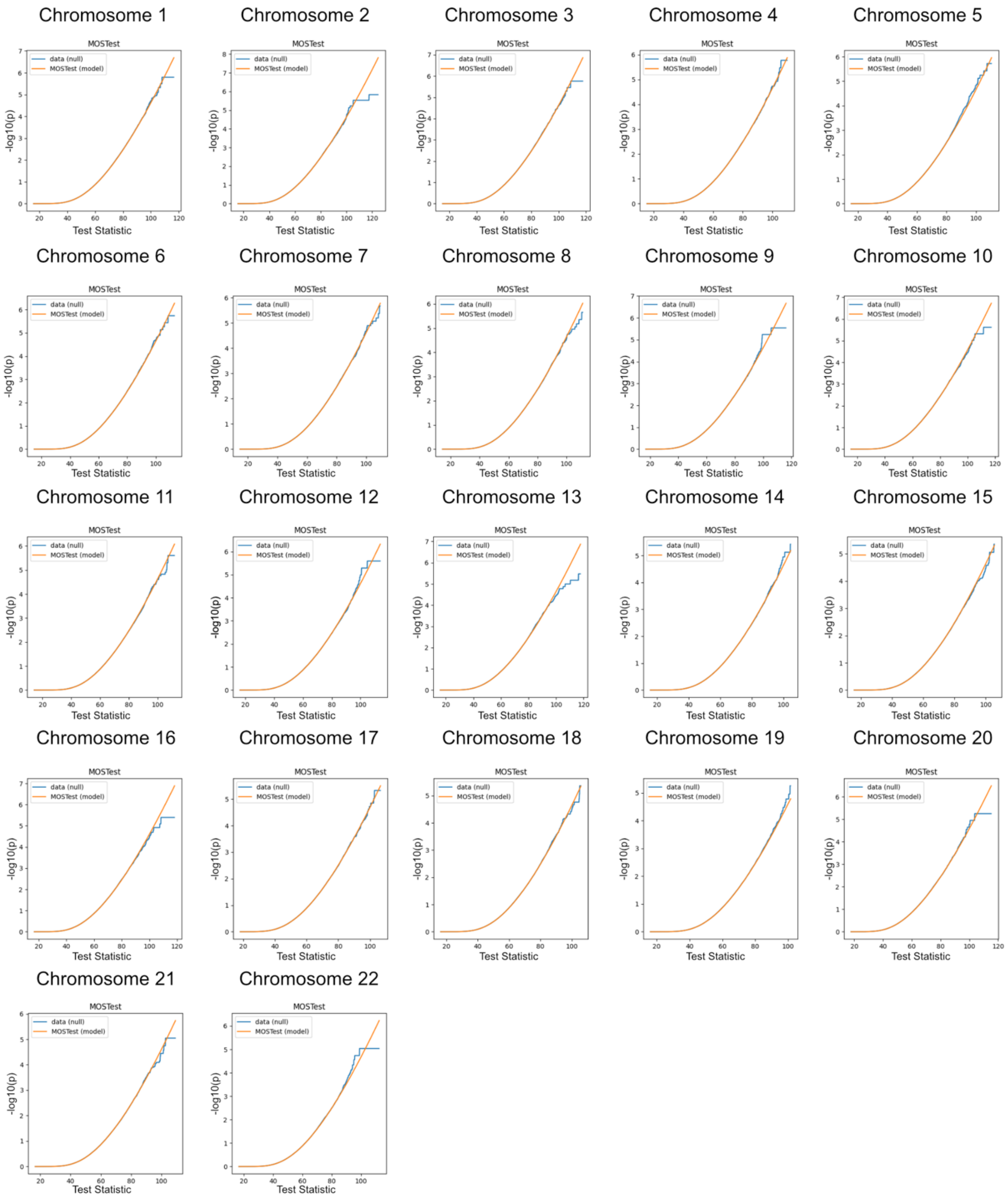


**Fig. S25. Chromosome-wise Comparison of Empirical and Analytical MOSTest Results Under the Null Hypothesis for left Hippocampus.** See description of Fig. S14.

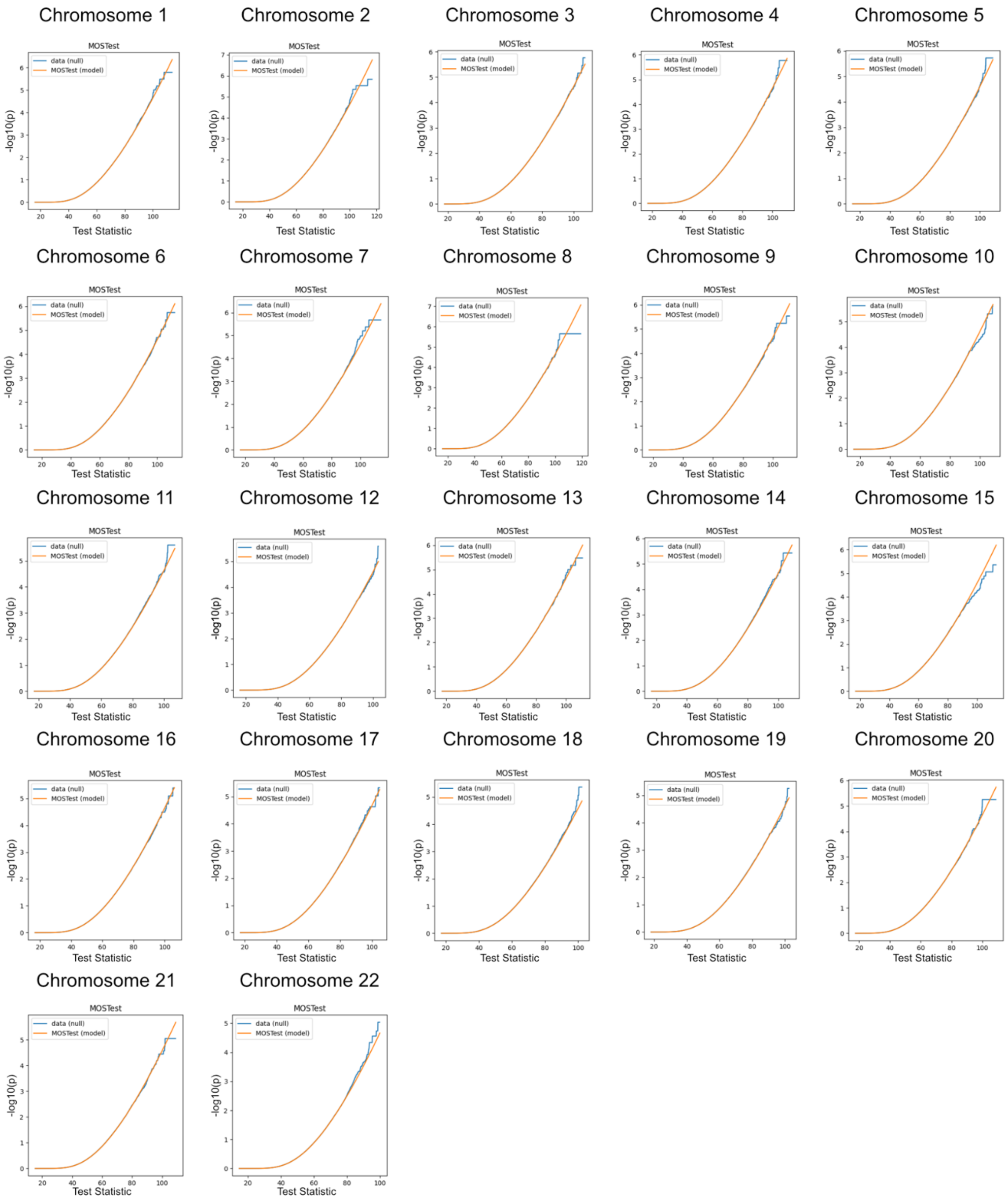


**Fig. S26. Chromosome-wise Comparison of Empirical and Analytical MOSTest Results Under the Null Hypothesis for right Hippocampus.** See description of Fig. S14.

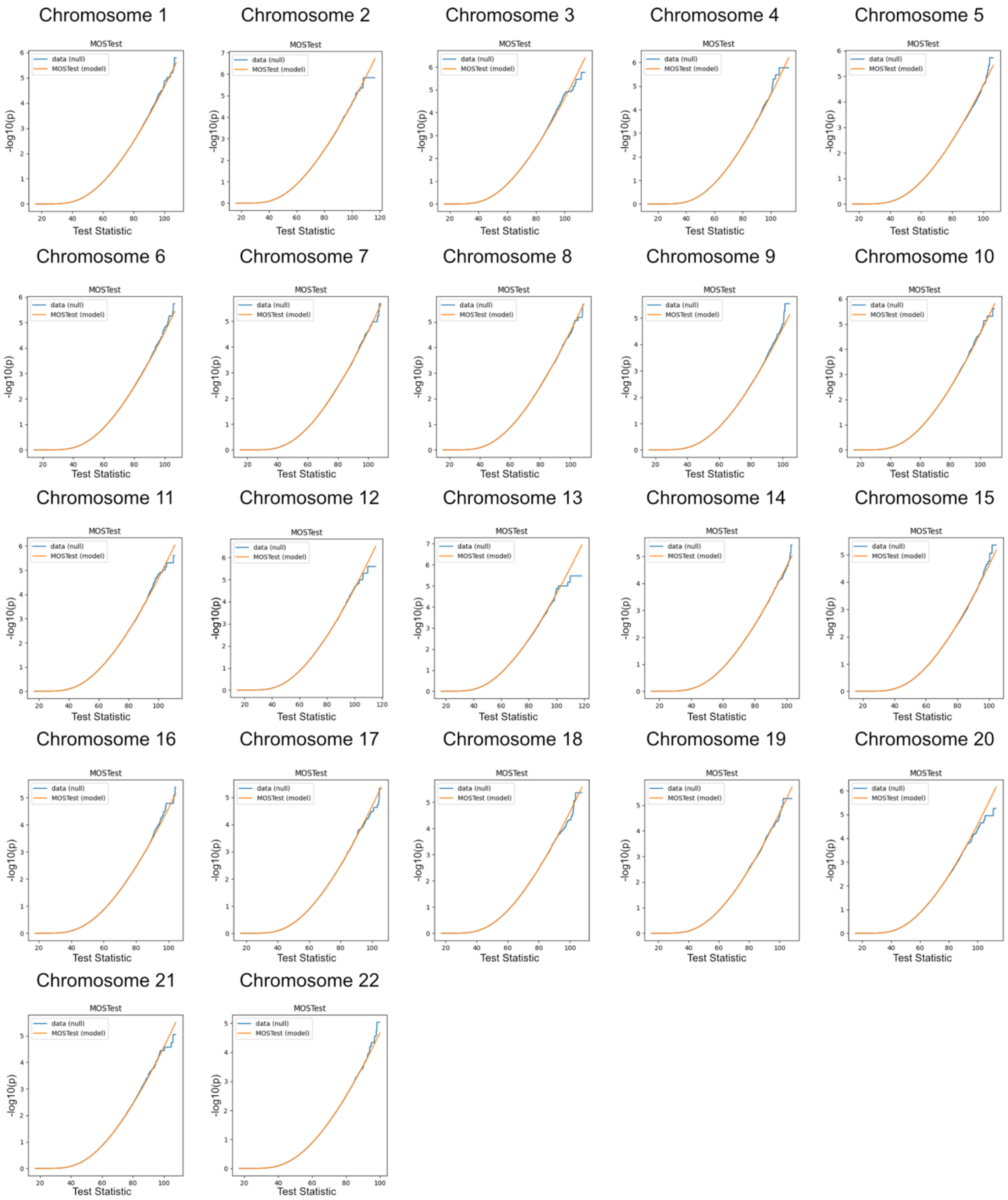


**Fig. S27. Chromosome-wise Comparison of Empirical and Analytical MOSTest Results Under the Null Hypothesis for left Pallidum.** See description of Fig. S4.

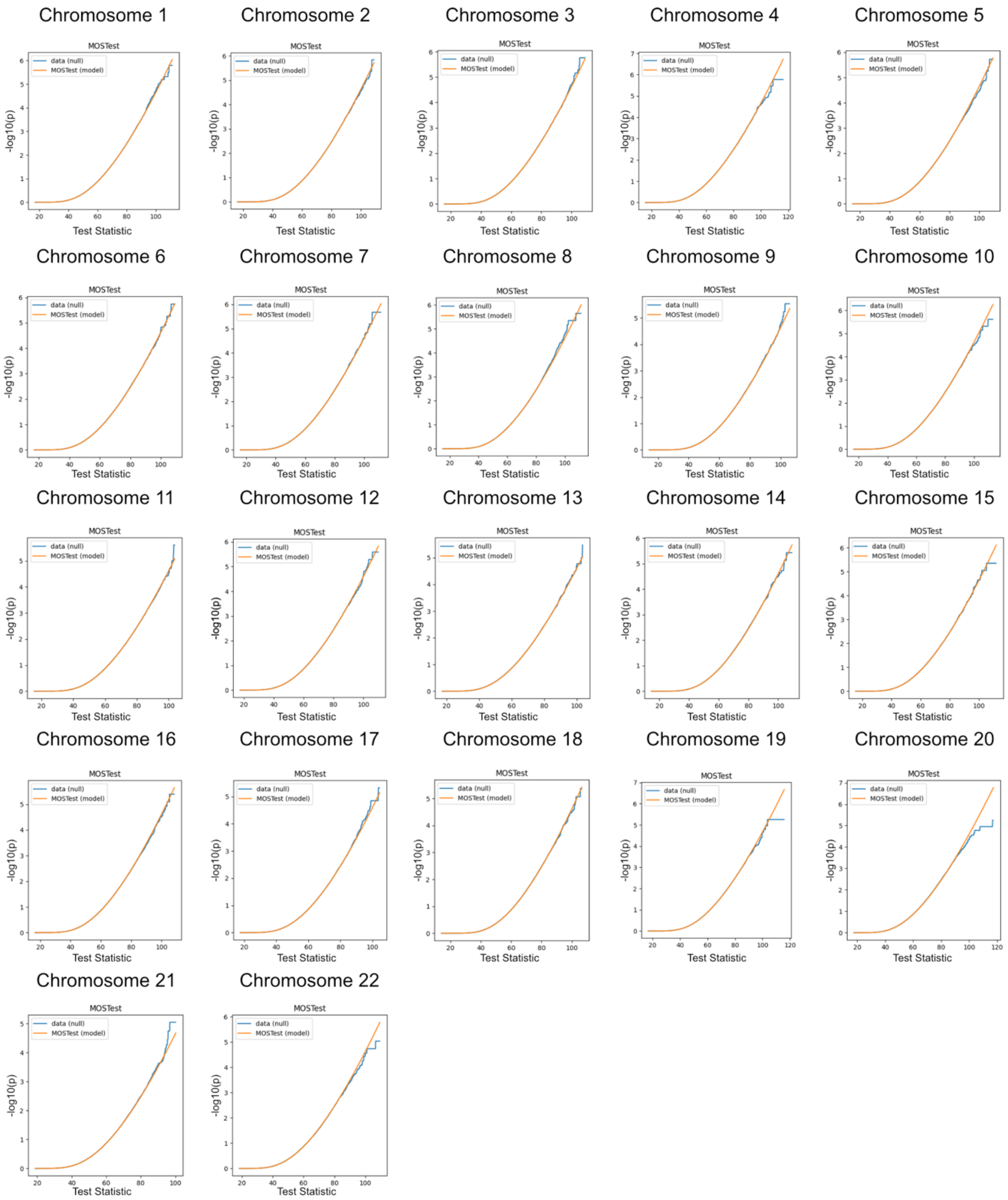


**Fig. S28. Chromosome-wise Comparison of Empirical and Analytical MOSTest Results Under the Null Hypothesis for right Pallidum.** See description of Fig. S14.

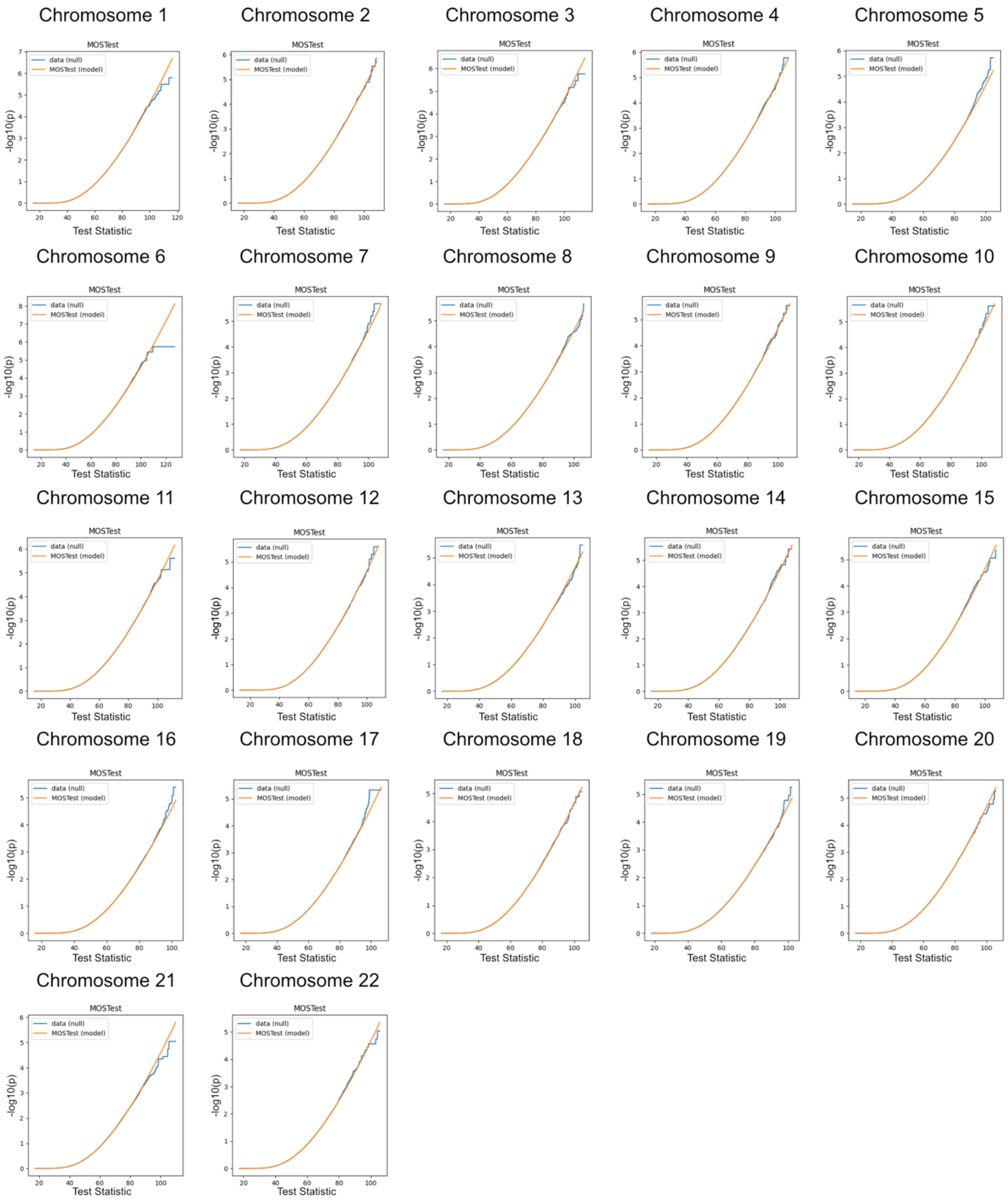


**Fig. S29. Chromosome-wise Comparison of Empirical and Analytical MOSTest Results Under the Null Hypothesis for left Putamen.** See description of Fig. S14.

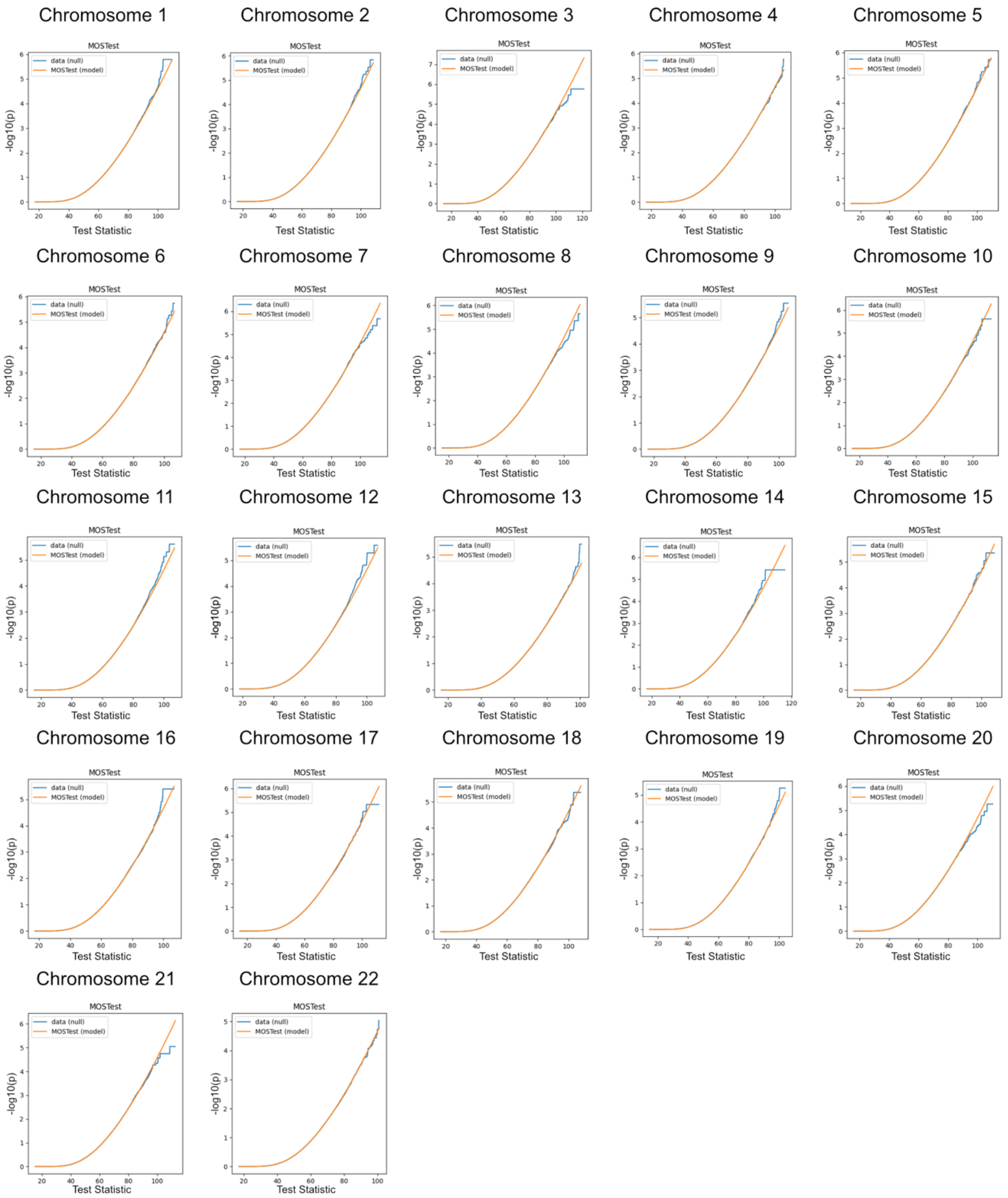


**Fig. S30. Chromosome-wise Comparison of Empirical and Analytical MOSTest Results Under the Null Hypothesis for right Putamen.** See description of Fig. S14.

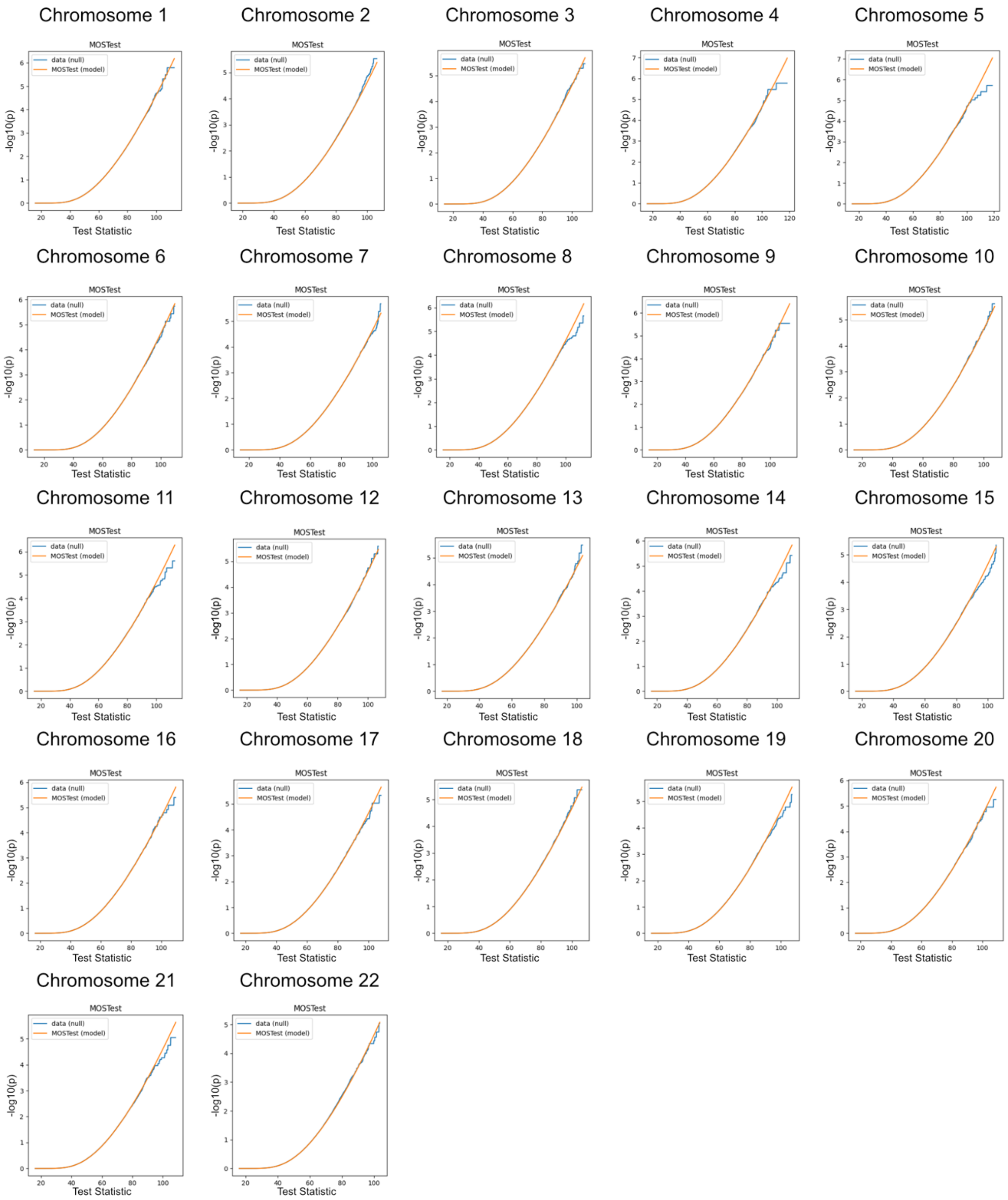


**Fig. S31 Chromosome-wise Comparison of Empirical and Analytical MOSTest Results Under the Null Hypothesis for left Thalamus Proper.** See description of Fig. S14.

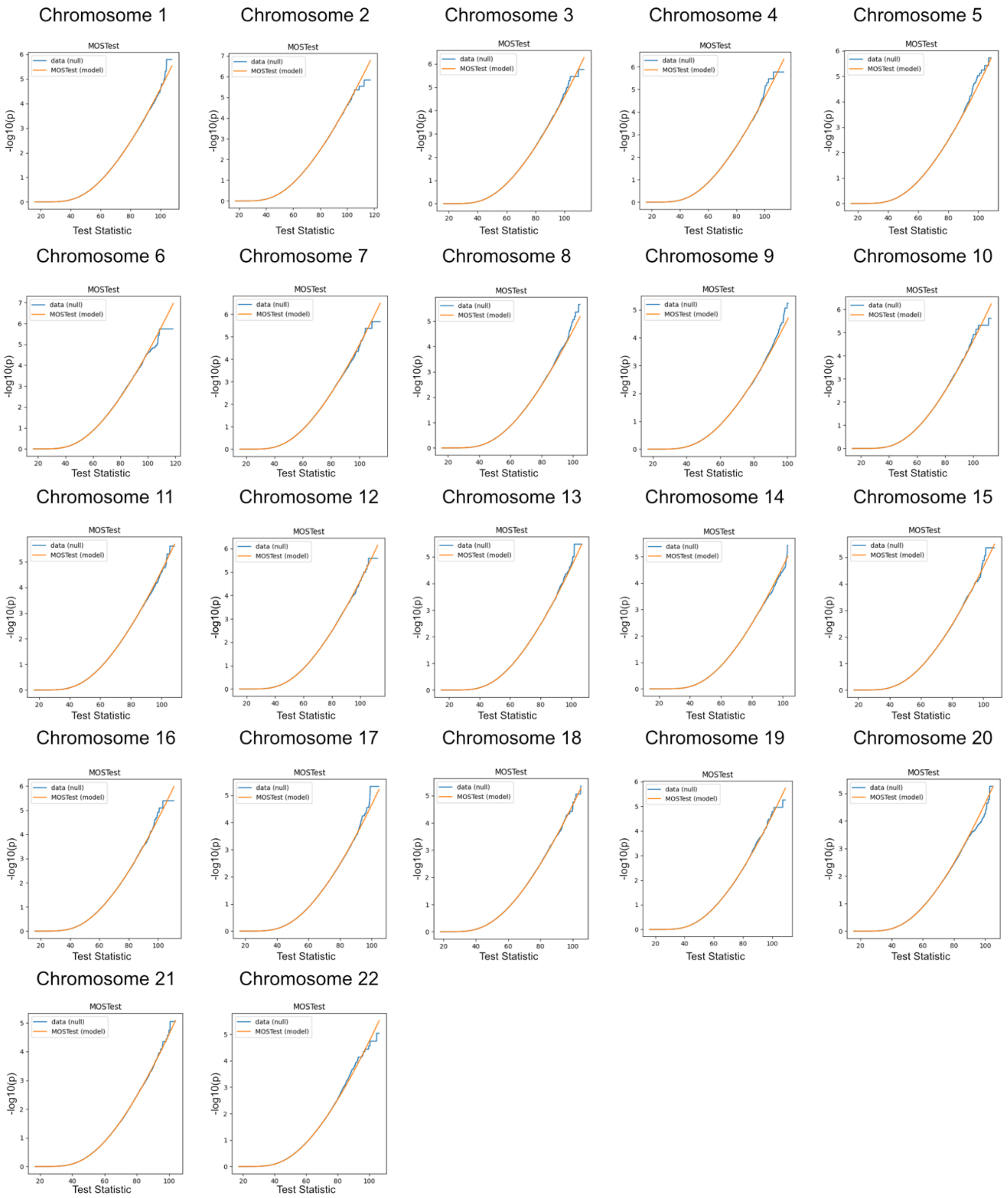


**Fig. S32. Chromosome-wise Comparison of Empirical and Analytical MOSTest Results Under the Null Hypothesis for right Thalamus Proper.** See description of Fig. S14.

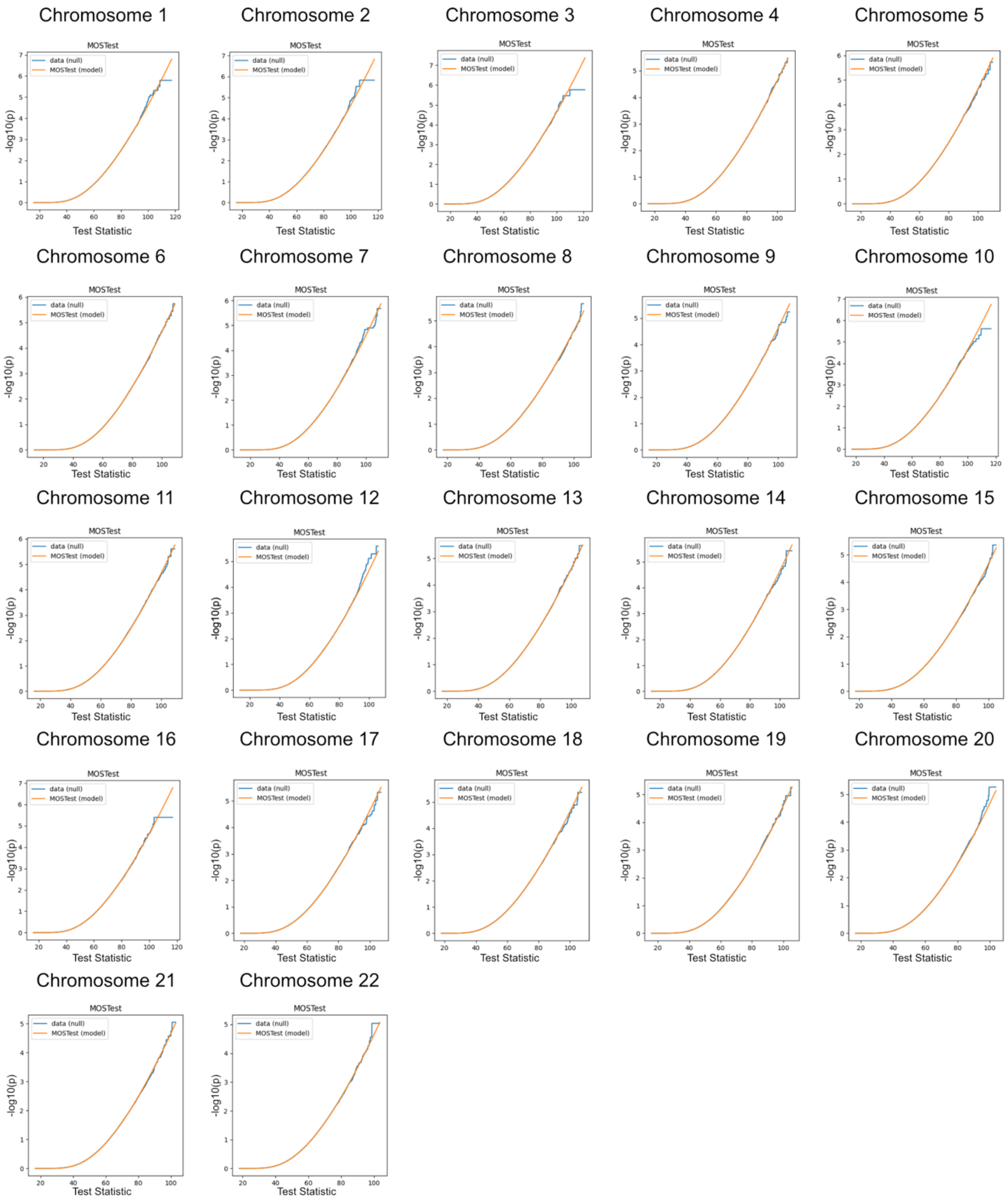


**Fig. S33. Chromosome-wise Comparison of Empirical and Analytical MOSTest Results Under the Null Hypothesis for left Ventral DC.** See description of Fig. S14.

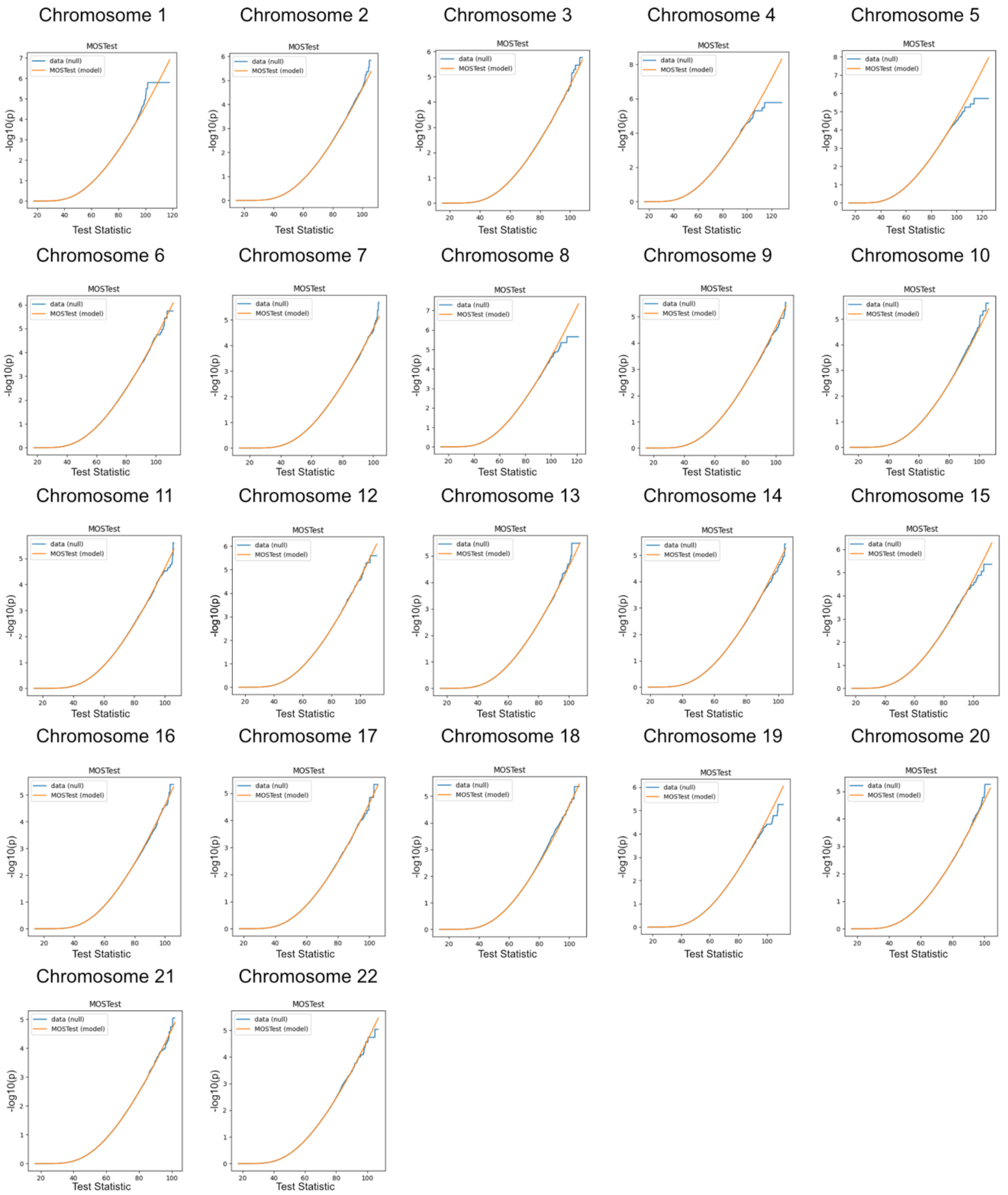


**Fig. S34. Chromosome-wise Comparison of Empirical and Analytical MOSTest Results Under the Null Hypothesis for right Ventral DC.** See description of Fig. S14.

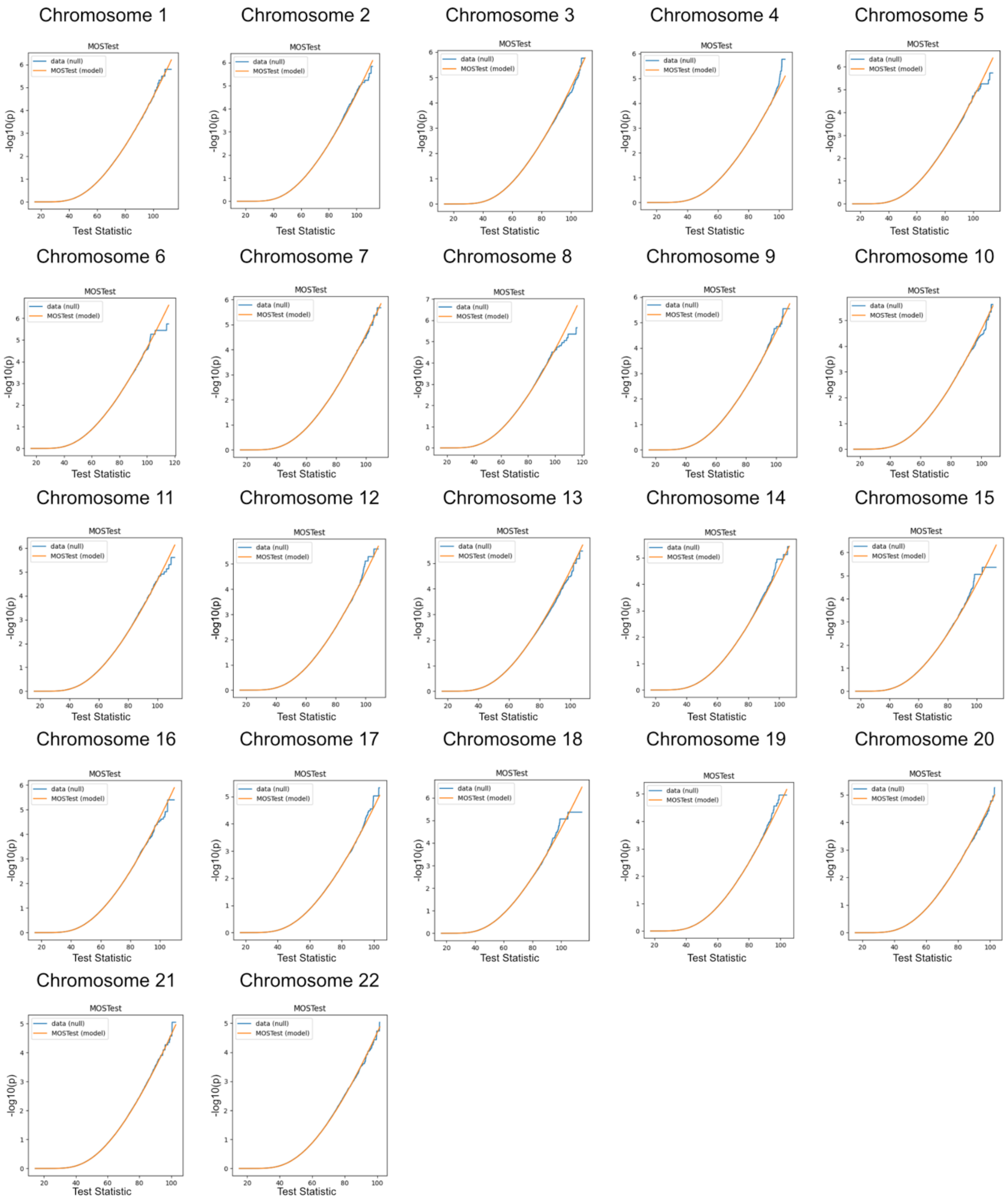


**Fig. S35. Chromosome-wise Comparison of Empirical and Analytical MOSTest Results Under the Null Hypothesis for 4th Ventricle.** See description of Fig. S14.

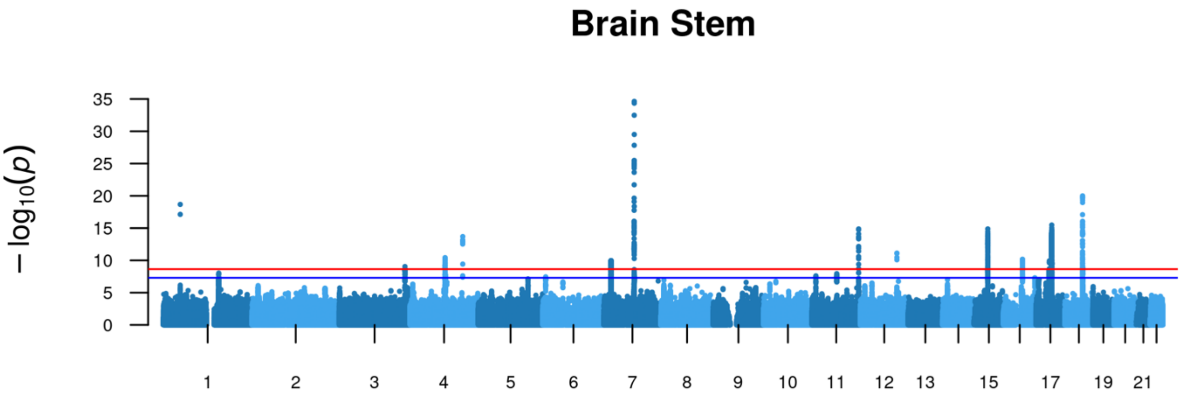


**Fig. S36. Robustness Analysis.** Manhattan plot of MOSTest results for brain stem LBS with height as additional covariate. Chromosomal locations of the SNPs are indicated on the x-axis and -log10 scaled p-values on the y-axis. Blue and red lines indicate standard genome-wide significance (p=5E-8) and genome-wide significance after Bonferroni correction for 22 brain structures (p=5E-8/22), respectively.

**Supplementary Tables**

An external Excel file contains all supplementary tables as follows:

**Table S1.** *Association of Independent Significant SNPs with GWAS Catalog Traits.* The individual columns show the following: *SNP*, 80 independent significant SNPs of the present study that were indicated by the analysis (MOSTest on LBS) of at least one brain structure; *LD_SNP1* and *LD_SNP2*, SNPs selected as independent significant by FUMA in at least one brain structure but belonging to the same locus ($r2 \geq 0.6$) as the variant in the SNP column; *GWAS catalog category*, whether GWAS catalog brain shape traits ("brain") defined as in Table S20, only other GWAS catalog traits ("onlyOtherTraits") or no GWAS catalog traits at all have been significantly associated with SNPs in the first three columns or proxies of them ($r2 \geq 0.6$). The subsequent columns relate the information in the fourth column to the specific brain structure(s) that were analyzed in the present study. *NA* indicates that there was no significant association after Bonferroni correction.

**Table S2**. *Functional Annotation of the 80 Independent Significant SNPs.* Results from FUMA with non-effect allele (NEA), effect allele (EA), effect allele frequency (EAF) based on the reference panel (see Methods), distance to nearest gene (dist), and functional annotation (func) according to ANNOVAR.

**Table S3.** *Exonic Nonsynonymous SNPs in LD with the 80 Independent SNPs - extension of Table 1.* Results from FUMA with non-effect allele (NEA), effect allele (EA), effect allele frequency (EAF), minimum p-value in all GWASs in the present study (min_gwasP), linkage disequilibrium (r2) to the independent SNP (IndSigSNP) in the respective brain structure GWAS (brainstructure), and exon number (exon) of the gene's transcript as chosen by ANNOVAR.

**Table S4.** *EQTL Associations of rs1687225 that is significantly associated with Brain Stem LBS.* Results from FUMA analysis of brain-related databases (db) with p-values (p) and statistics from the eQTL association.

**Table S5.** *ExNS eQTLs of Protein Coding Genes in Genomic Locus 12 of Brain Stem LBS GWAS with rs568589031 as Lead SNP.* Results from FUMA as in Table S4, eQTL gene (eqtlGene) and the positional gene (posGene) as obtained by ANNOVAR.

**Table S6.** *P-values of the 80 Independent Significant SNPs.* Table contains SNPs (rsID) with effect allele (EA), non-effect allele (NEA), and their p-values in all brain structure GWASs with the number of significant structures under different significance thresholds. P-values are colored green ($p < 5E\text{-}8/22$), blue ($5E\text{-}8/22 < p < 0.05/(80*22)$), light pink ($p > 0.05/(80*22)$ and p of contralateral structure $< 0.05/(80*22)$), dark pink ($p > 0.05$ and contralateral $p < 0.05/(80*22)$) or grey ($p > 0.05$).

**Table S7.** *Results from GWAS Catalog for rs6658111 and rs12146713 Using LDTrait.*

**Table S8.** *Prioritized Genes Across all Brain Structures.* Binary coding if the gene was prioritized for the respective structures (1: True, 0: False).

**Table S9.** *Pearson Correlations Between Polygenic Risk Scores – Correlation coefficients.*

Table contains polygenic risk scores for Alzheimer's disease (AD), bipolar disorder (BD), ischemic stroke (ISS), multiple sclerosis (MS), Parkinson's disease (PD), schizophrenia (SCZ), and alcohol use disorder (ALC).

**Table S10.** *Pearson Correlations Between Polygenic Risk Scores - Unadjusted P-values.* See description for Table S9.

**Table S11.** *Pearson Correlations Between Polygenic Risk Scores – Test Statistics.* See description for Table S9.

**Table S12.** *Extended heritability results of single brain structures.* Table extends Table 2 with data on heritability variance (h^2_var) and results from the heritability Wald test (WaldStats, WaldP, WaldP_log10).

**Table S13.** *Extended heritability results of combined brain structures.* Table extends Table 2 with results from the heritability Wald test of each combined structure (WaldStats, WaldP, WaldP_log10) and from the Wald test on difference in heritability between our results and those of (*6*) including absolute difference of heritability estimates (her_diff) and p-values of the test (waldP_diff).

**Table S14.** *Results of the 80 Independent Significant SNPs in Replication Data Set.* Results of all Bonferroni significant associations of the 80 independent significant SNPs (148 occurrences in total) are listed. Table shows the results with base pair (BP) and chromosome (CHR) position, effect allele (A1), non-effect allele (A2), p-value in discovery data set (PVAL_disc), in replication set (PVAL_rep), and in replication set with FDR correction for all associations in the respective brain structure (PVAL_rep_fdr). SNP is marked with a "*" for nominal significance (PVAL_rep < 0.05) or with "**" for FDR-corrected significance (PVAL_rep_fdr < 0.05).

**Table S15.** *Results of the 80 Independent Significant SNPs in Discovery Data Sets.* See description of Table S14 with rep_i describing discovery data set *i*.

**Table S16.** *Comparison of Amygdala Variants.* Table shows 12 loci from (*100*) with effect allele (A1) and non-effect allele (A2). Next, p-values and FDR-adjusted p-values are listed for left and right amygdala normalized and regressed as in the main study (*_left_vol_surf, *_right_vol_surf) and for left and right amygdala not normalized and without surface as covariate (*_left_no_norm, *_right_no_norm).

**Table S17.** *Results of the 80 Independent Significant SNPs in GWAS with Volume as Additional Covariate.* See description of Table S14. P-values of GWAS with volume as additional covariate (PVAL_rep) are listed. SNPs are marked additionally with "***" for suggestive significance (Bonferroni correction for 80 SNPs and 22 brain structures: PVAL_rep < 0.05/(80*22)), with "!" for genome-wide significance (PVAL_rep < 5E-8) and with "!!" for Bonferroni-corrected genome-wide significance (PVAL_rep < 5E-8/22). The genomic loci were defined in the FUMA analysis of the discovery analysis.

**Table S18.** *Mean Loadings of CCA Results.* The table shows the mean of all loadings for each canonical correlation between polygenic risk score and brain structure eigenvalue spectrum. In parentheses, the number of negative loadings is stated. NA indicates zero negative loadings.

**Table S19.** *Results from Canonical Correlation Analysis.* Table contains canonical correlations (*_CCA) between each polygenic risk score (abbreviations see description of Table S9) and each brain structure and the corresponding unadjusted p-value of the correlation (*_pval).

**Table S20.** *Reported Brain Shape Related Traits from GWAS Catalog*. Table lists all traits for brain volume measurements, cortical surface area measurements and cortical thickness as extracted from GWAS catalog. The last column lists additional brain shape traits from GWAS catalog, which are significantly associated with SNPs (or their proxies) that have also been found to be significant in the present study. All these traits were used to categorize SNPs into the "brain" category in Table S1.

**Table S21.** *Coordinates of Principal Components 1-3 of log10-scaled P-values of 80 Independent Significant SNPs.*

**Table S22.** *Replication of 37 Independent Significant SNPs of Brain Stem GWAS with Height as Additional Covariate.* Description see Table S17.
